# Pan-Omics Fusion and Machine Learning Unveil Congenital Tooth Agenesis-Ecto-mesodermal Diseases Link and Biomarker Discovery

**DOI:** 10.1101/2025.03.09.25323497

**Authors:** Prashant Ranjan, Chandra Devi, Vinay Kumar Srivastava, Meenakshi Chandel, Garima Jain, Parimal Das

## Abstract

**Background:** Congenital tooth agenesis (CTA) is a common developmental anomaly with complex genetic and molecular mechanisms. Previous studies have primarily focused on candidate gene mutations, often lacking a pan-omics perspective.

**Methods:** This study integrates metabolomics, proteomics, microarray, and genomics with machine learning to identify biomarkers and elucidate disease mechanisms. A random forest-based classification achieved high AUC-ROC scores (0.95 for proteomics, 0.98 for metabolomics), validating the biomarker discovery framework.

**Results:** Several biomarkers were identified in this study that enhance our understanding of CTA. Furthermore, our findings reveal a significant association between CTA and ecto-mesodermal diseases, which has not been extensively explored before. Notably, 24 dual-expression genes were expressed in both pre- and post-developmental stages, suggesting a regulatory role in tooth integrity, repair, and homeostasis. Metabolomics analysis revealed 28 upregulated and 17 downregulated metabolites uniquely associated with CTA. Key metabolic alterations involved nucleotide metabolism, purine metabolism, oxidative stress, and Wnt signaling. High-performing metabolites (AUC ≥ 0.90), including PEG n5 (0.99), PEG n6 (0.98), PEG-4 (0.97), PEG n7 (0.96), PEG n8 (0.95), caffeine (0.94), hydroxycaproic (0.91) and alpha-aspartylphenylalanine (0.90) demonstrated strong diagnostic potential. CTA patients showed 292 unique metabolites vs. 238 in controls, indicating metabolic pathway alterations. Proteomic analysis identified 76 upregulated and 33 downregulated genes, with key biomarkers [*SERPINA1* (0.92), *PZP* (0.90), *FGA* (0.91), *TLN1*(0.94), *FGB* (0.95)] displaying AUC-ROC ≥ 0.90. Pan-omics fusion followed by STRING analysis identified 20 central hub genes strongly correlated with congenital tooth agenesis signaling.

**Conclusion:** This study pioneers the systemic association of CTA with ecto-mesodermal diseases, revealing novel signatures, disrupted pathways, and therapeutic targets.

**Graphical Abstract:** 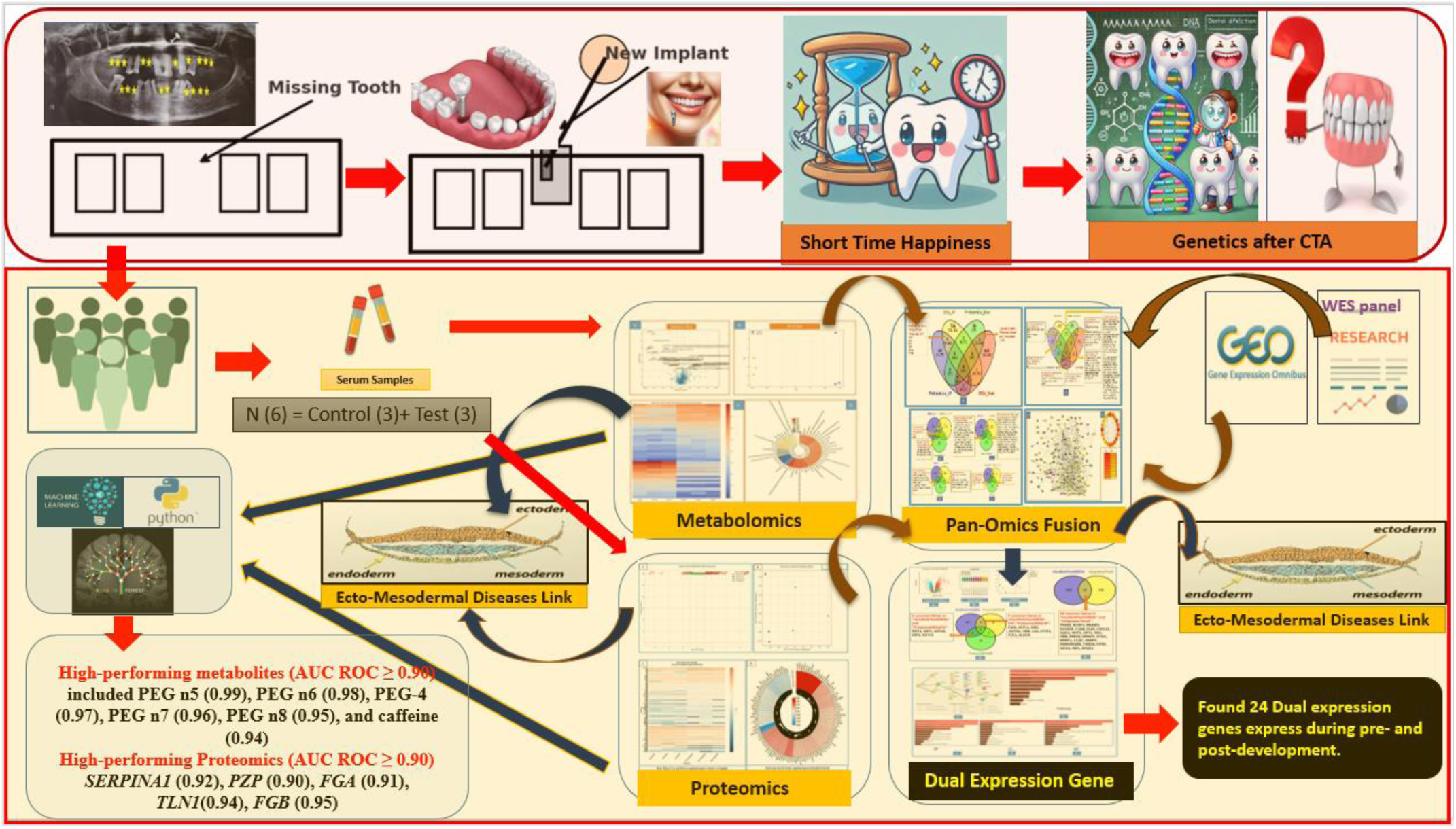

## 1. Introduction

Congenital tooth agenesis (CTA) is a prevalent dental anomaly characterized by the absence of one or more teeth at birth. This condition not only affects dental health and aesthetics but also has profound implications for overall oral function (Vastardis, 2000). Understanding the molecular underpinnings of tooth agenesis is crucial for the development of targeted therapeutic interventions and preventive strategies.

Tooth development is orchestrated by highly conserved signaling pathways, such as Wnt, BMP, FGF, and EDA-NF-κB, which mediate interactions between epithelial and mesenchymal tissues during odontogenesis (Biggs & Mikkola, 2014). Genetic mutations in key regulatory genes, including *PAX9*, *MSX1*, *EDA*, and *WNT10A*, have been implicated in CTA. Additionally, recent studies have highlighted the pivotal roles of microRNAs (miRNAs) as post-transcriptional regulators of gene expression, further enriching our understanding of the regulatory networks involved (Ranjan Jr et al., 2024).

Advancements in high-throughput technologies have enabled comprehensive profiling of biological systems, generating data across various molecular layers, including genomics, transcriptomics, proteomics, and metabolomics. However, understanding complex biological processes, such as tooth development and its associated anomalies, requires more than analyzing individual omics datasets in isolation. The integration of Pan-omics data provides a systems-level perspective, bridging the gap between different molecular layers to uncover the intricate regulatory mechanisms governing these processes (Huang et al., 2023)). Discovering biomarkers for complicated diseases is now easier because to this change, which is essential for improving disease diagnosis, prognosis, and therapy. The integration of pan-omics data from many sources has enabled the identification of reliable and accurate biomarkers. Biomarkers can be extracted from this pan-omics data, and researchers can use these biomarkers to create a single predictive model, multiple models based on various biomarkers combined, or a mixed strategy for integration (H. Yang et al., 2023).

Proteomics provides insights into the signaling cascades and protein-protein interactions involved in tooth development and agenesis by facilitating the large-scale research of proteins, their structures, and activities. The study of metabolites and metabolic pathways, or metabolomics, offers a glimpse of the biochemical processes occurring within cells, tissues, and organisms. This helps to clarify the metabolic changes linked to tooth agenesis.

Thousands of gene expressions may be examined simultaneously using microarray analysis, which makes it easier to identify regulatory networks and differentially expressed genes that contribute to congenital tooth agenesis. Conversely, whole exome sequencing (WES) looks at the genome’s coding sections to find genetic variations that could be crucial to the cause of tooth agenesis.

This comprehensive pan-omics exploration aims to elucidate the molecular drivers of CTA by integrating data from proteomics, metabolomics, microarray analysis, and WES. By leveraging the strengths of these diverse technologies, we strive to construct a detailed molecular map of tooth agenesis, identifying biomarkers, diseases association, dual expression biomarkers (expressed during and post-embryonic stages), and potential therapeutic targets. Additionally, this study seeks to uncover the association between congenital tooth agenesis and other diseases, shedding light on their underlying origins

## 2. Methodology

### 2.1 Sample Collection

This study followed to the principles outlined in the Declaration of Helsinki. Written informed consent was obtained from all participants or, in the case of minors, from their parent or legal guardian. Serum samples were obtained from 3 CTA patients (Den 31, 32 and 33) (average Age 35.0 ± 5.77 years) and 3 healthy controls (Control 1,2 and 3) (average Age 42.3 ± 4.34 years) after informed consent. Blood samples were collected and processed to obtain serum. The serum was stored at -80°C until further analysis. Ethical approval for the study was obtained from the Ethics Committee of the Institute of Science, Banaras Hindu University, India.

### 2.2 Sample Groups

It is important to note that the 3 serum samples of CTA patients and 3 healthy control participants used in the proteomic and metabolomics study were different from those in the WES study (Ranjan et al., 2024). The microarray gene expression data, as mentioned (Ranjan et al., 2025), was obtained from publicly available database to provide additional context and insights for the omics-based analyses.

### 2.3 Proteome and Metabolome Analysis

Before being evaluated, serum samples were thawed on ice. Proteins in the serum samples were precipitated using methanol (LC-MS grade) as an organic solvent at a 1:3 ratio (sample: solvent), and the mixture was well mixed by vortexing. To improve protein precipitation, the samples were then incubated for 30 minutes at -20°C. Following incubation, samples were centrifuged at 12,000 x g for 10 minutes at 4°C. The metabolite-containing supernatant was then transferred to new tubes. Using a Speed Vac vacuum concentrator, the supernatant was meticulously dried. After drying, the samples were reconstituted in water of LC-MS quality and vortexed to ensure full dissolution. Following a centrifugation to eliminate any particles, the reconstituted samples were filtered using a 0.22 µm syringe filter before being subjected to an LC-MS analysis.

A Dionex UltiMate 3000 RSUHPLC system for ultra-high-performance liquid chromatography (UHPLC) was utilized in conjunction with an Orbitrap Eclipse Tribrid Mass Spectrometer (Thermo Fisher Scientific) for mass spectrometry analysis. A Hypersil GOLDTM C18 HPLC column (1.9 µm particle size, 2.1 mm diameter, 100 mm length) was used to separate the metabolites. Solvents A, B, and C were 100% water and 0.1% formic acid, 80% acetonitrile and 0.1% formic acid, and 100% methanol and 0.1% formic acid, respectively. The run time was 30 minutes, the injection volume was 5 μl, and the flow rate was 0.3 ml/min. A mass spectrometer was linked to the column output using electrospray ionization, or H-ESI. The chemicals that were channelized using Orbitrap were ionized utilizing both the negative and positive modes of H-ESI.

### 2.4 Data acquisition and Statistical Analysis

Data analyses were done using default parameters of “Compound Discoverer 3.3.3.200” software using online databases to identify differentially expressed proteins and metabolites between CTA patients and healthy controls. The untargeted metabolomics workflow included retention time alignment, detection of unknown compounds, and compound grouping across all samples. The software predicted elemental compositions, filled data gaps, and removed chemical background using blank samples. Identified compounds were annotated using the mzCloud (ddMS2) and ChemSpider databases. The biological pathway mapping was done using Metabolika. Normalization was applied using control (QC) samples. The differential analysis (ANOVA), p-values, adjusted p-values, ratios, fold change, CV, etc) were calculated. A false discovery rate (FDR) correction was applied to account for multiple comparisons. Human Metabolome Database (HMDB) was used for metabolite identification.

### 2.4 Volcano Plot Generation

The proteins that were expressed differentially in the Sample and Control were visualized using a Python script. We retrieved q-values (qvals) and log2 fold change (log2fc) from protein expression data imported from a CSV file. To cope with q-values of 0, a constant of 1e-300 was employed. A scatter plot with color coding based on thresholds was created using Matplotlib. Proteins with non-significant high fold-change (|log2fc| > 1, q ≥ 0.05) were blue, proteins with significant low fold-change (|log2fc| ≤ 1, q < 0.05) were green, and other proteins were grey. For simple viewing, negative log10(q) was placed on the y-axis. Significant proteins were marked on the figure, and threshold lines (|log2fc| = ±1, q = 0.05) were added for reference.

To display the findings of the differential analysis for the metabolomics data, a volcano graphic was created. The y-axis shows the -log10 p-value, while the x-axis shows the log2 fold change. Significant differences between groups can be identified since each data point correlates to a metabolite. Cut-off values for the p-value and fold change, respectively, are shown by dashed horizontal and vertical lines, which represent the significance levels that were used. To differentiate between characteristics that were highly upregulated and downregulated, metabolites that above these levels were highlighted.

The script had a legend that represented protein classifications and used Seaborn (Sial et al., 2021) for consistent aesthetics. Total proteins, significantly modified proteins, and their up- and down- regulation were compiled in the analysis.

### 2.5 PCA Plot

PCA was conducted using the scikit-learn library (Kramer, 2016) to reduce the dataset’s dimensionality and capture the maximum variance. The analysis extracted two principal components (PC1 and PC2) for simplified visualization and meaningful interpretation. Scatter plots were created to illustrate the PCA results, with each data point representing either a healthy control or a CTA sample. Unique colors and annotations were employed to distinguish between groups and identify clusters or patterns effectively. Python (v3.12) (Python, 2021) was utilized for the analysis, with pandas handling data preprocessing and organization. The matplotlib library (Hunter & Dale, 2007)was used for generating clear and informative visualizations.

### 2.6 HeatmapVisualization

Control and sample area values were extracted for analysis. Control values were obtained from columns labelled as Control 1, Control 2, and Control 3, while sample values were derived from Den 31, 32 and 33. These data columns were combined into a single dataset for subsequent transformations. To normalize the dataset and minimize the influence of extreme values, a log2 transformation was applied. A constant value of 1 was added to all area values prior to transformation to prevent undefined logarithmic calculations at zero. The log2-transformed dataset provides a normalized view of area values suitable for comparative analysis. The log2-transformed area values were visualized using a heatmap. This graphical representation highlights patterns and differences between control and sample groups. A heatmap was generated to visualize Z-score normalized and log2-transformed gene expression data using Python Script. For proteome gene expression visualization, log values of ≥1 were considered upregulated, while values < -0.5 were considered downregulated. For metabolome analysis, log values ≥1 indicated upregulation, and values ≤ -1 indicated downregulation. Data was pre-processed to include only relevant gene symbols and samples. Heatmap styling was applied with the Seaborn library (Sial et al., 2021).

### 2.7 Machine Learning for Biomarker Identification

#### 2.7.1 Data Preparation

There are received proteomic and metabolomics datasets containing control and sample groups’ expression abundances (Supplementary Information 1 and 2). Each marker was defined in terms of the expression of proteins and metabolites, and each of the samples was divided into two categories: Control 0 and Sample 1. The data required processing activities such as normalizing and scaling so that distribution would be the same throughout the data.

#### 2.7.2 Feature Selection and Data Splitting

To enhance the model and decrease dimension size, criteria features were chosen in accordance to biological and statistical relevance. The dataset was divided into a training set, which comprised 70% of the data, and a testing set, which comprised the other 30%. The division was done in a stratified manner to retain the class distribution.

#### 2.7.3 Random Forest Classification

The scikit-learn framework was used to develop a Random Forest classifier. In order to increase predictive power, this model was first fitted to the training dataset using the default hyperparameters, which comprised decision trees and a bootstrapping aggregation technique. In order to categorize the samples that were reserved for testing, the model fitting process involved feature generation, decision tree construction, and majority voting.

#### 2.7.4 Model Evaluation

Using the test set, the metrics that were defined above were calculated for the performance of the model.

##### 2.7.4.1 ROC Calculation

For this metric, the area under the receiver operating characteristic curve (AUC-ROC) was calculated using the function roc_auc_score. Discrimination was more accurate when higher AUC values were achieved in comparison to the control group and sample group.

##### 2.7.4.2 Sensitivity and Specificity

A confusion matrix was instrumental in extracting True Positive (TP), False Positive (FP), True Negative (TN), and False Negative (FN) values. To see how well the model was able to correctly classify the sample and control groups, sensitivity (TP / (TP + FN)) and specificity (TN / (TN + FP)) measures were computed.

##### 2.7.4.3 Probability Estimation

The predict_proba command was employed to provide class probability estimates, which were used for classification decisions. To test the stability of the model, bootstrapping methods were employed to test the statistical significance of the AUC-ROC metrics. The Random Forest model’s feature importance scores were used to analyze the effect of different features of the model on the classification metrics.

##### 2.7.4.4 Performance Evaluation and Statistical Validation

The following was the interpretation of AUC values: < 0.5 (poorer than random categorization) >0.9 (great), 0.8-0.9 (excellent), 0.7-0.8 (good), and 0.5-0.7 (poor discrimination). For the identification of proteomic and metabolomic biomarkers with strong diagnostic potential, our technique guarantees reliable and repeatable AUC estimations. AUC > 0.7 biomarkers were found and taken into consideration for further validation in separate cohorts.

##### 2.7.4.5 Software and Tools

Python (Version 3.11.6) was used for all studies, including NumPy (Harris et al., 2020) for numerical calculations and Pandas for data management. Scikit-learn was used for machine learning and statistical studies, with confusion_matrix being used for sensitivity and specificity evaluations and roc_curve and auc for ROC and AUC computations (Simfukwe et al., 2024). CSV file handling was used to store the processed results in a structured way (gene_metrics_final.csv). For the discovery of biomarkers in proteomic and metabolomic datasets, our integrated technique guaranteed reliable and effective AUC, sensitivity, and specificity computations.

### 2.8 Microarray analysis

The GEO database provided the gene expression data (GSE48150). Affymetrix microarrays were used for RNA extraction and hybridization after tooth germs from molars, incisors, and canines at the cap stage were removed from 12-week-old human embryonic oral cavities (Hu et al., 2013). The Robust Multi-array Average (RMA) technique in R was used to analyze and standardize the raw data. (Ritchie et al., 2015) used the limma package in R (version 3.5.2) to assess differentially expressed genes, adopting a log2 fold change (Log2FC) criterion of >1 for upregulation and <-1 for downregulation. To evaluate statistical significance, a p-value threshold of less than 0.05 was used.

### 2.9 Genes Panel

The metabolite-associated gene panel was curated using the OMIM database, focusing on the top eight pathways identified from upregulated and downregulated metabolites, as well as metabolites unique to controls and patients. Pathway-specific keywords were employed in the OMIM database to construct this panel. Proteome analysis, based on a significant list of upregulated and downregulated genes, was also utilized for gene panel preparation. Additionally, the differential expression genes (DEGs) panel and whole exome sequencing (WES) gene panel were developed in our previous work(Ranjan et al., 2024). The DEGs panel was derived from the GEO dataset ID GSE56486, as reported in the study by (Ranjan et al., 2025). The dual DEGs panel was obtained from microarray analysis of the GEO dataset GSE48150.

### 2.10 Pathway, Gene Ontology, Diseases and Network analysis

Enrichment analysis of upregulated, downregulated, and unique metabolites present in controls and patients was performed using MetaboAnalyst 6.0 (Pang et al., 2024). In parallel, enrichment analysis of upregulated and downregulated genes from proteomics data was conducted using Enrichr (Evangelista et al., 2023). In MetaboAnalyst 6.0, a list of metabolites with corresponding fold-change values was input for analysis. The Metabolite Set Enrichment Analysis (MSEA) module was utilized to identify enriched pathways, drawing from curated databases such as KEGG (Kanehisa, 2002) and HMDB (Wishart et al., 2022).

For network-based analysis, Metscape (Karnovsky et al., 2012)), a plugin for Cytoscape 2.8 (Smoot et al., 2011), was used to construct metabolic interaction networks. Metabolites and their associated genes, derived from sources such as the OMIM database (Amberger et al., 2015), proteomics data, differentially expressed genes (DEGs), whole exome sequencing (WES) panels, and the integrated gene panel, were analyzed using Enrichr for gene enrichment and pathway interactions to explore network dynamics and identify critical pathways (KEGG) , Gene ontology ( Biological Processes, Molecular Functions, and Cellular Components). This integrated approach facilitated a comprehensive analysis of metabolites and genes, revealing critical pathways and biological processes contributing to the dataset’s underlying biological mechanisms. Network analysis of proteome data, the integrated gene panel, and dual-expression genes was performed using STRING (Szklarczyk et al., 2019) with a default score of 0.4, followed by visualization in Cytoscape.

## 3. Result

### 3.1 CTA patient’s clinical features

Three patients diagnosed with CTA underwent serum-based metabolomics and proteomics analyses. Orthopantomograms (OPGs) confirmed oligodontia, characterized by the absence of multiple permanent teeth, including molars, premolars, and incisors. Patient Den 31 showed missing upper and lower premolars and lower incisors, Den 32 lacked upper lateral incisors, premolars, and lower molars, while Den 33 exhibited extensive tooth loss (Figure1).

### 3.2 Metabolomics and Proteomics profile

The comparative analysis of metabolomics and proteomics profiles between control and patient samples reveals significant variations, highlighting potential biomarkers for disease diagnosis and mechanistic insights. In metabolomics profiles, the chromatogram for control samples (Figure 2) displays distinct peaks representing various metabolites, with retention times along the x-axis and intensity on the y-axis. In contrast, the patient sample chromatogram (Figure 2) shows altered peak patterns, indicating significant changes in metabolite levels in the patient group. Similarly, the proteomic chromatogram for control samples (Figure 2) features peaks corresponding to different proteins, with retention time and intensity as axes. The patient sample chromatogram exhibits notable variations in peak patterns, suggesting differential protein expression between the two groups. These differences in both metabolomic and proteomic profiles underscore the altered biochemical and molecular landscapes in patients, providing a foundation for identifying potential biomarkers and understanding the underlying disease mechanisms.

**Figure 1:**
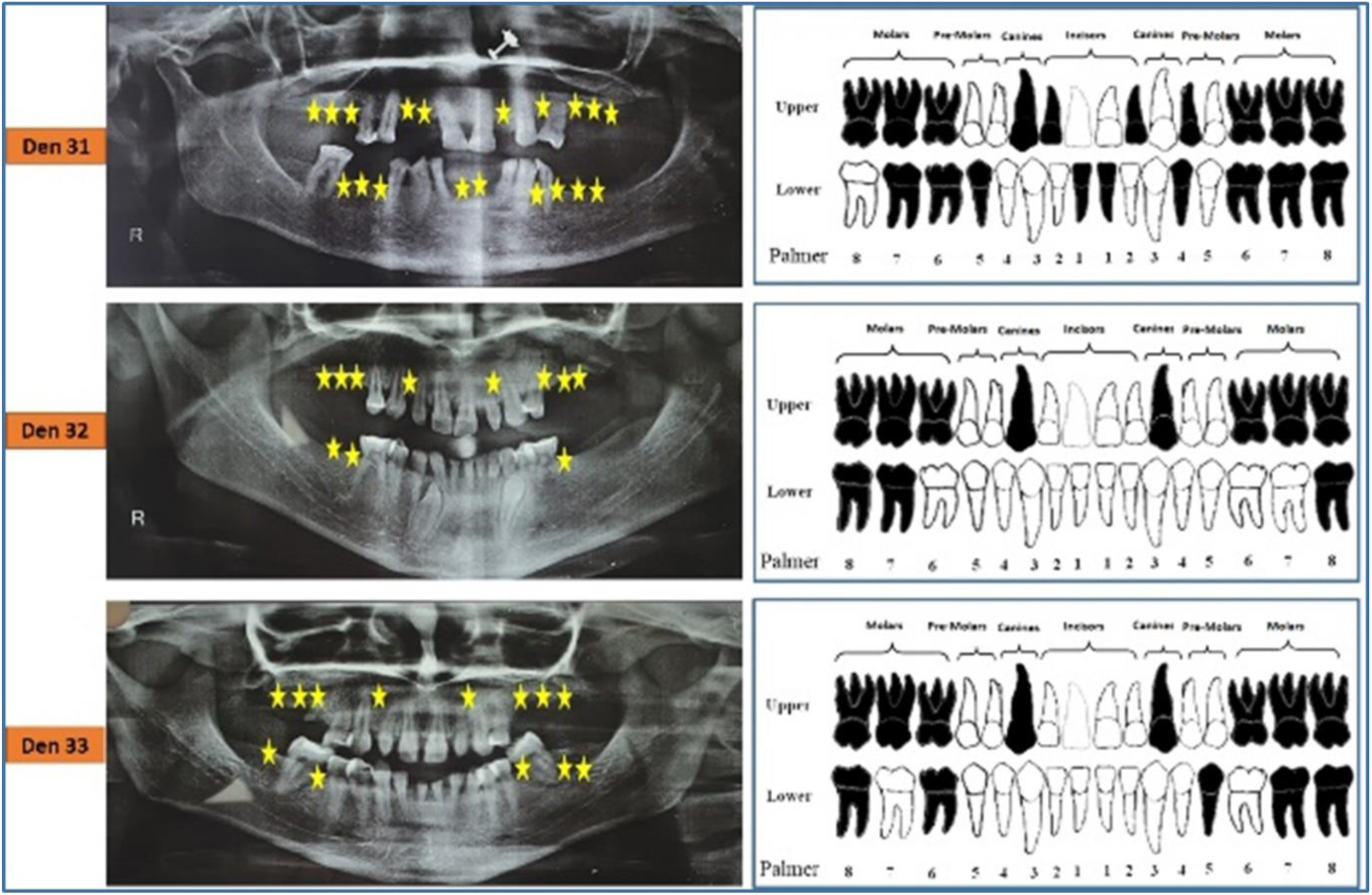
Orthopantomogram images of patients with CTA illustrating missing teeth. This figure illustrates panoramic dental X-rays from three different patients labeled as Den 31, Den 32, and Den 33. Each X-ray is accompanied by a corresponding dental chart that indicates missing teeth. The missing teeth are marked with yellow stars on the X-rays and are shaded black on the dental charts (Palmer Nomenclature). The charts are divided into upper and lower sections, categorizing teeth as molars, pre-molars, canines, and incisors. The Palmer notation system is employed to identify the teeth accurately.

**Figure 2:**
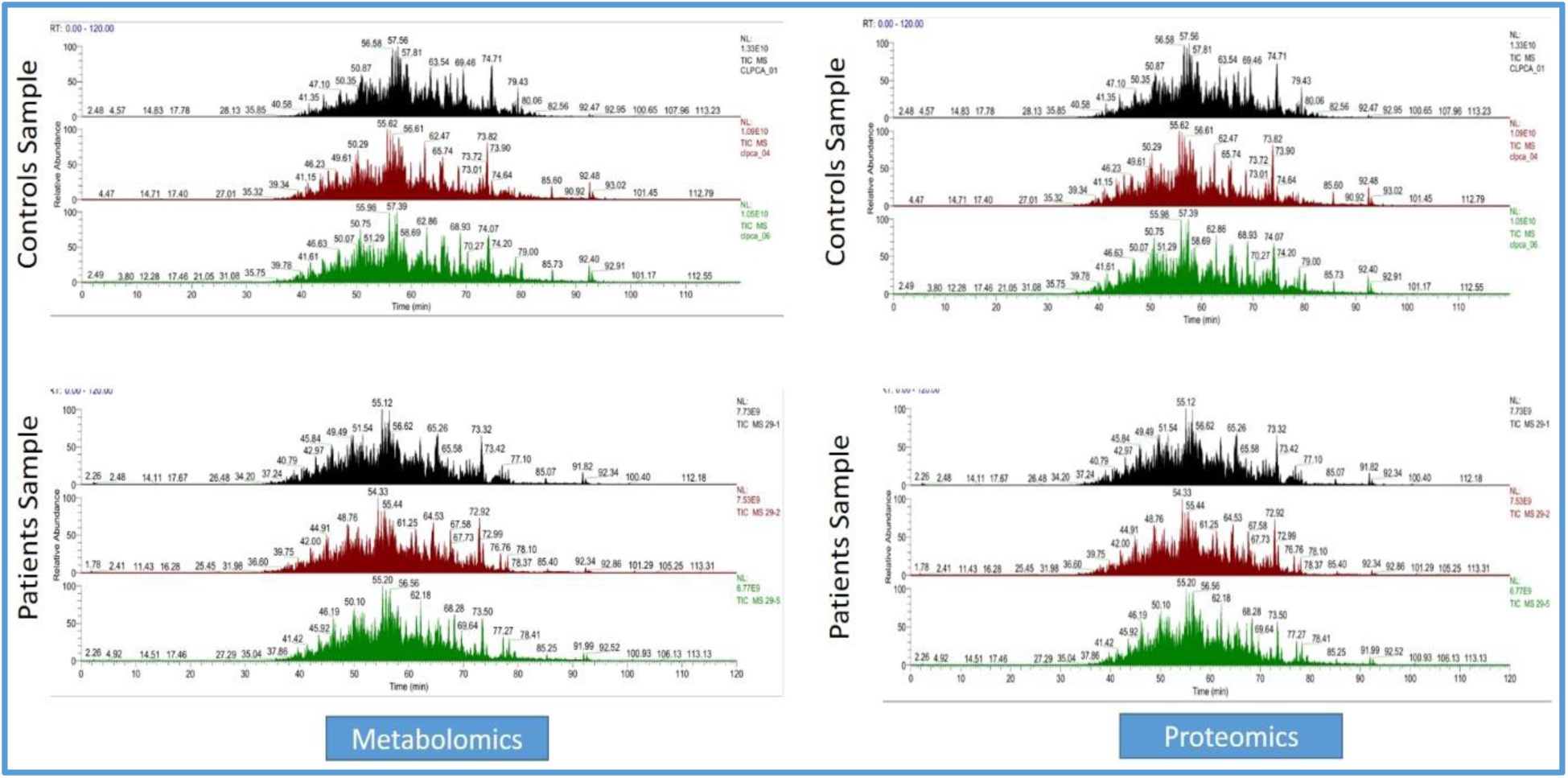
The figure presents the Total Ion Chromatogram (TIC) analysis comparing control and patient samples. The TICs display the retention time (x-axis) versus the intensity of detected ions (y-axis) for both metabolomics and proteomics analysis. A. Control Samples: The top panels show the TICs for control samples. The left panel represents metabolomics data, while the right panel represents proteomics data. Each chromatogram includes multiple experimental conditions or replicates, indicated by different colors. B. Patient Samples: The bottom panels depict the TICs for patient samples. Similarly, the left panel shows metabolomics data, and the right panel shows proteomics data, with different colors representing various experimental conditions or replicates. Peaks in the chromatograms indicate the presence and abundance of various metabolites or proteins. Differences in peak patterns and intensities between control and patient samples highlight potential biomarkers or disease-related changes.

The metabolite profiles revealed significant differences between CTA patients and control subjects, emphasizing the dysregulated metabolic landscape associated with the condition. Significance analysis identified key metabolites in Table1 and Figure 3. Metabolites with positive log2 fold changes were upregulated in CTA patients, while those with negative values were downregulated, highlighting a clear pattern of metabolic alterations. Principal Component Analysis (PCA) demonstrated distinct separation between control and patient groups, reflecting robust metabolic dysregulation in CTA patients Figure 3. This separation underscores the potential of metabolite profiling in distinguishing between these groups. A heatmap of log2-transformed metabolite area values further illustrated differential expression across the samples Figure 3. Variations in expression intensities highlighted significant metabolic differences, with certain metabolites showing markedly higher or lower levels in CTA patients compared to controls. Overall, the analysis underscores the extent of metabolite dysregulation in CTA patients, providing valuable insights into potential biomarkers and pathways implicated in the condition.

**Figure 3:**
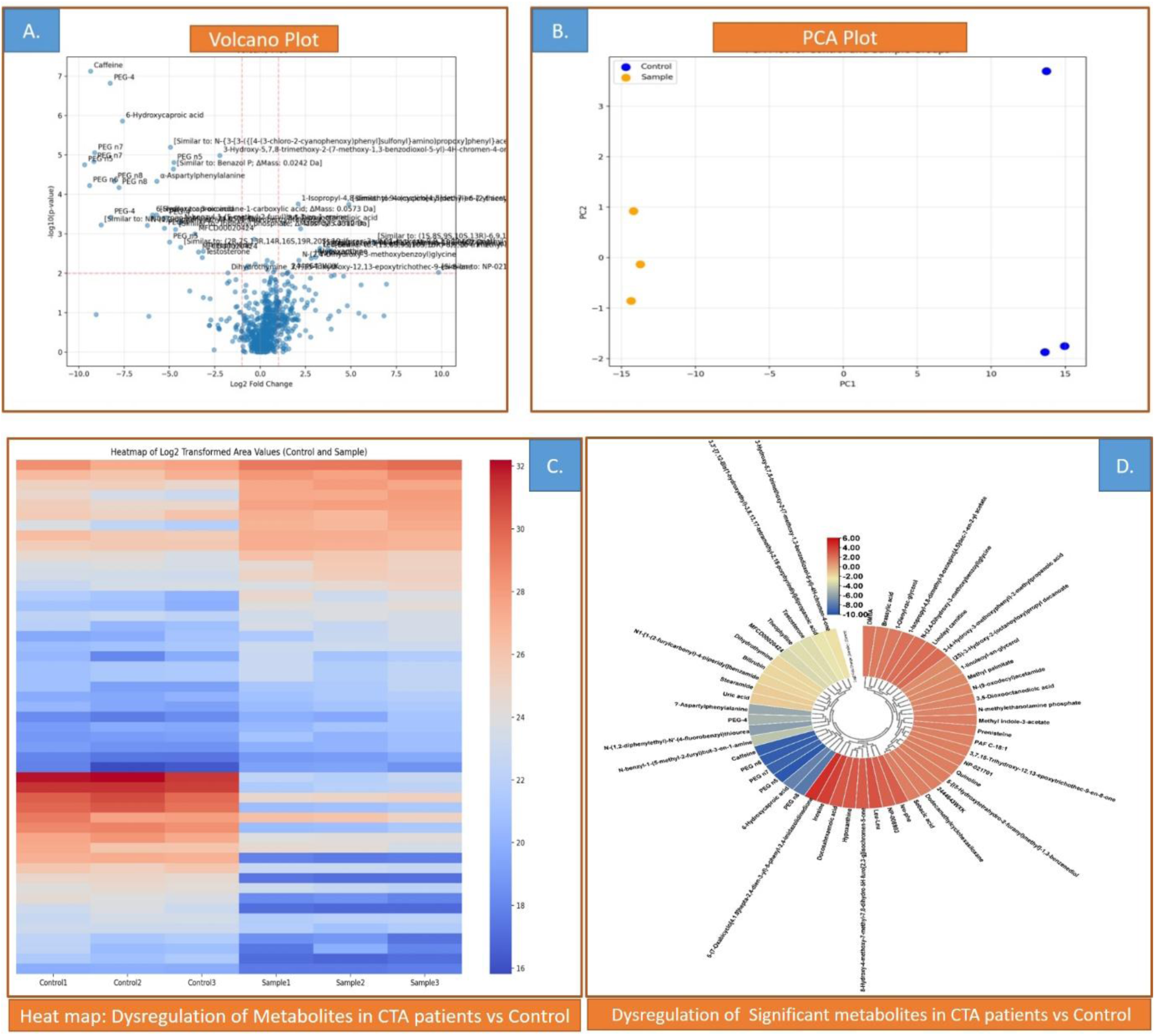
Metabolite Dysregulation in CTA: Comprehensive Analysis (A) Volcano Plot: The plot depicts the log2 fold change on the x-axis and the -log10 p-value on the y-axis, identifying significantly dysregulated metabolites in CTA patients compared to controls. Metabolites with significant upregulation in CTA patients are marked on the right, while those with significant downregulation are marked on the left. (B) PCA Plot: Principal Component Analysis (PCA) plot demonstrates clear separation between control (blue dots) and CTA sample groups (orange dots). The clustering pattern highlights distinct metabolic profiles between the groups. (C) Heatmap: A heatmap showing log2- transformed metabolite area values for both control and CTA sample groups. The color gradient (blue to red) represents the range of metabolite expression levels, with blue indicating lower expression and red indicating higher expression. (D) Circular Bar Heatplot: A circular bar plot visualizes the dysregulation of metabolites in CTA patients relative to controls. The fold change is depicted with a color gradient from blue (downregulation) to red (upregulation), highlighting the degree of metabolic alteration in CTA patients.

### 3.3 Dysregulated Metabolites

Metabolite analysis revealed significant dysregulation in comparison to the known ecto-mesodermal metabolite panel (Known_MB_Panel) Figure 4. A total of 591 metabolites (92.1%) were exclusive to the Known ecto-mesodermal Metabolite Panel, while 28 metabolites (4.4%) were detected exclusively in upregulated samples (UP_MB), and 17 metabolites (2.7%) were identified exclusively in downregulated metabolites (Down_MB). Three metabolites, inosine, stearic acid, and Leu-Leu, were shared between Known_MB_Panel and UP_MB, whereas testosterone, benzoic acid, and uric acid were common between Known_MB_Panel and Down_MB Figure 4.

**Figure 4:**
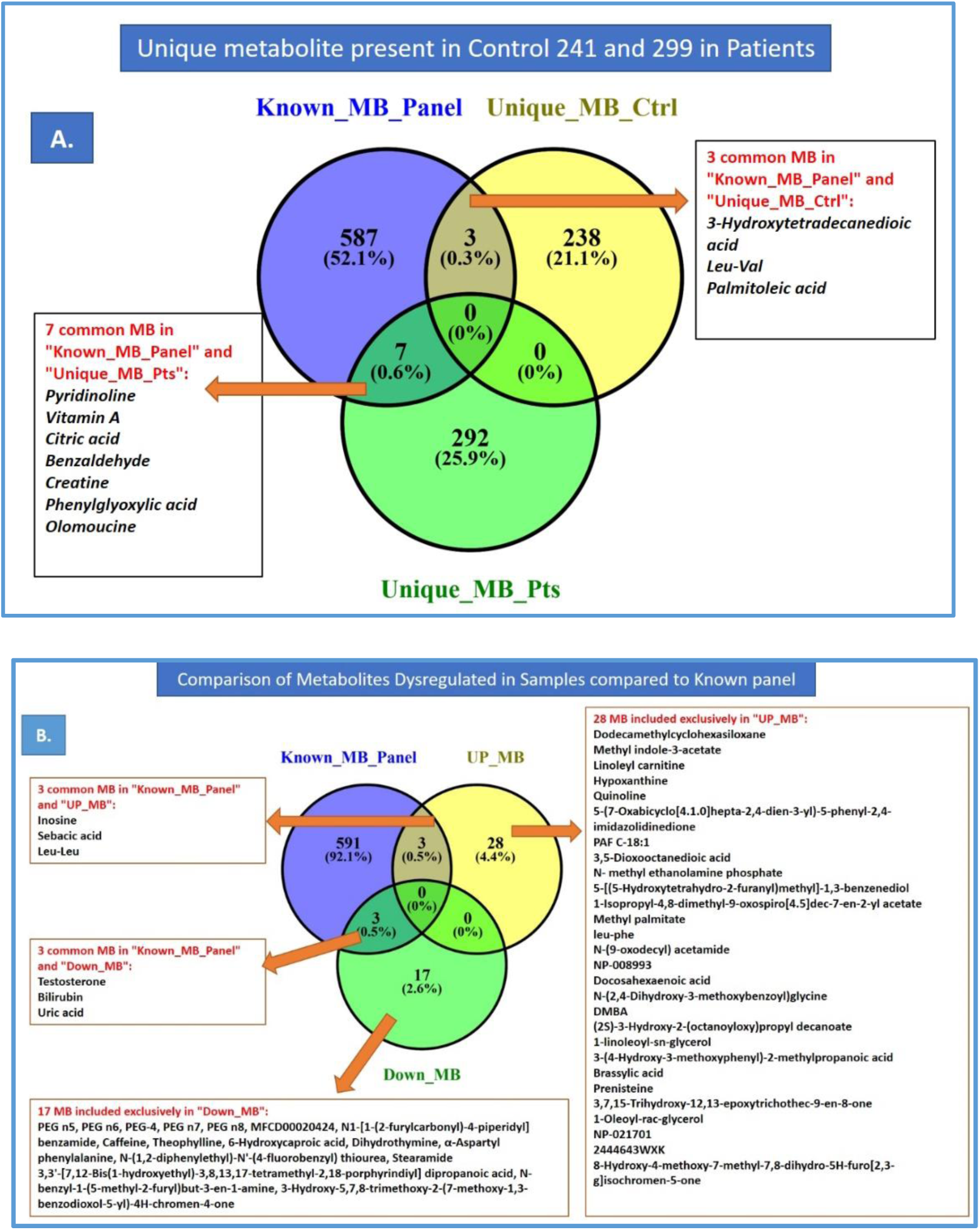
Distribution of Unique Metabolites and Dysregulated Metabolites Across Control and CTA Groups. A. The three sets represent Known Metabolites Panel (blue) with 587 unique metabolites (52.1%), Unique Metabolite present in control (yellow) with 238 unique metabolites (21.1%), and Unique metabolites present in only CTA group (green) with 292 unique metabolites (25.9%). The intersections indicate shared metabolites, with 7 common between Known Metabolites Panel and unique metabolites in CTA group (Pyridinolone, Vitamin A, Citric acid, Benzaldehyde, Creatine, Phenylglyoxylic acid, and Olomoucine) and 3 commons between Known Metabolites Panel and Unique Metabolites in control group (3-Hydroxytetradecanedioic acid, Leu- Val, and Palmitoleic acid). B. Venn diagram shows the overlap between Known Metabolites Panel (blue), UP regulated metabolites (green, upregulated metabolites), and Down regulated Metabolite (yellow, downregulated metabolites). Common metabolites include Inosine, Sebacic acid, and Leu-Leu between Known Metabolites Panel and UP metabolites, while Testosterone, Bilirubin, and Uric acid are shared between Known Metabolites Panel and down regulated Metabolites. Additionally, 17 metabolites are exclusively downregulated in down regulated Metabolites, including various PEG compounds and other significant molecules. This diagram highlights metabolites uniquely altered in the studied conditions.

The 28 metabolites unique to UP_MB included docosamethylencholestanone, methyl undecanoate, hexyl isovalerate, quinoline derivatives, and phosphatidylcholines, among others. Conversely, the 17 metabolites exclusive to Down_MB included PG 45, MG 45, PEG 4, phosphatidylcholine derivatives, and benzodioxole derivatives among others Figure 4. These findings indicate distinct metabolic shifts, with specific metabolites being upregulated or downregulated in the analyzed samples. The upregulated DHA metabolite has been associated with the Wnt signaling pathway and NF-κB signaling, both of which are involved in tooth development Figure 5.

**Figure 5.**
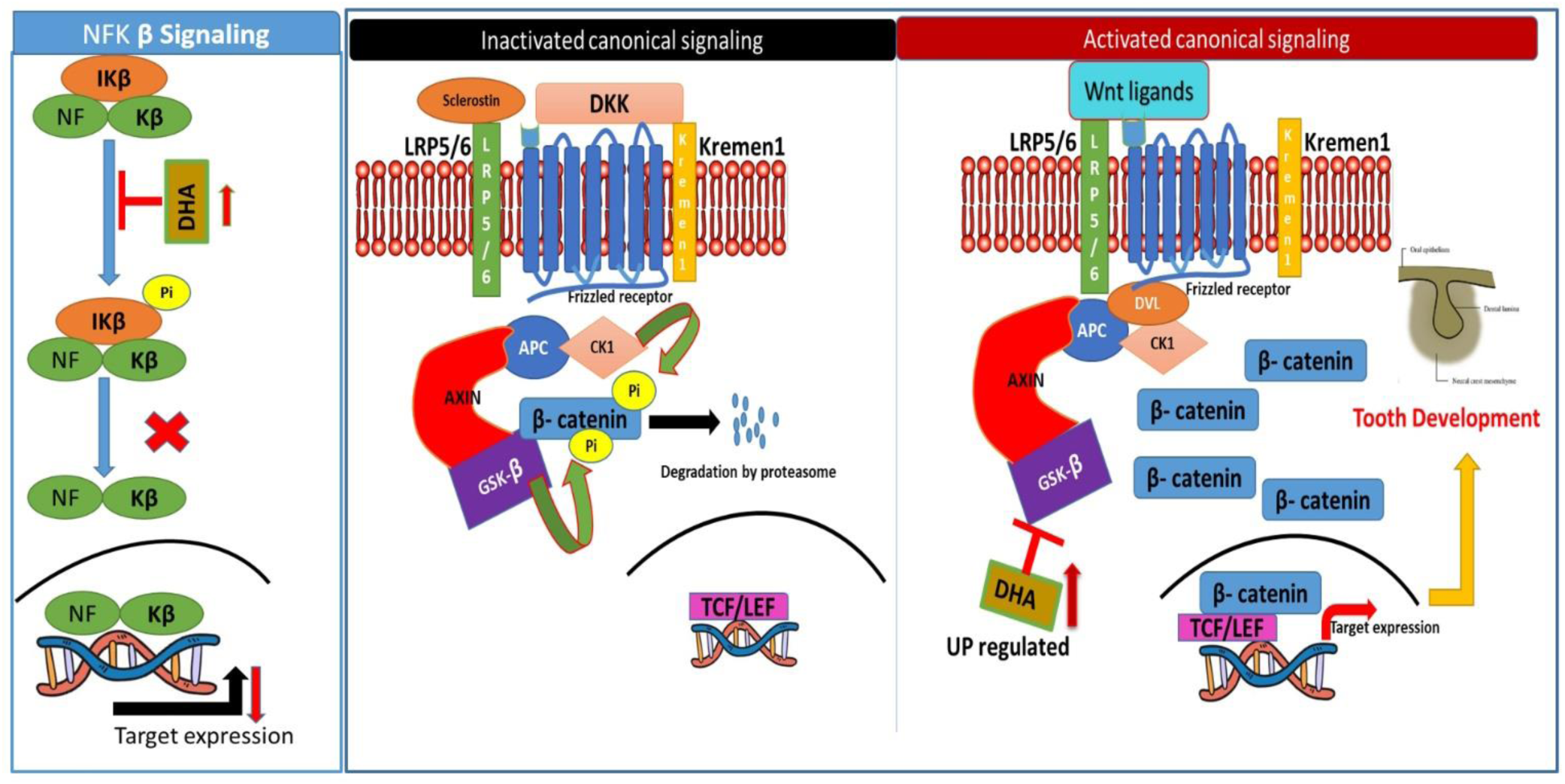
Effect of upregulated DHA on Wnt and NF-κB signaling pathways contributing to tooth development. The NF-κB signaling pathway is modulated by DHA, which inhibits the phosphorylation of IκB, preventing the release of NF-κB subunits for nuclear translocation. This regulation results in reduced target gene expression associated with inflammation. In parallel, DHA promotes activation of the canonical Wnt signaling pathway by suppressing inhibitors such as DKK and enhancing β-catenin stabilization. This leads to the activation of TCF/LEF transcription factors, thereby increasing target gene expression associated with cell proliferation and differentiation. These coordinated actions of DHA highlight its role in promoting cellular mechanisms essential for tooth development.

### 3.4 Comparison of Unique Metabolites in Control and Patient Samples

The metabolite analysis revealed distinct distributions of unique metabolites between control and patient samples, highlighting significant metabolic alterations associated with the disease. Control samples contained 238 unique metabolites, accounting for 21.1% of the total identified metabolites, while patient samples exhibited 292 unique metabolites, representing 25.9% of the total (Figure 4). A known ecto-mesodermal metabolites panel included 587 metabolites(Ranjan et al., 2025), covering 52.1% of the total identified pool. Common metabolites were minimal, with only three—3-Hydroxytetradecanedioic acid, Leu-Val, and Palmitoleic acid—overlapped between the known metabolite panel and control samples, representing 0.3% of the total metabolites (Figure 4). Seven metabolites, including Pyridinolone, Vitamin A, Citric acid, Benzaldehyde, Creatine, Phenylglyoxylic acid, and Olomoucine, were shared between the known metabolite panel and patient samples, accounting for 0.6% of the total (Figure 4). These results underscore the significant metabolic distinctions between control and patient samples, with the identification of unique and overlapped metabolites offering insights into disease-associated pathways and potential biomarkers for diagnosis and therapy.

### 3.5 Pathway, Disease Insights and Metabolite-Metabolite network

Pathway analysis revealed distinct metabolic differences between unique metabolites in control and patient samples with congenital tooth agenesis (Figure 6; Supplementary Information Table1). Unique metabolites identified in patient samples were mapped to pathways, including lipid metabolism, vitamin metabolism, amino acid transport, inflammatory regulation, pluripotency/differentiation, energy metabolism, and hormonal pathways (Figure 6). Comparative analysis highlighted dysregulation in these pathways, indicating their potential involvement in the etiology of tooth agenesis. These findings are represented in **Figure 6** and summarized in Supplimentary Information Table1 and **Table -4**, which detail the metabolites unique to each pathway and their differential expression in controls versus patients.

**Figure 6:**
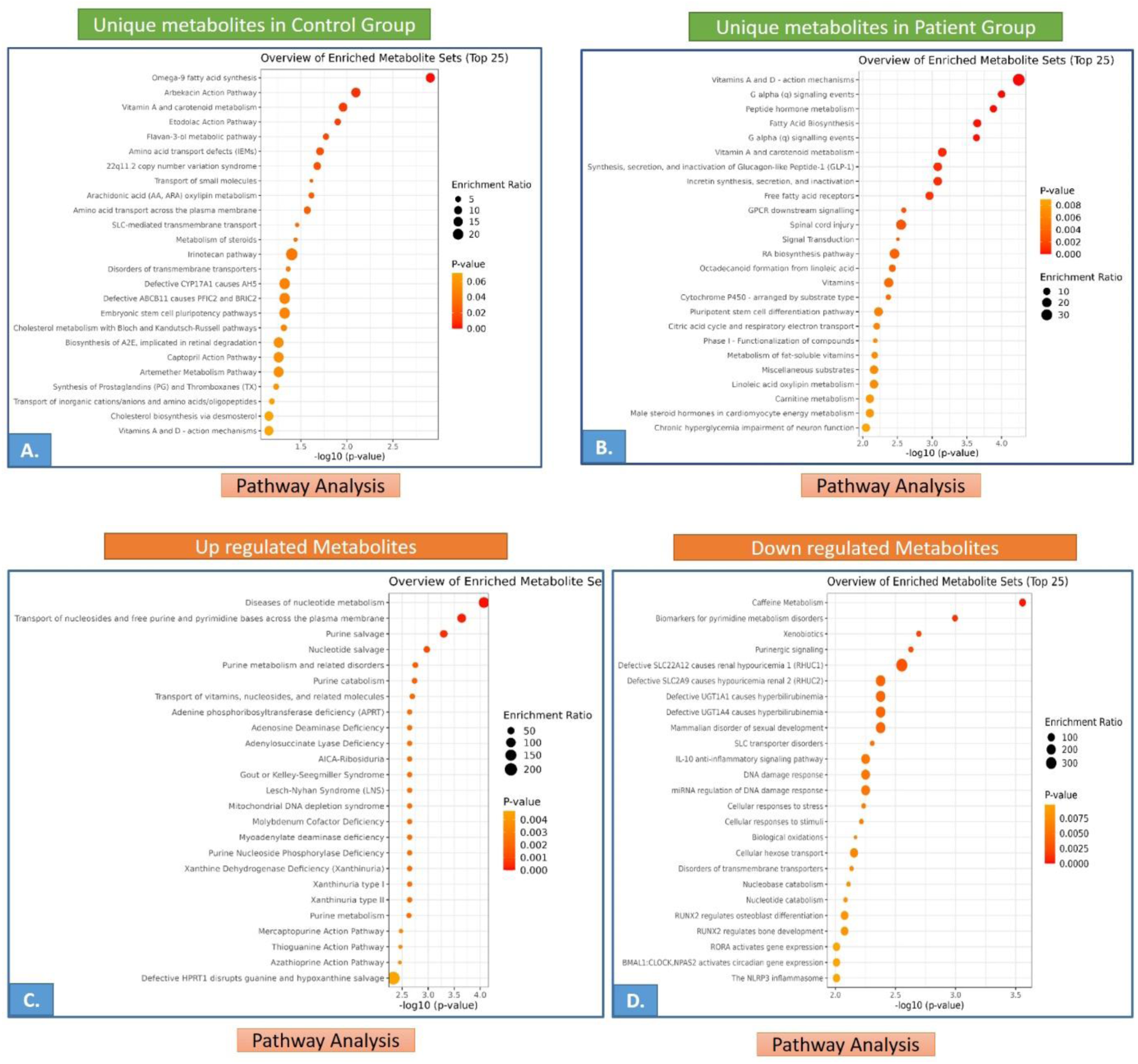
Pathway analysis of metabolites across control and patient groups is depicted in four panels. **A** shows the top 25 enriched pathways of unique metabolites in the control group, while **B** highlights the top 25 pathways of unique metabolites in the patient group. **C** presents the top 25 pathways enriched for upregulated metabolites, and **D** illustrates the top 25 pathways enriched for downregulated metabolites in the patient group. The x-axis represents the -log10 (p- value), the y-axis lists pathways, with dot size indicating the enrichment ratio and the color gradient representing p-value significance.

Upregulated metabolite pathways involved in congenital tooth agenesis include Nucleotide Metabolism and Transport and Purine Metabolism, which are essential for DNA and RNA synthesis and cellular energy metabolism, ensuring proper cellular function (**Figure 6**). Downregulated metabolite pathways include Caffeine Metabolism, Purinergic Signaling, Defective Transporters, IL-10 Anti-inflammatory Signaling, and DNA Damage Response and Cellular Stress, reflecting disruptions in cellular communication, transporter function, inflammatory responses, and DNA repair mechanisms, all of which could contribute to tooth agenesis (**Figure 6)**. The identification of the top 10 significant diseases associated with unique metabolites in the patient and control groups revealed a strong link with ectodermal and mesodermal disorders, as determined by the analysis of upregulated and downregulated metabolites. **(Table 3**. Several metabolites based on previous research literature showed potential associations with pathways relevant to CTA (Ranjan et al., 2025). Upregulated metabolites include 6-Hydroxycaproic acid and Stearamide (lipid metabolism), Dihydrothymine, Uric acid, and Inosine (purine metabolism), Docosahexaenoic acid (DHA) (NKB and Wnt signaling pathway), and Testosterone (hormonal signaling). Downregulated metabolites such as PEGs, bilirubin, and caffeine require further investigation. Detailed associations and pathway links are summarized in **Table 4**. MetScape analysis revealed metabolite-metabolite interactions among unique metabolites identified in control and patient samples, as well as dysregulated metabolites (**Figure 7)**. This analysis provides insights into how these metabolites interact with other metabolites presents in metscape database, shedding light on their potential roles and involvement in the molecular mechanisms underlying CTA (**Figure 7)**.

**Figure 7:**
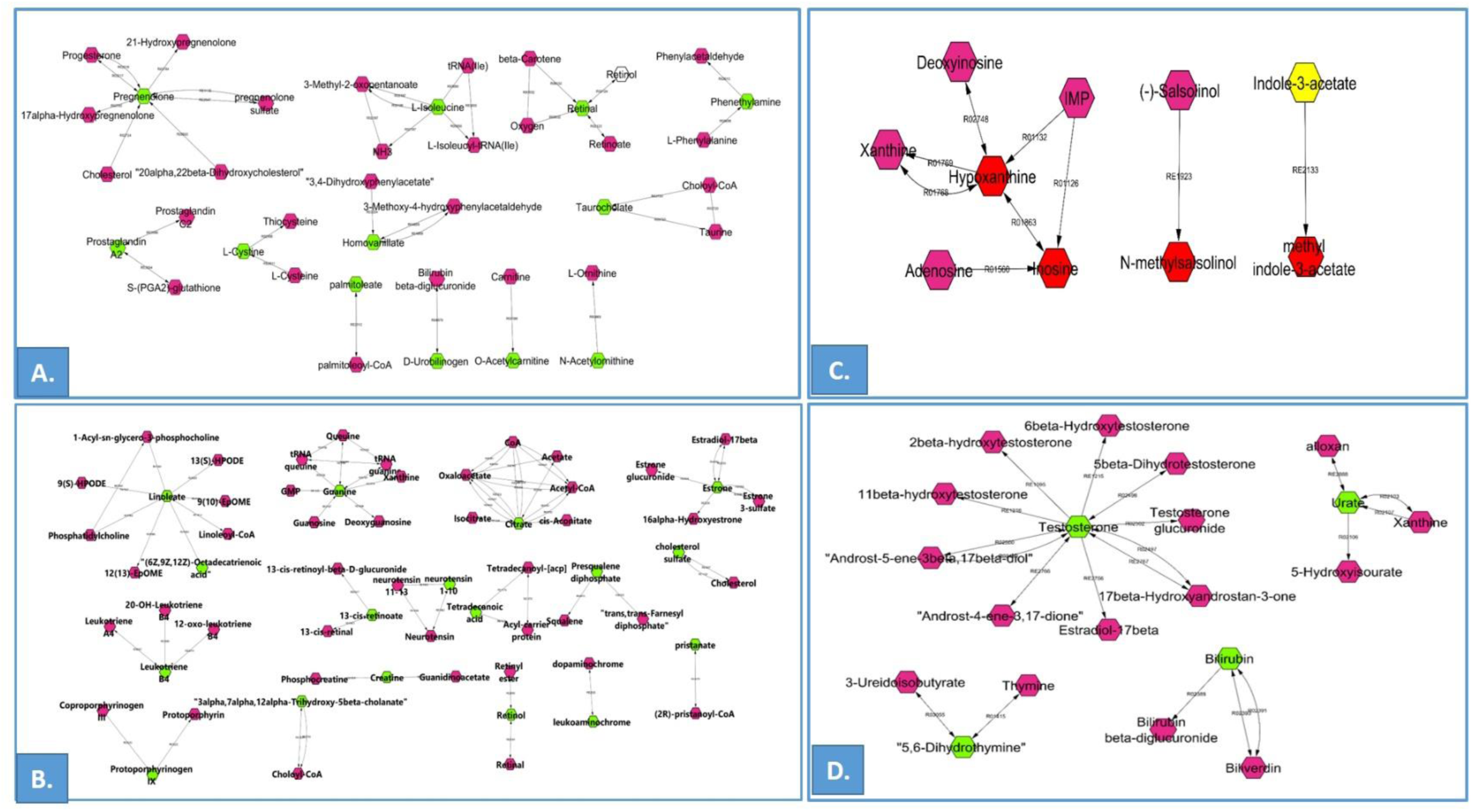
Metabolite Interaction Networks in Control and Patient Groups**. A:** Unique metabolite interaction network in the control group. Green nodes represent metabolites exclusively present in the control group, while other nodes indicate interacting metabolites. **B:** Unique metabolite interaction network in the patient group. Green nodes denote metabolites unique to the patient group, with other nodes representing their interacting metabolites. **C:** Upregulated metabolites identified through MetScape and network analysis. Red nodes signify upregulated metabolites, and other nodes represent their interacting metabolites. **D:** Downregulated metabolites identified through MetScape and network analysis. Green nodes indicate downregulated metabolites, with other nodes depicting their interacting metabolites.

**Table 1:**
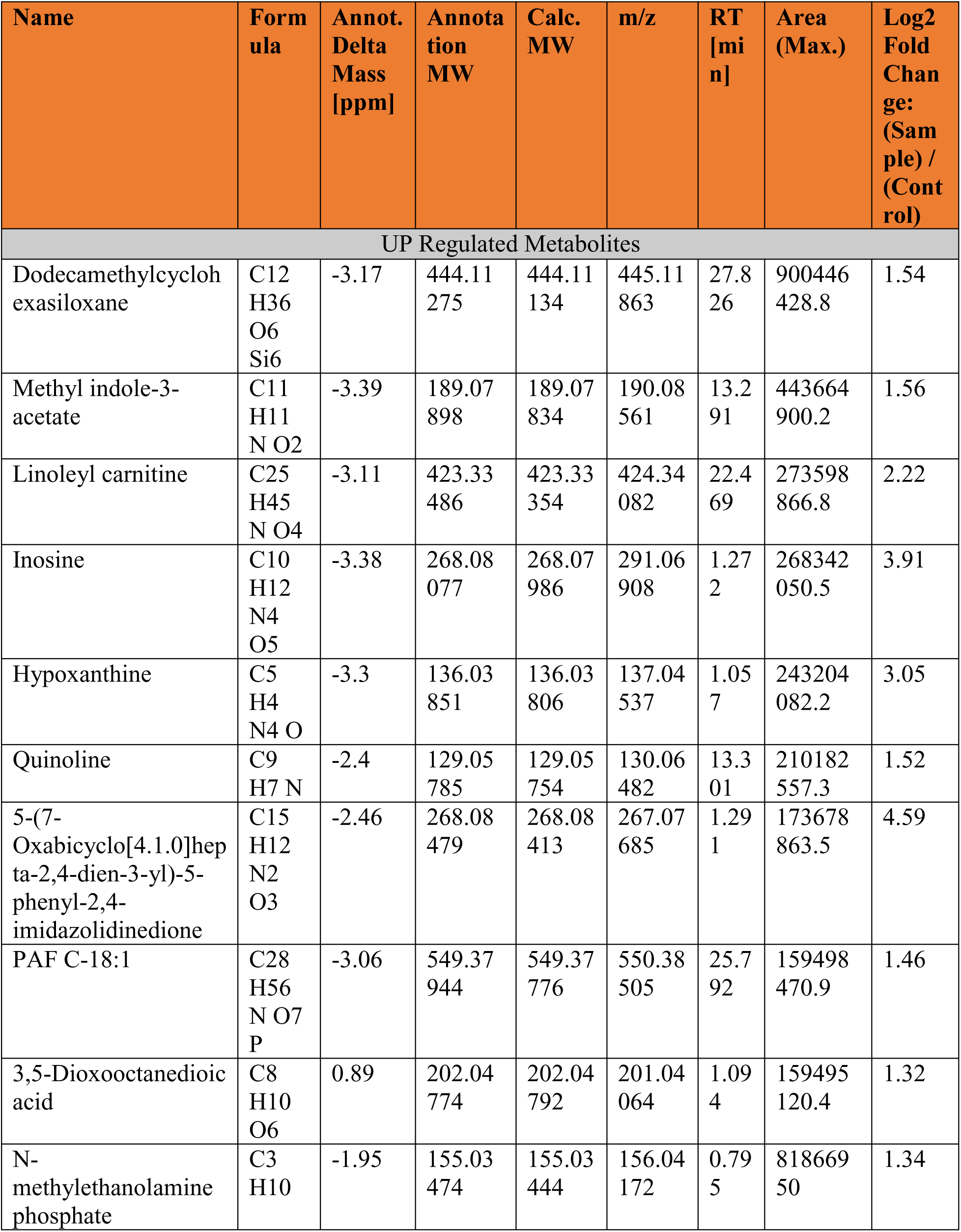
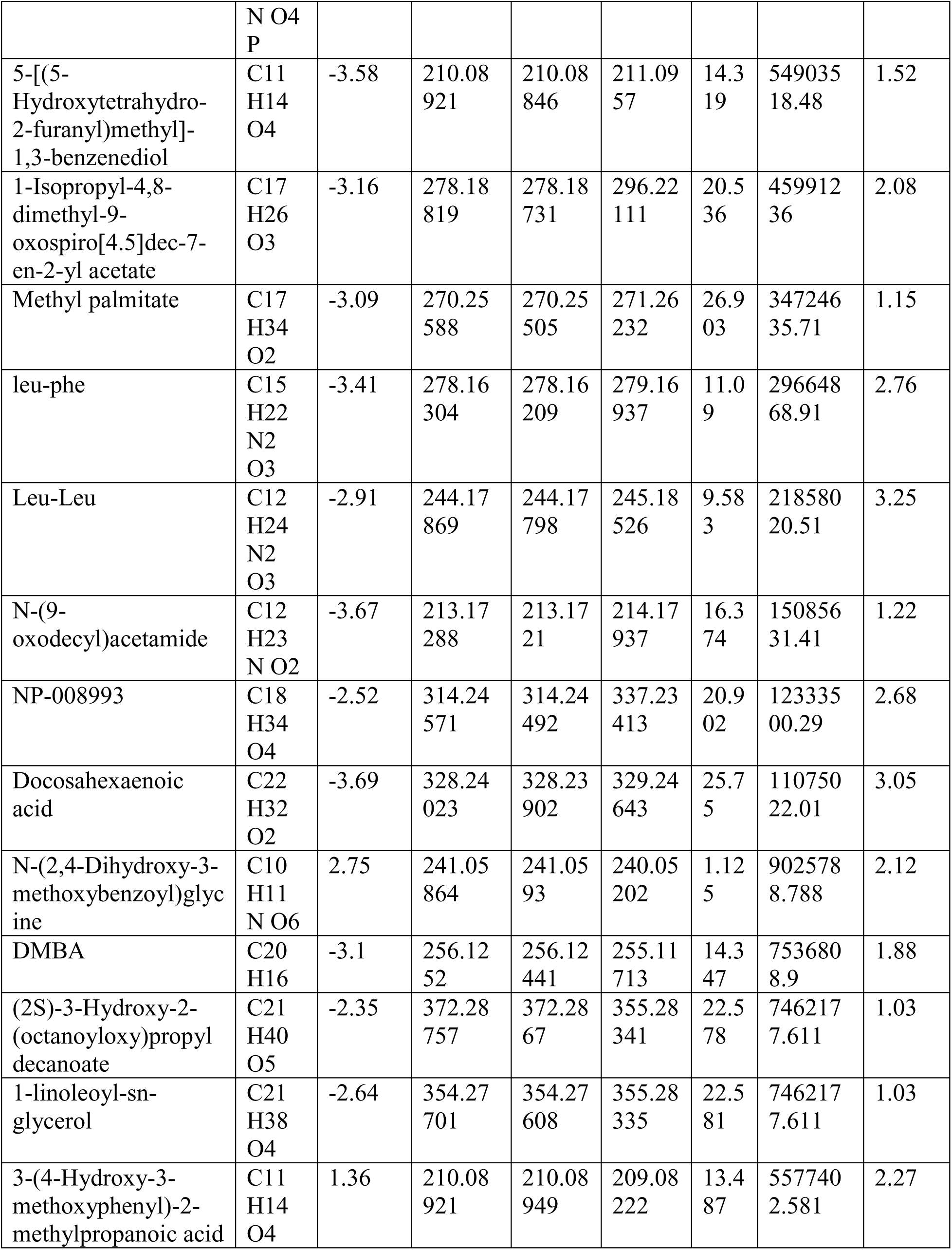
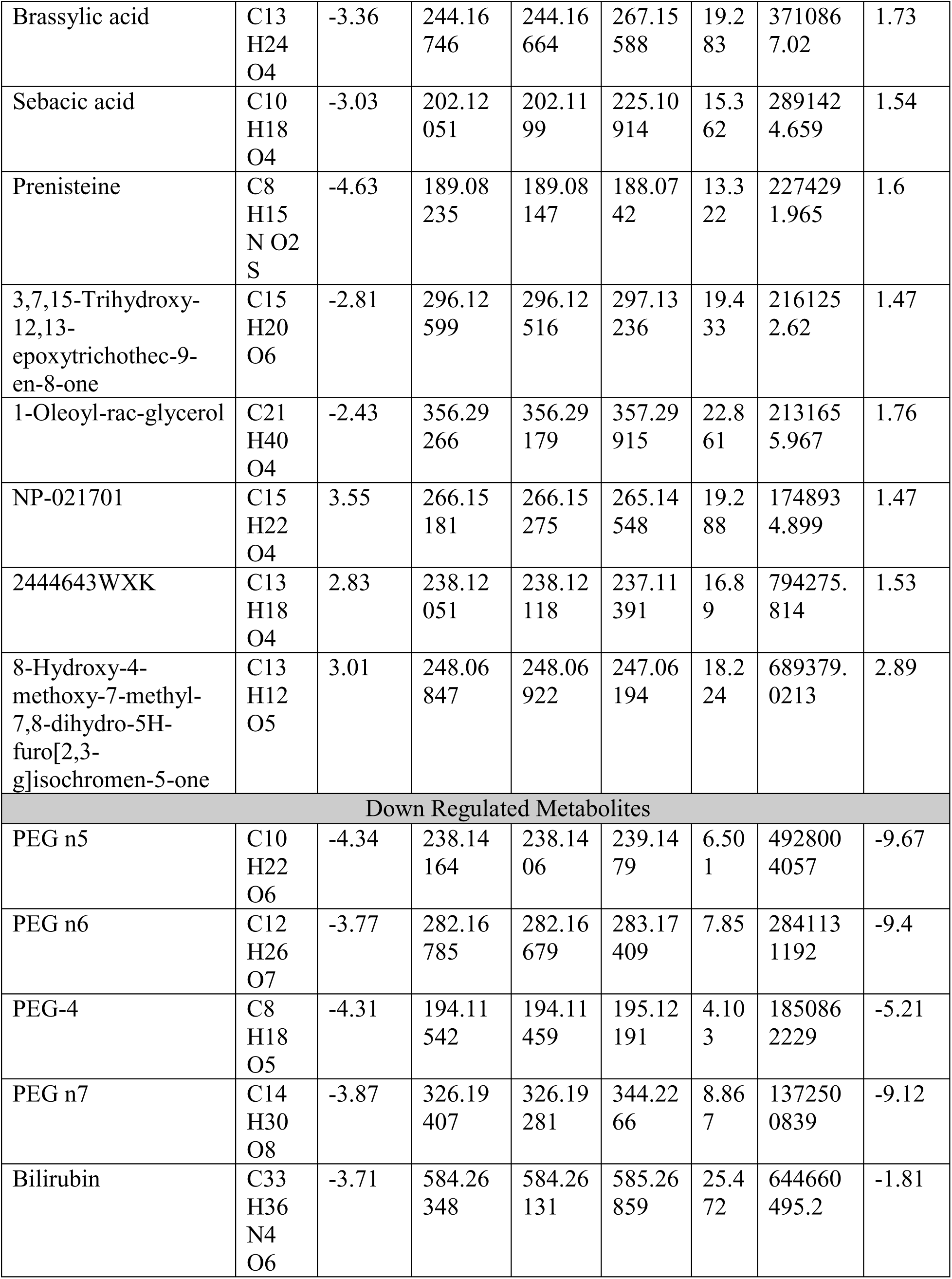
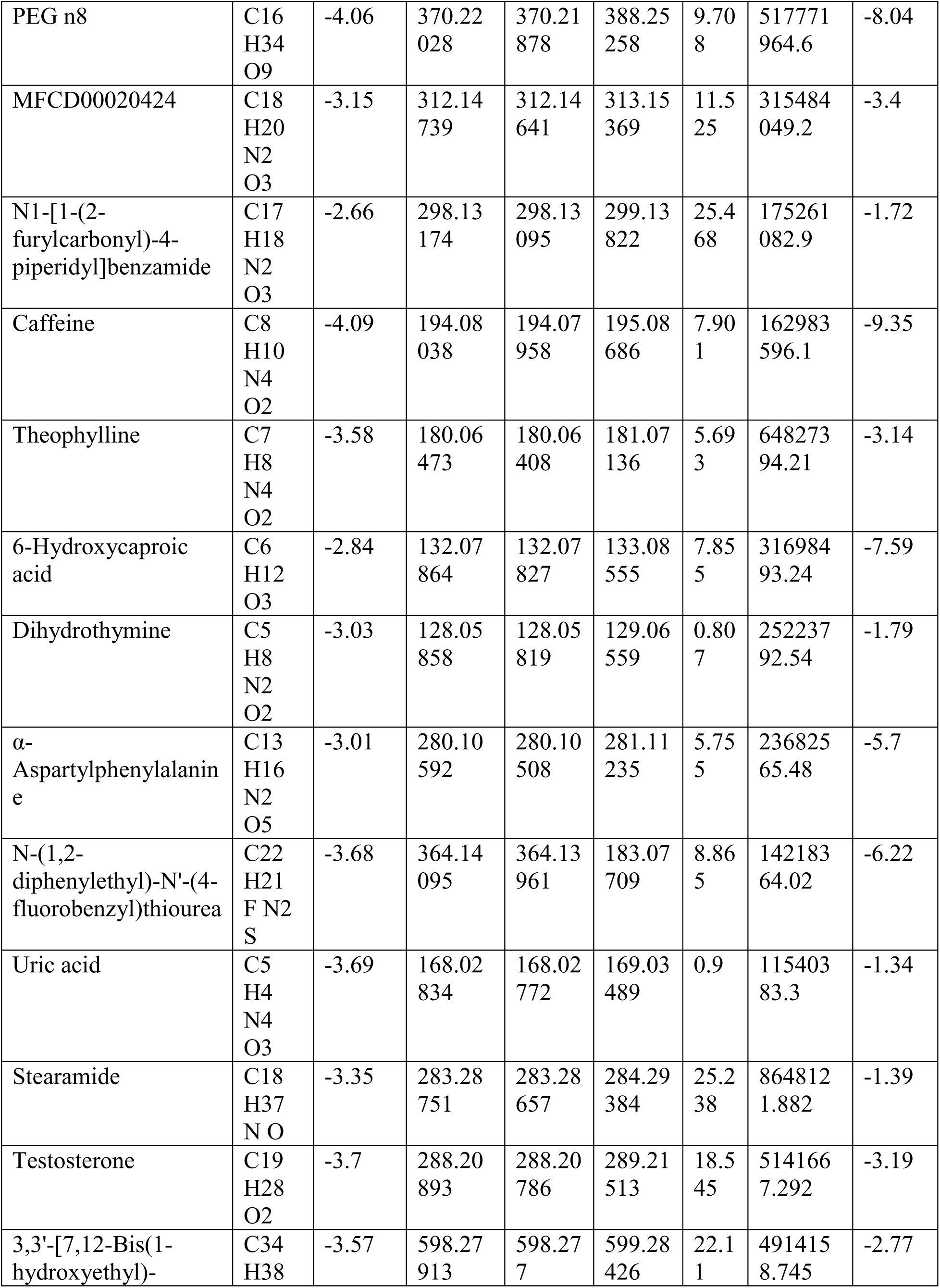
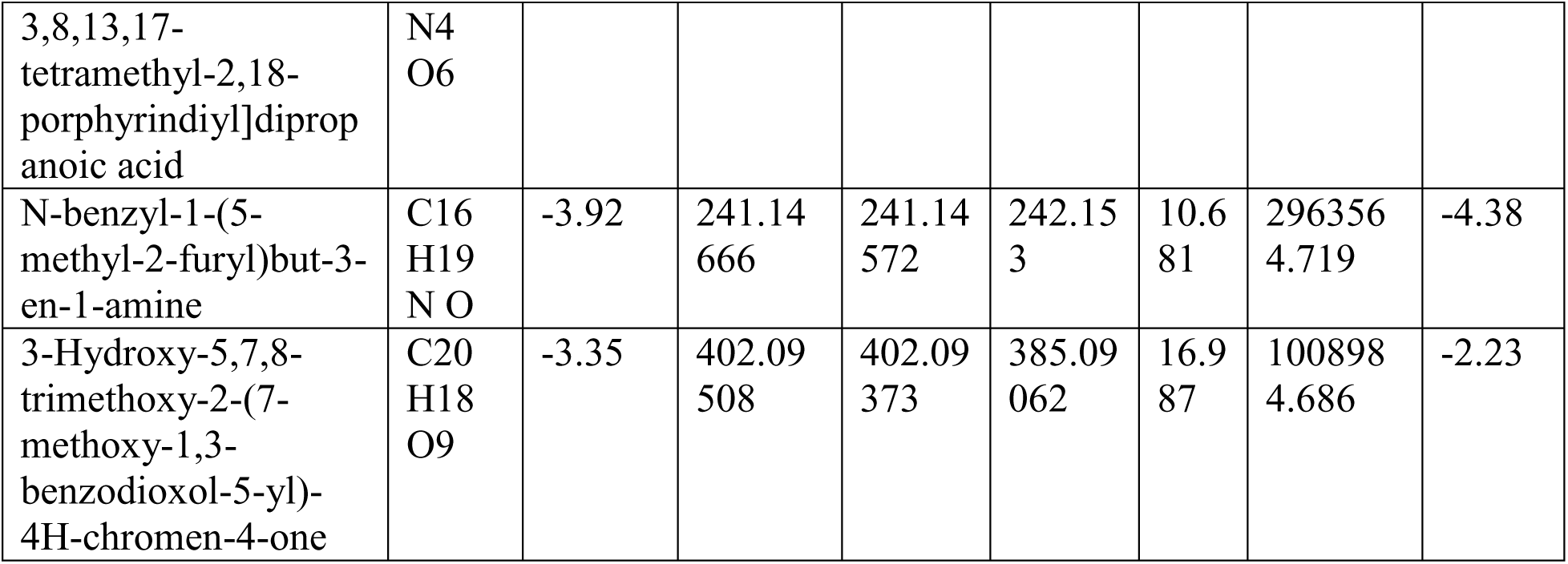
List of major metabolite compounds identified by UHPLC-HRAMS analysis through negative and positive ion showing chemical formula and annotation.

**Table 2:**
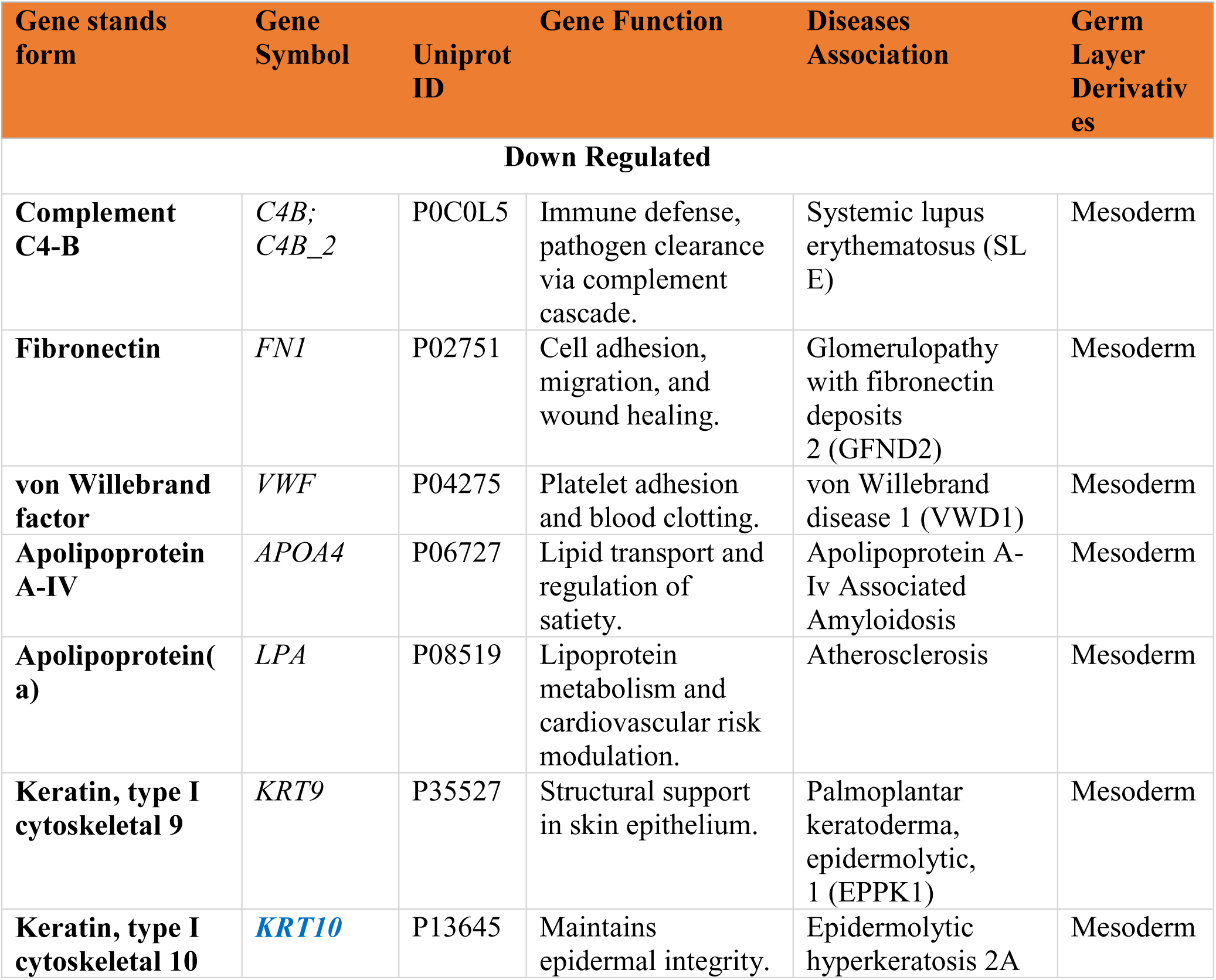
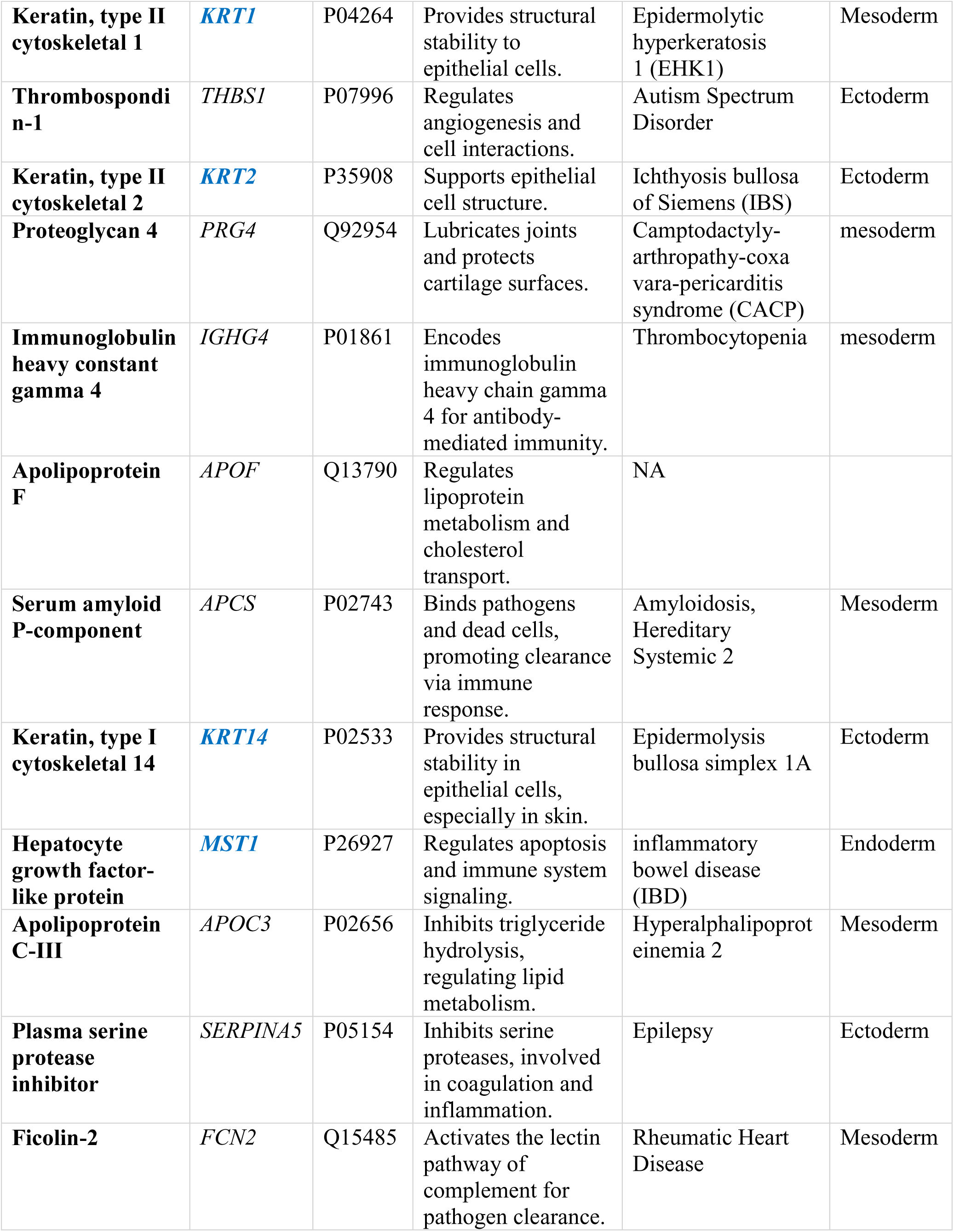
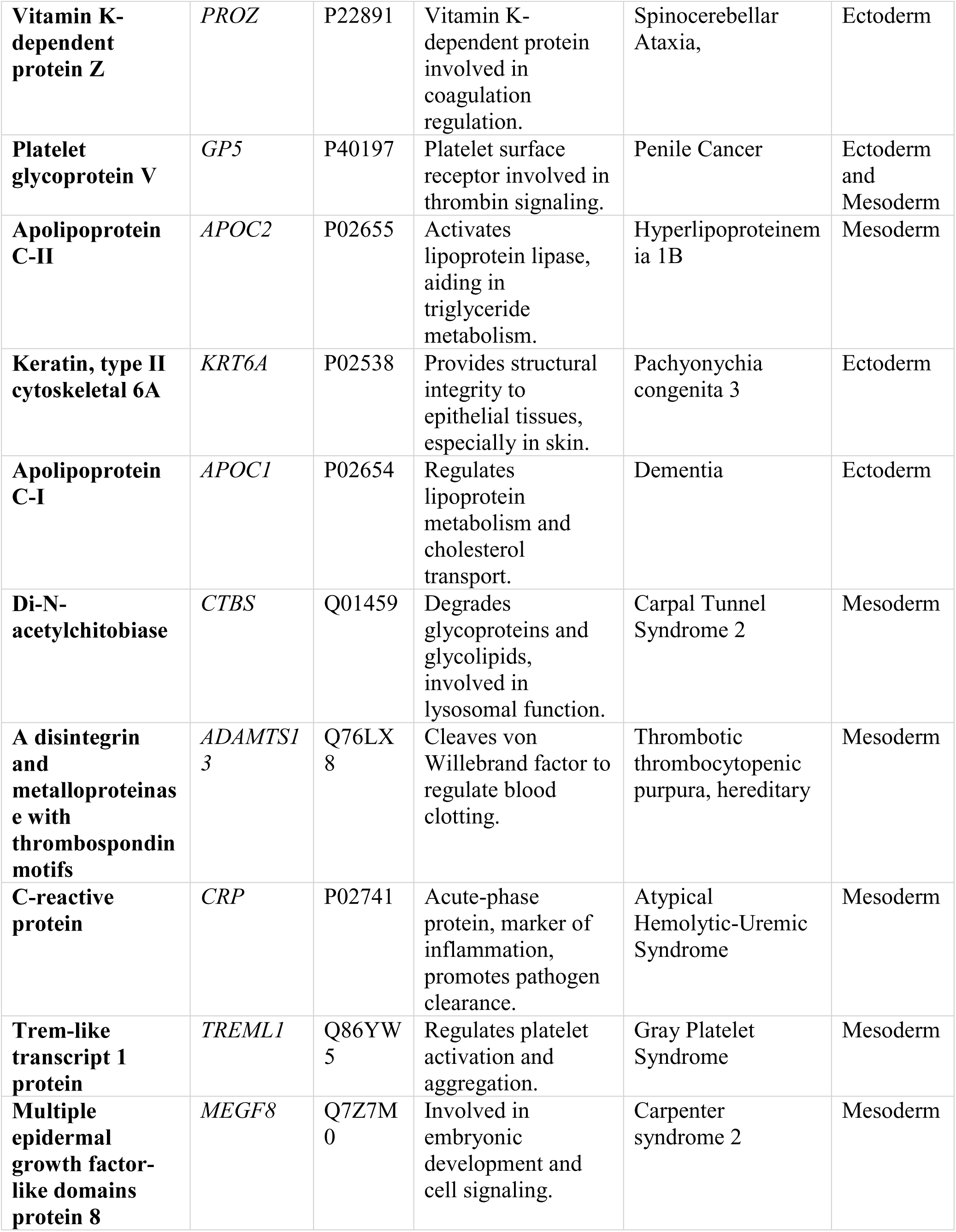
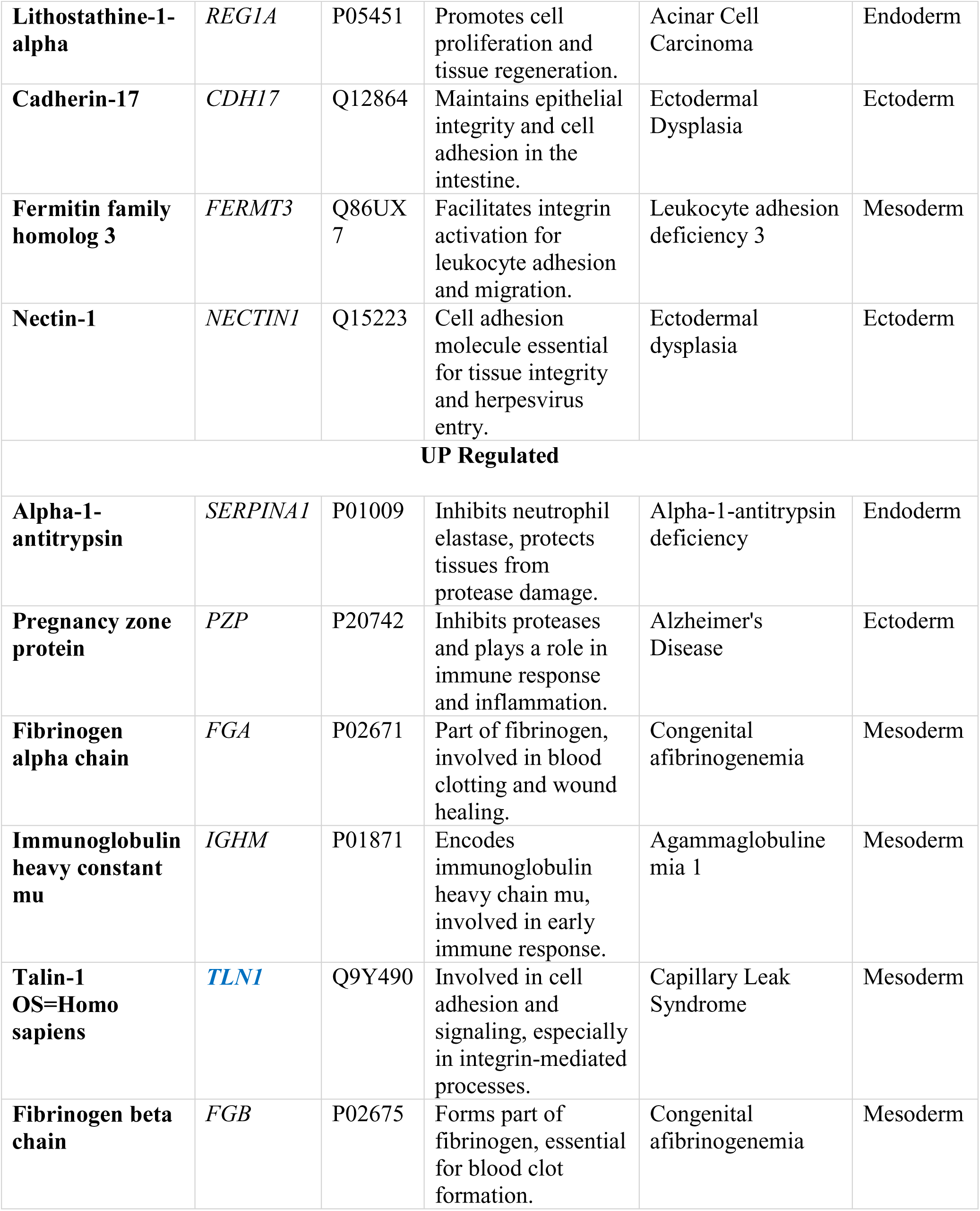
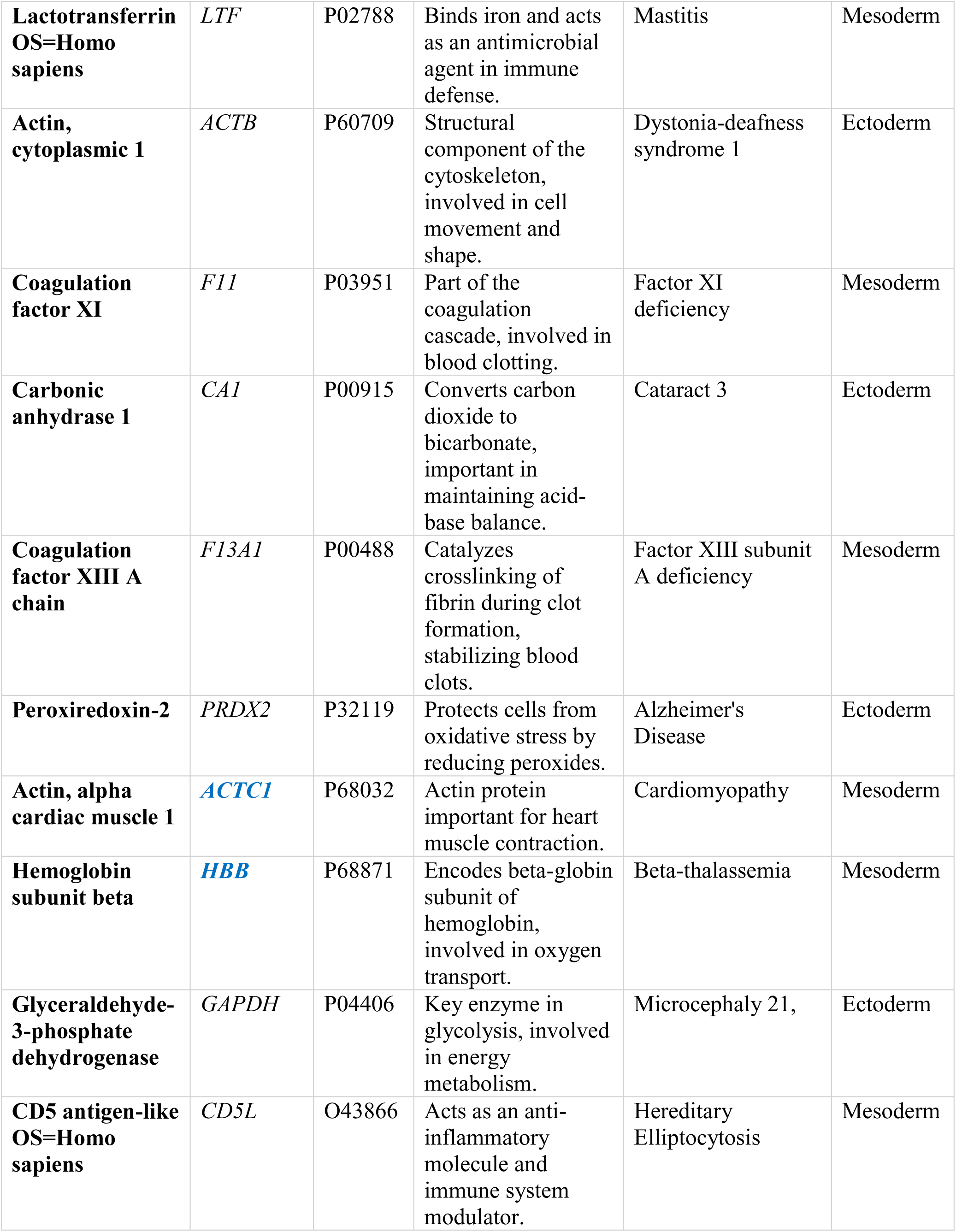
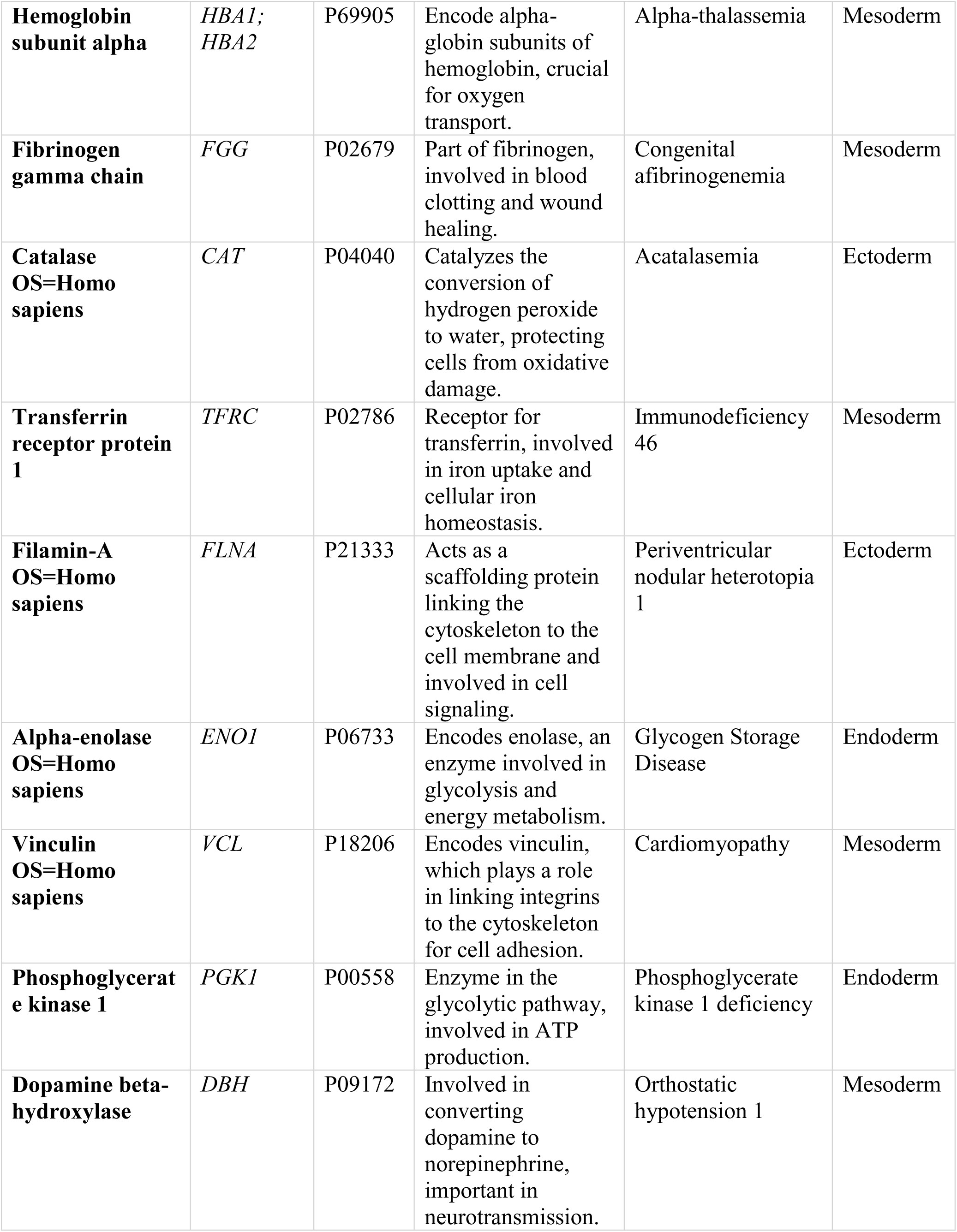
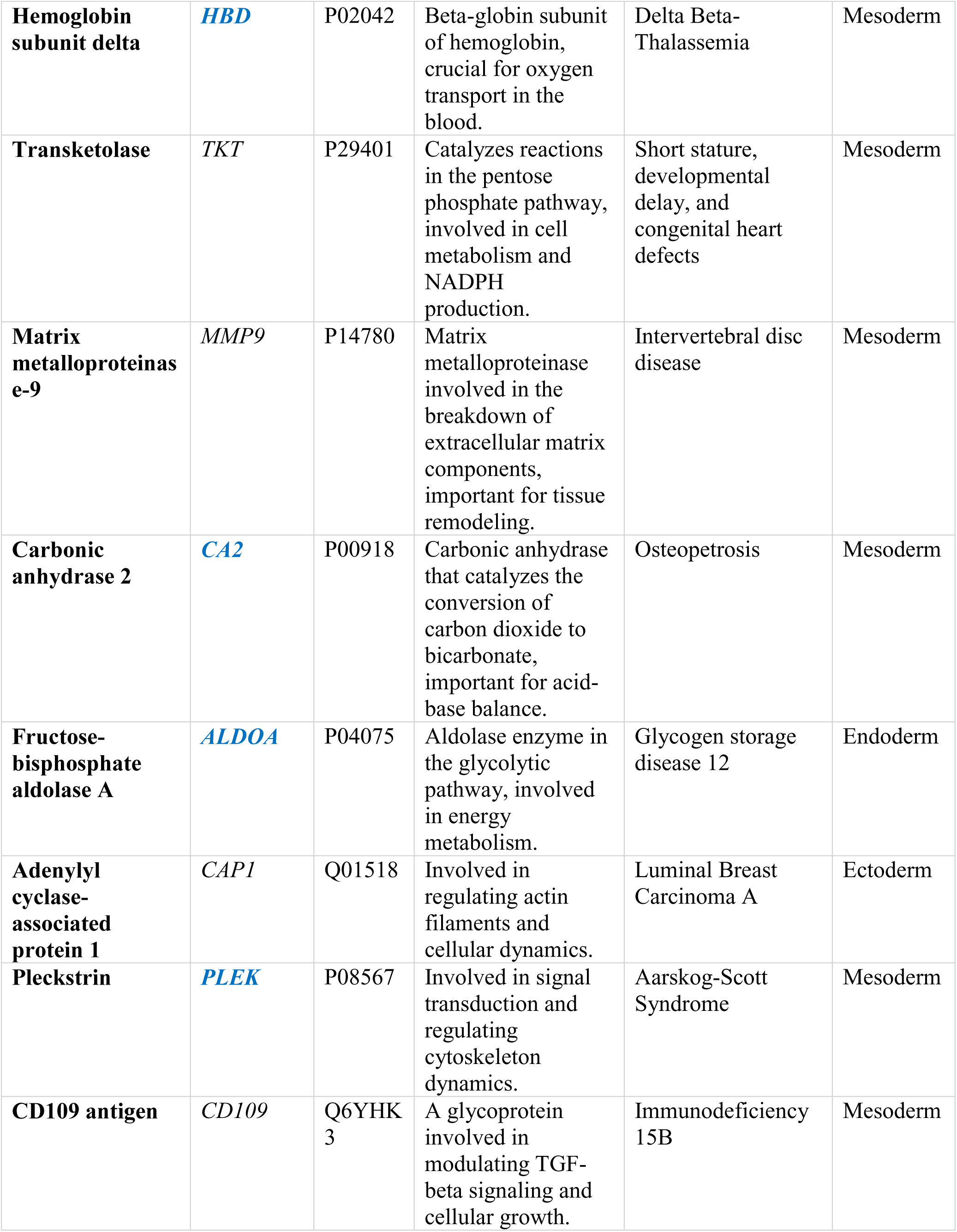
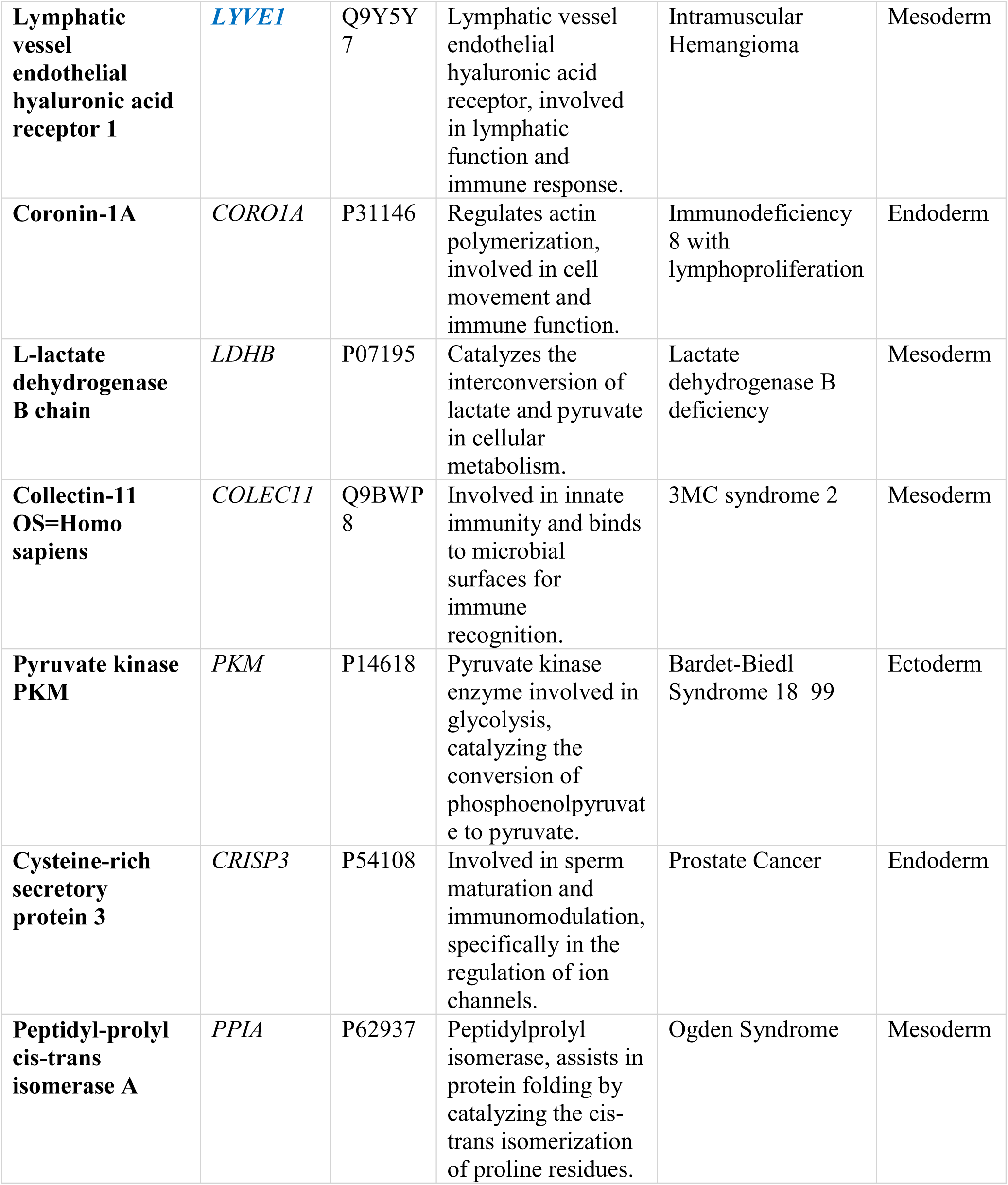
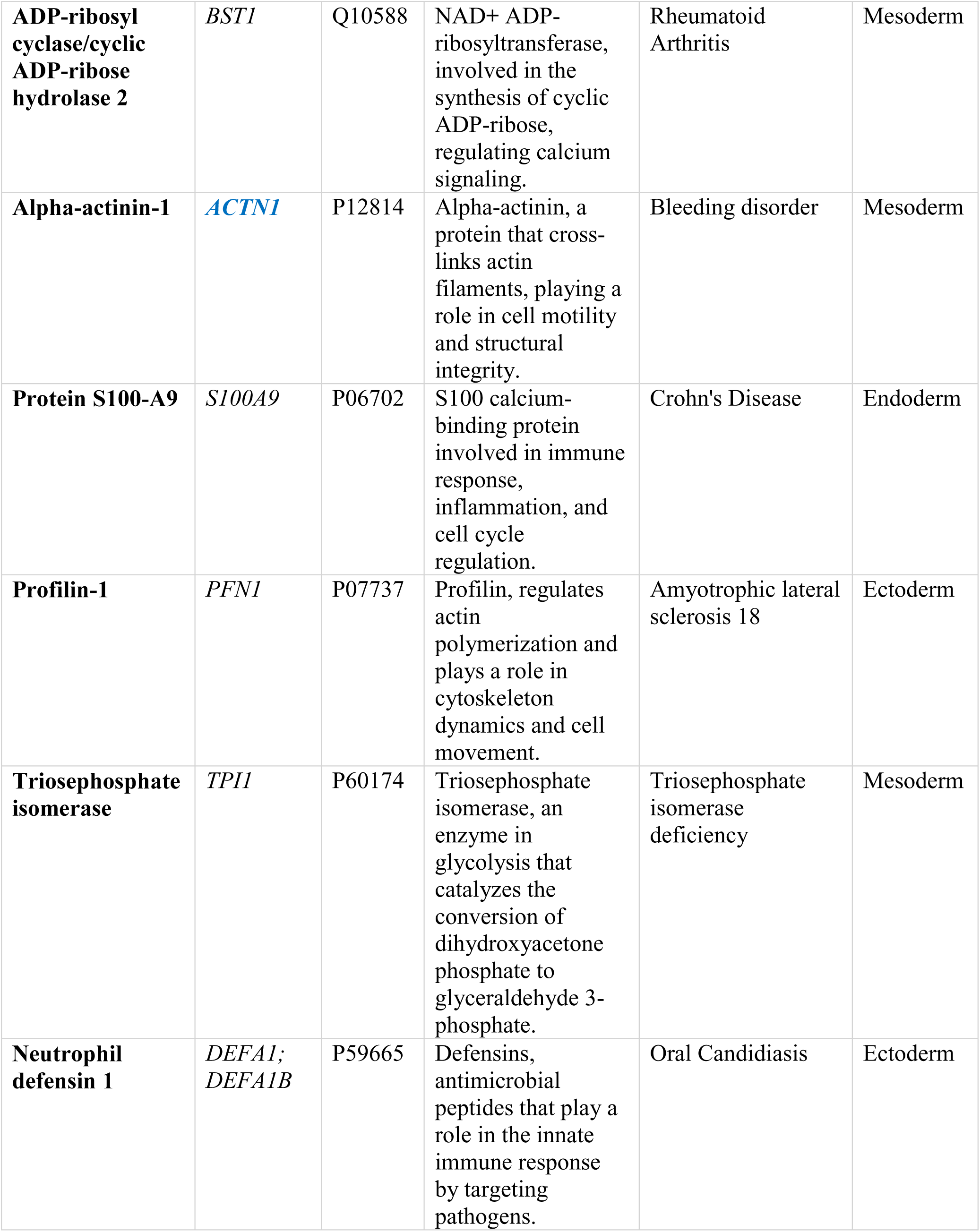
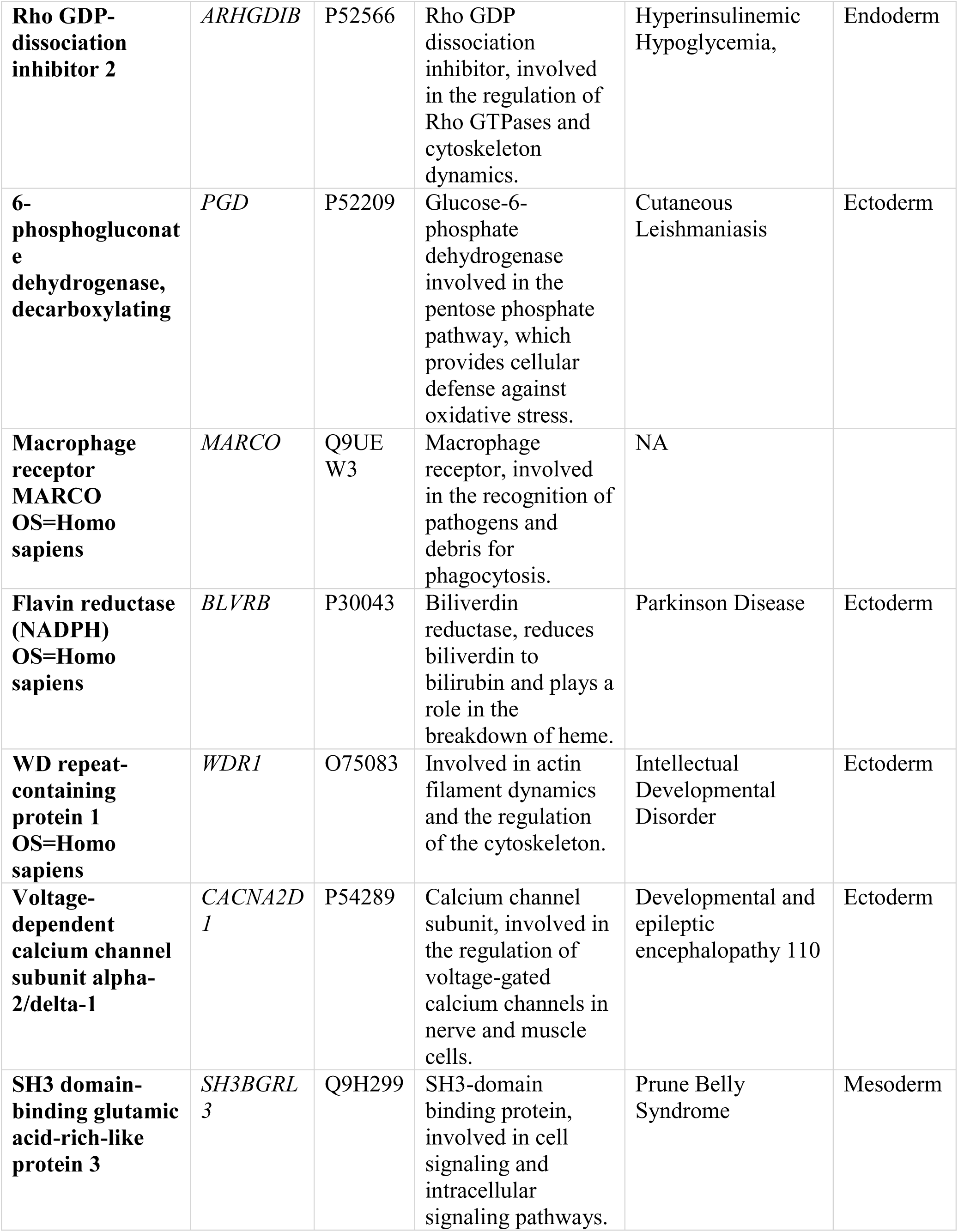
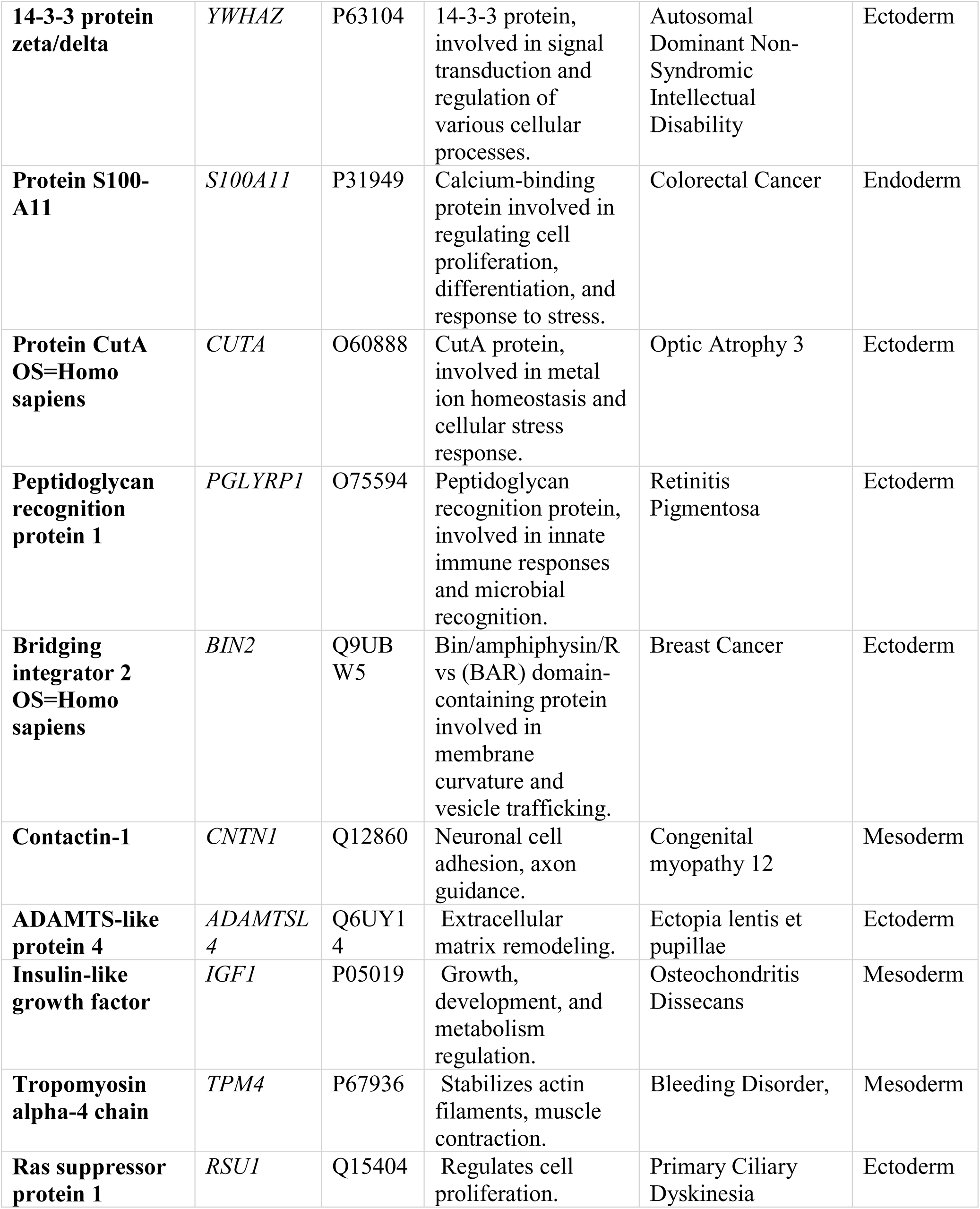
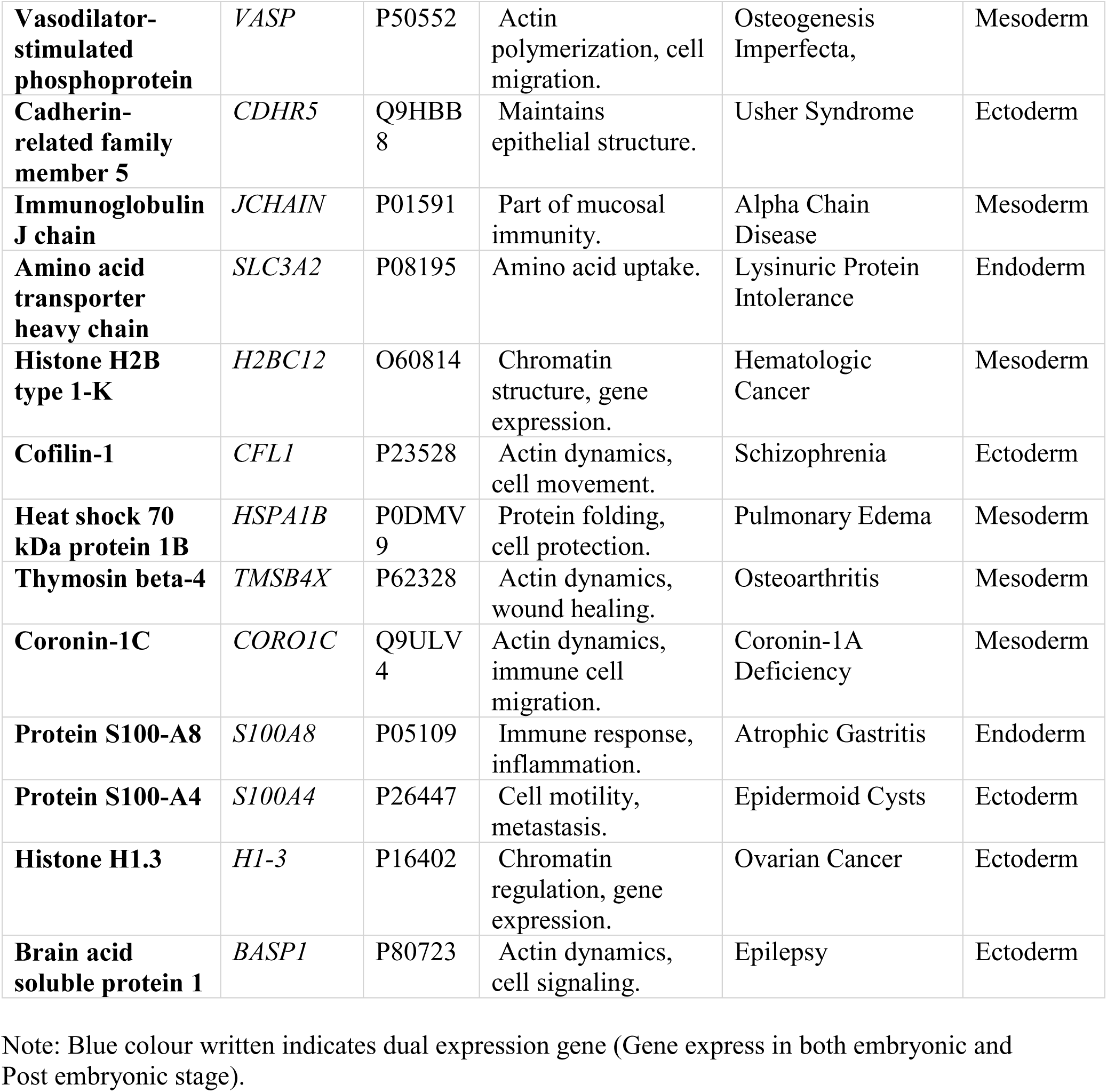
Proteome Analysis of Serum samples of Control vs CTA patients sample.

**Table 3:** Ectomesodermal diseases associated with CTA. This table presents the ectomesodermal diseases linked to congenital tooth agenesis, highlighting their genetic and molecular associations. The diseases are categorized based on their ectodermal and mesodermal origins, with relevant genes, signaling pathways, and clinical manifestations provided for each condition.

| **Top 10 Significant diseases identified from unique metabolites present in control** |  |
| --- | --- |
| **Disease** | **Germ Layer** |
| Isovaleric acidemia | Endoderm |
| Schizophrenia | Ectoderm |
| Colorectal cancer | Mesoderm |
| Alzheimer's disease | Ectoderm |
| Adenomyeloencephalopathy | Mesoderm |
| Premenstrual dysphoric disorder | Mesoderm |
| Juvenile myoclonic epilepsy | Ectoderm |
| Peptic ulcer | Endoderm |
| Refractory localization-related epilepsy | Ectoderm |
| Short-chain L-3-Hydroxyacyl-CoA dehydrogenase deficiency | Endoderm |
| **Top 10 Significant diseases identified from unique metabolites present in Patients** |  |
| **Disease** | **Germ Layer** |
| Schizophrenia | Ectoderm |
| Pregnancy | Mesoderm |
| Pancreatic cancer | Mesoderm |
| Cirrhosis | Endoderm |
| Late-onset preeclampsia | Mesoderm |
| Early preeclampsia | Mesoderm |
| Colorectal cancer | Mesoderm |
| Cerebral creatine deficiency syndrome 1 | Ectoderm |
| Cerebral creatine deficiency syndrome 3 | Ectoderm |
| Menstrual cycle | Mesoderm |
| **Top 10 Significant diseases identified from Up regulated metabolites** |  |
| **Disease** | **Germ Layer** |
| Isovaleric acidemia | Endoderm |
| Schizophrenia | Ectoderm |
| Colorectal cancer | Mesoderm |
| Alzheimer's disease | Ectoderm |
| Adenomyeloencephalopathy | Mesoderm |
| Premenstrual dysphoric disorder | Mesoderm |
| Juvenile myoclonic epilepsy | Ectoderm |
| Peptic ulcer | Endoderm |
| Refractory localization-related epilepsy | Ectoderm |
| Short-chain L-3-Hydroxyacyl-CoA dehydrogenase deficiency | Endoderm |
| **Top 10 Significant diseases identified from Down regulated metabolites** |  |
| **Disease** | **Germ Layer** |
| Primary hypomagnesemia | Mesoderm |
| Schizophrenia | Ectoderm |
| Hypogonadism | Mesoderm |
| D-Lactic Acidosis | Mesoderm |
| Lecithin cholesterol Acyltransferase deficiency | Mesoderm |
| Molybdenum co-factor deficiency | Mesoderm |
| Molybdenum cofactor deficiency | Mesoderm |
| Wolcott-Rallison syndrome | Mesoderm |
| Fanconi Bickel syndrome | Mesoderm |
| Fanconi syndrome | Mesoderm |
| **Top 10 Significant diseases Identified from Proteome Data** |  |

| **Disease** | **Primary Germ Layer(s) Involved** |
| --- | --- |
| Primary Familial Hypertrophic Cardiomyopathy | Mesoderm |
| Familial Dilated Cardiomyopathy | Mesoderm |
| Primary Dilated Cardiomyopathy | Mesoderm |
| Atrial Septal Defect | Mesoderm |
| Cardiomyopathy | Mesoderm |
| Venous Thrombosis | Mesoderm |
| Myocardial Infarction 1 | Mesoderm |
| Susceptibility to Malaria | Mesoderm |
| Glycogen Storage Disease | Endoderm |
| **Top 10 Significant disease identified from Integrative Gene Panel** |  |
| **Disease** | **Germ Layer** |
| Neoplasm of the breast | Ectoderm |
| Familial cancer of the breast | Ectoderm |
| Ashkenazi Jewish disorders | Ectoderm |
| Systemic lupus erythematosus | Ectoderm |
| Charcot-Marie-Tooth disease | Mesoderm |
| Acute myeloid leukemia | Mesoderm |
| Chronic granulomatous disease | Mesoderm |
| Disorder of organic acid metabolism | Endoderm |
| Disorders of intracellular cobalamin metabolism | Endoderm |
| Susceptibility to malaria | Endoderm |

**Table 4.**
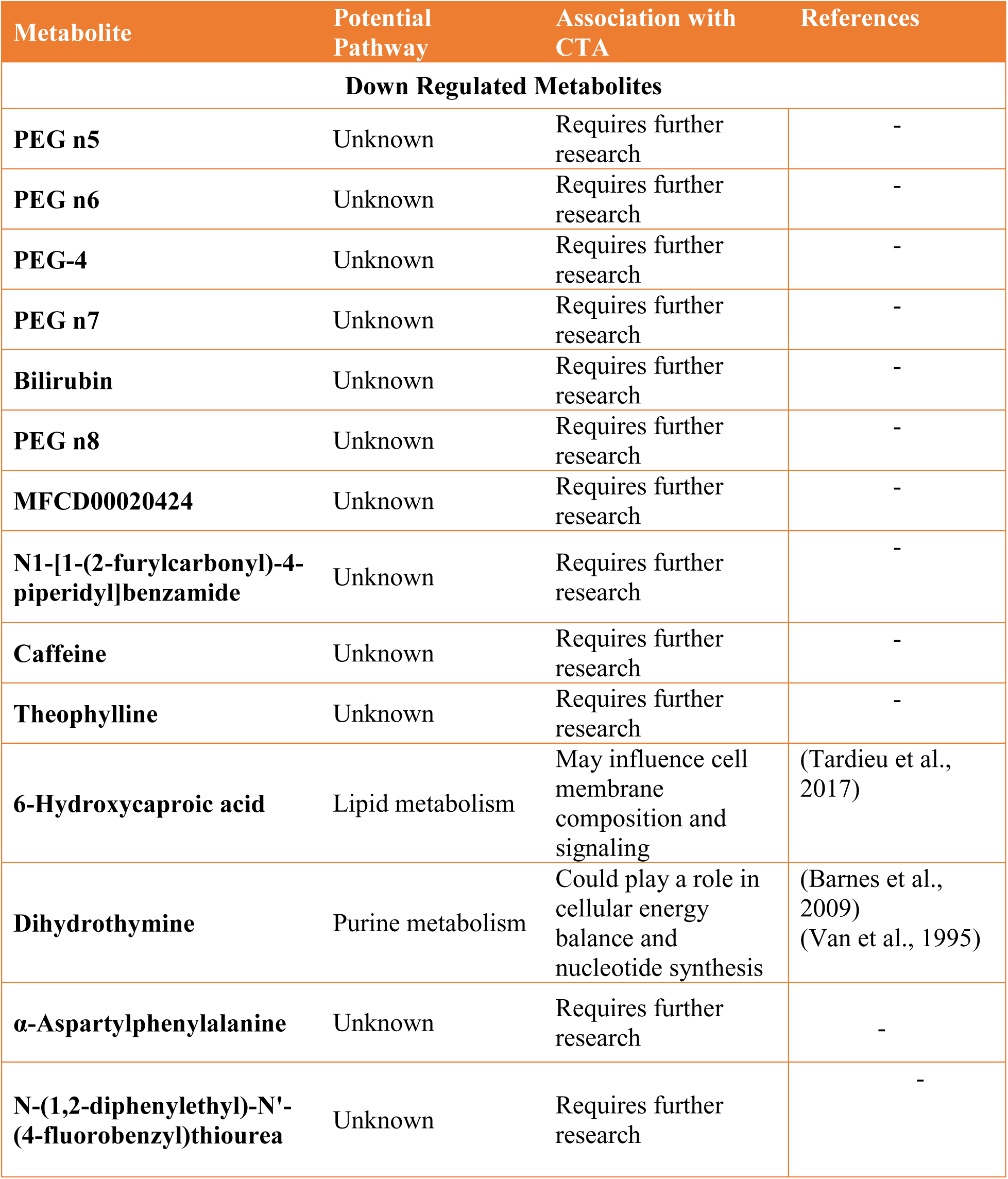
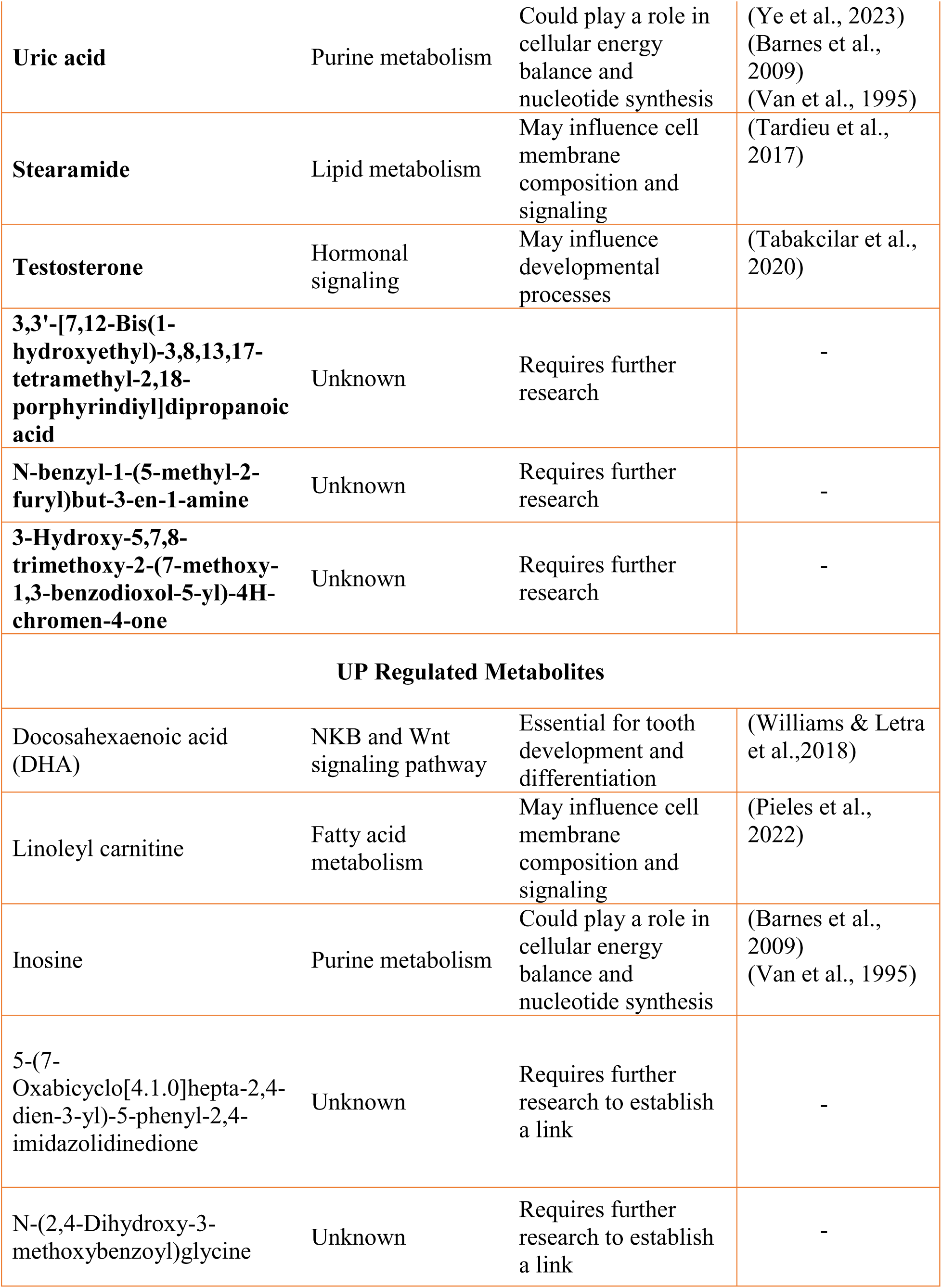
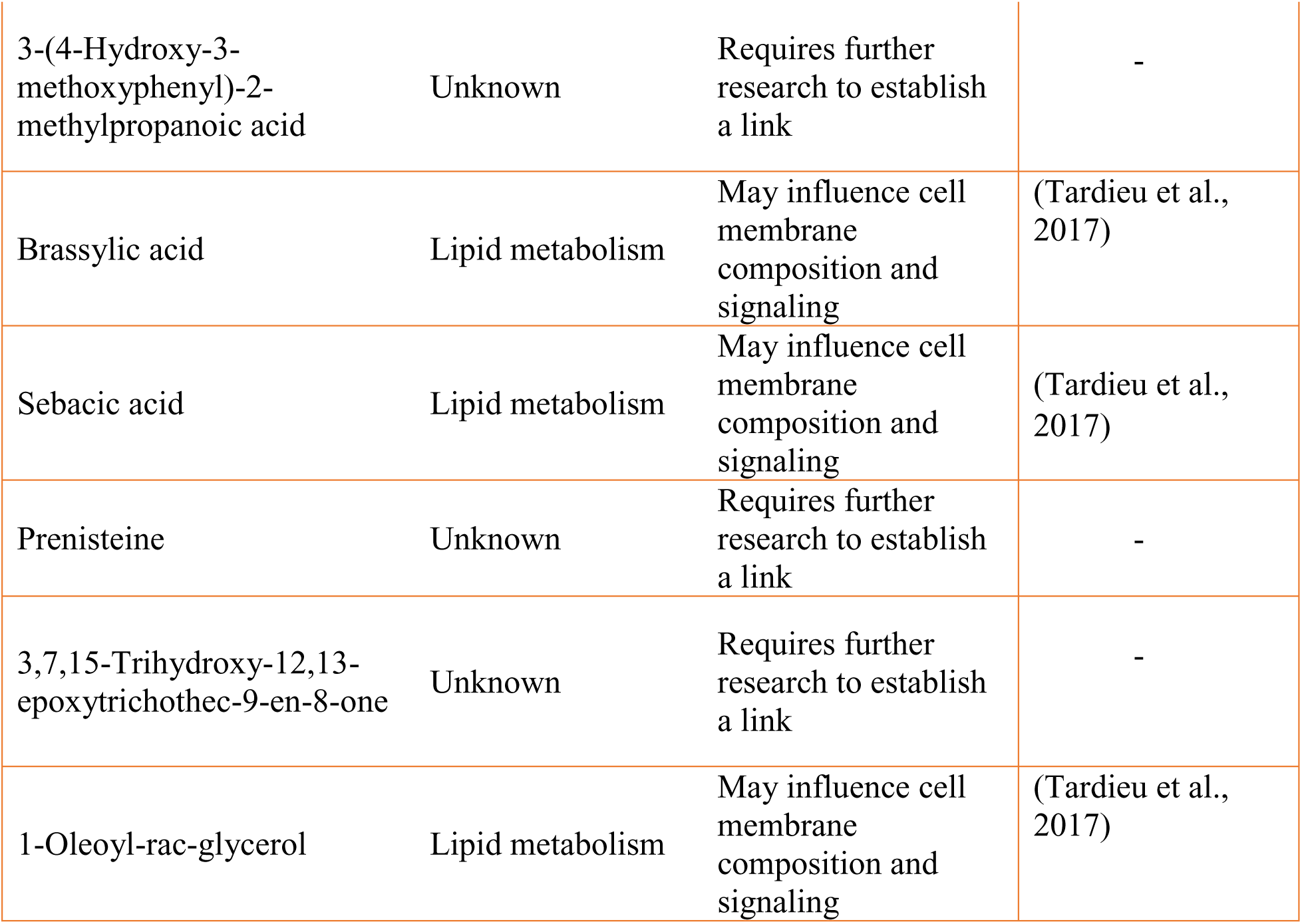
Identification of pathways associated with differentially expressed metabolites, and their linkage to CTA.

**Table 5.**
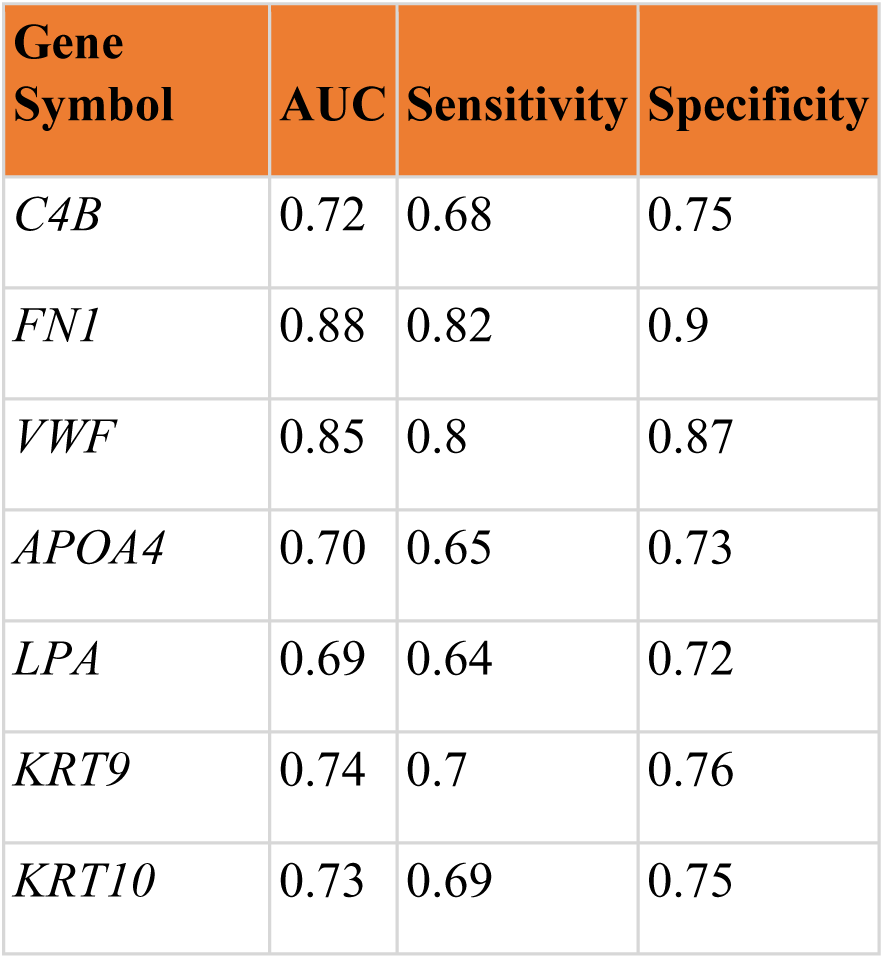
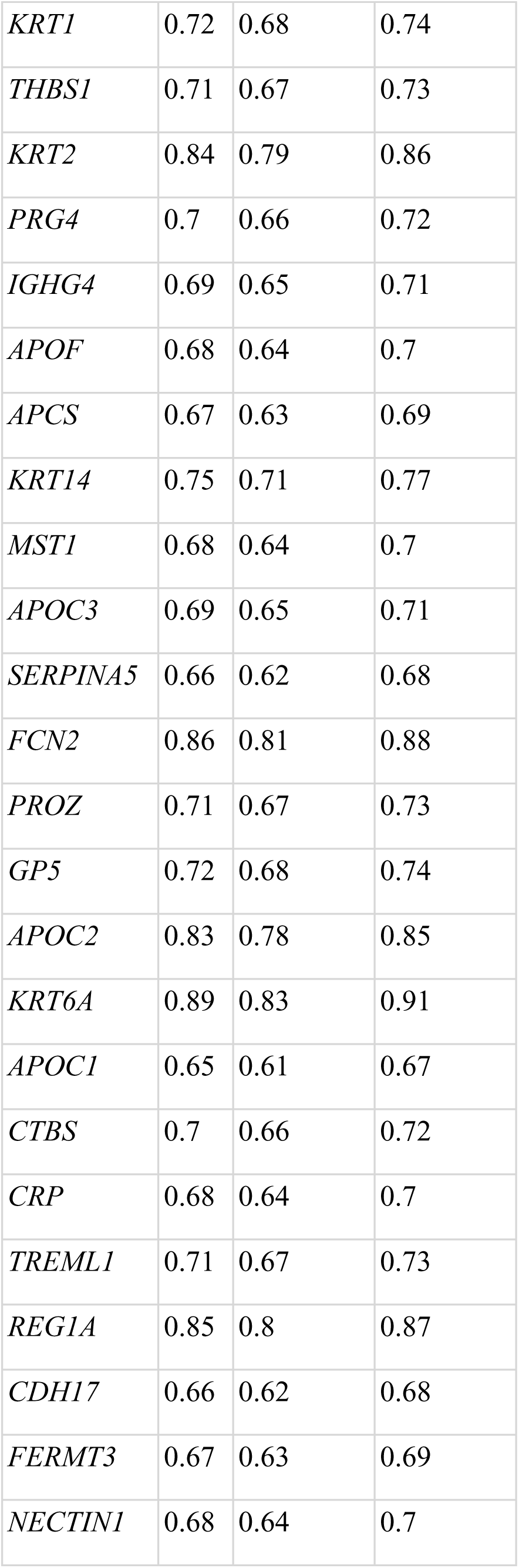
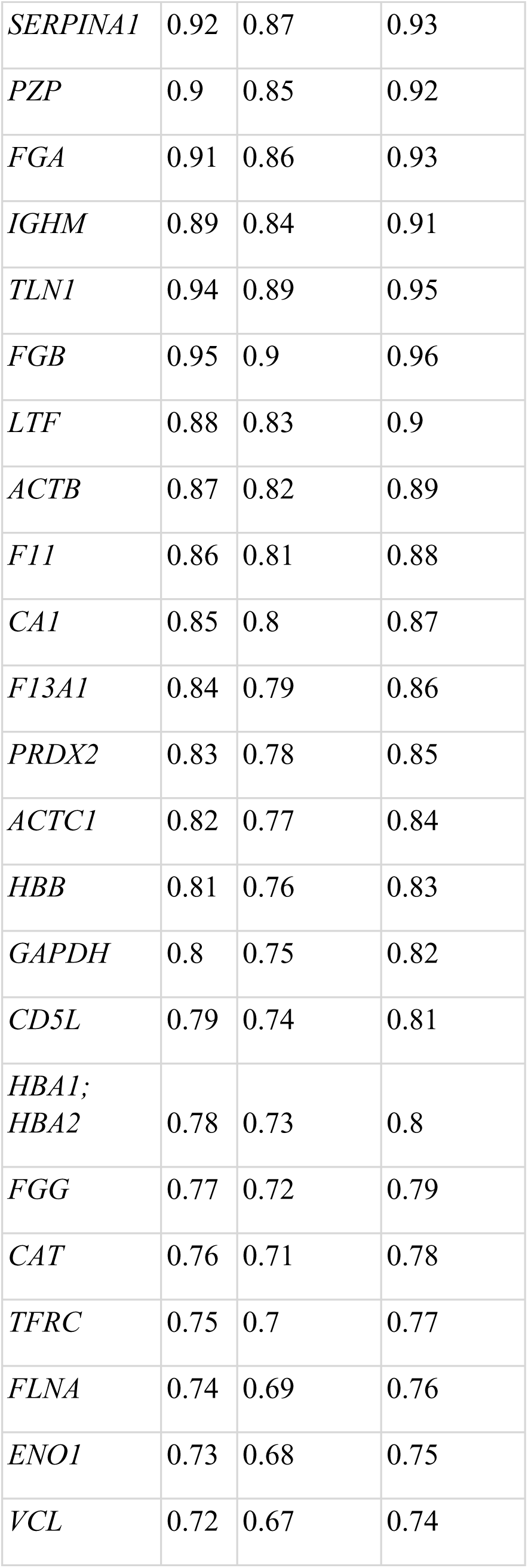
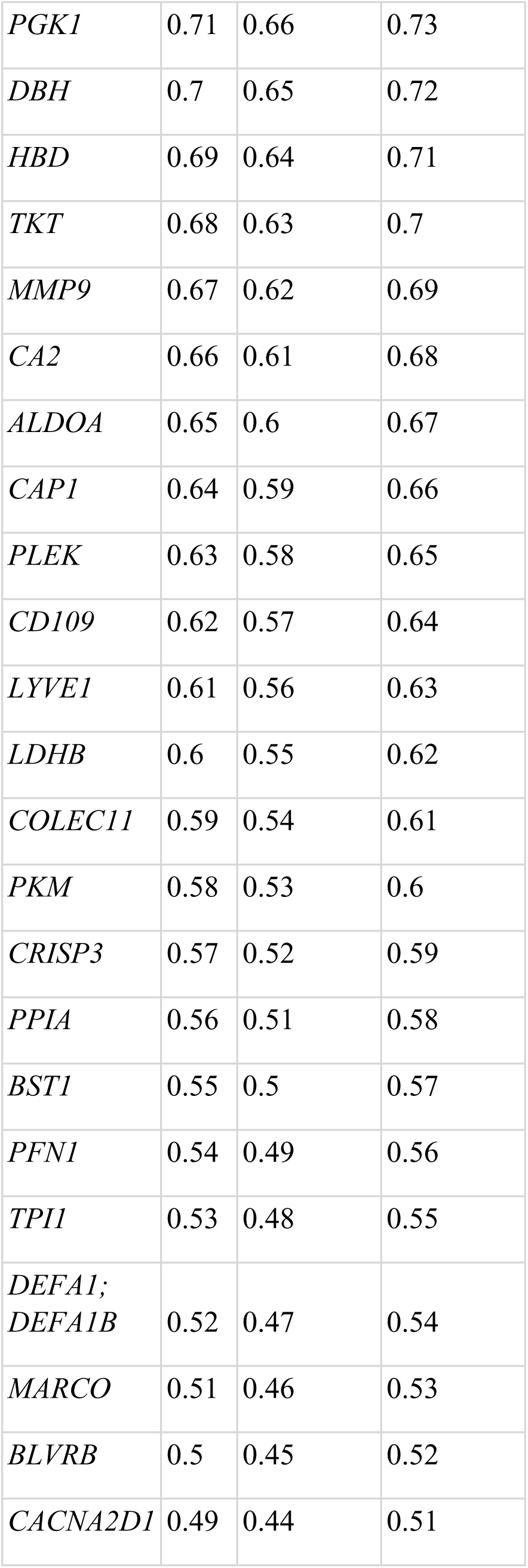
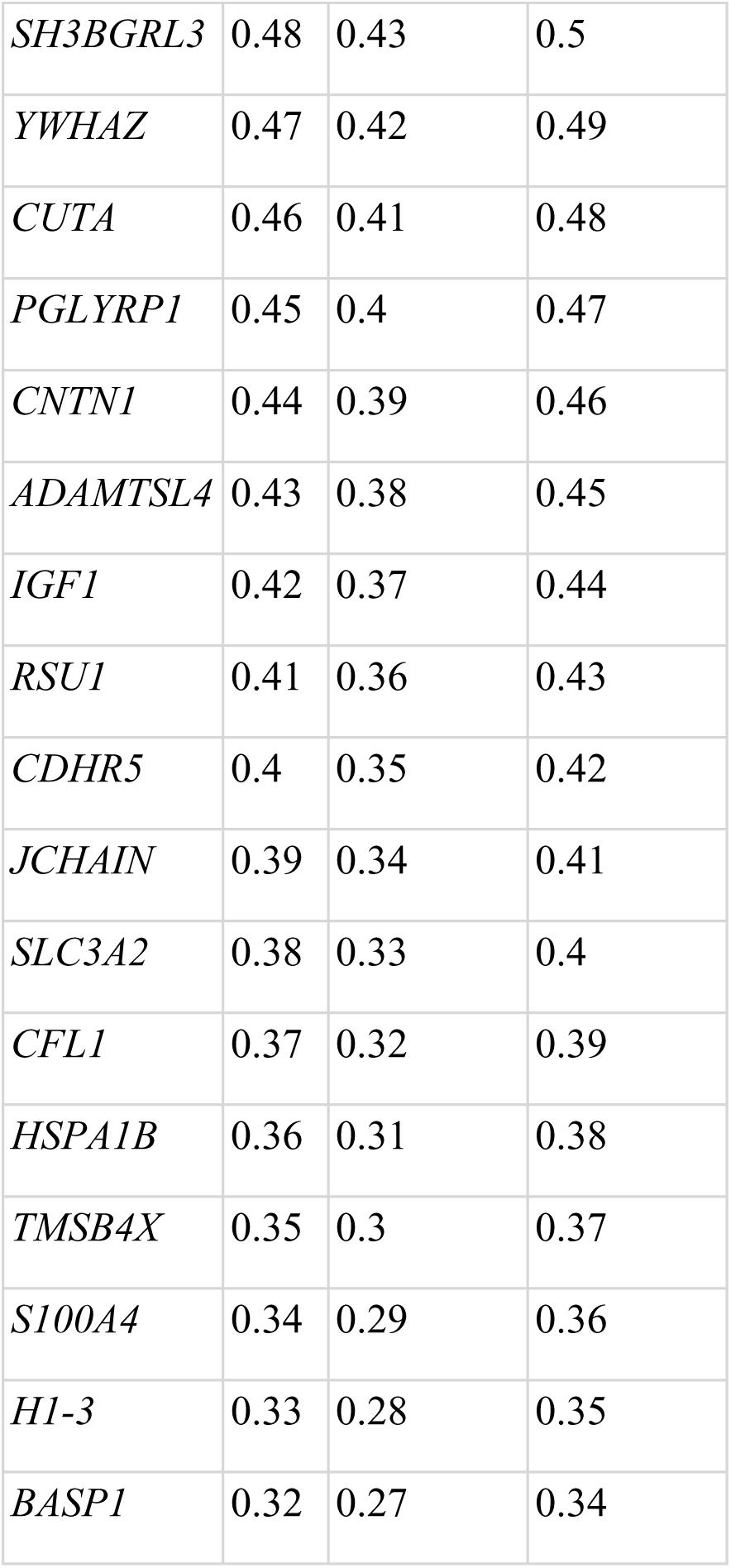
Proteomics Data: AUC, Sensitivity, and Specificity for Gene Markers.

### 3.6 Proteomics Analysis of CTA Patients

Proteomic analysis revealed significant alterations in protein expression between control and CTA patient groups (Figure 8). The 76 genes were upregulated. While 33 were downregulated in CTA individual. The volcano plot identified differentially expressed proteins categorized by significance and fold change. PCA indicated distinct clustering between control and CTA samples, highlighting variance in expression profiles (Figure 8). A heatmap of differentially expressed proteins demonstrated Z-score normalized expression levels, while a circular heatmap depicted upregulated and downregulated proteins with clear differentiation (Figure 8). These findings underscore the potential role of altered protein expression in CTA pathophysiology.

**Figure 8:**
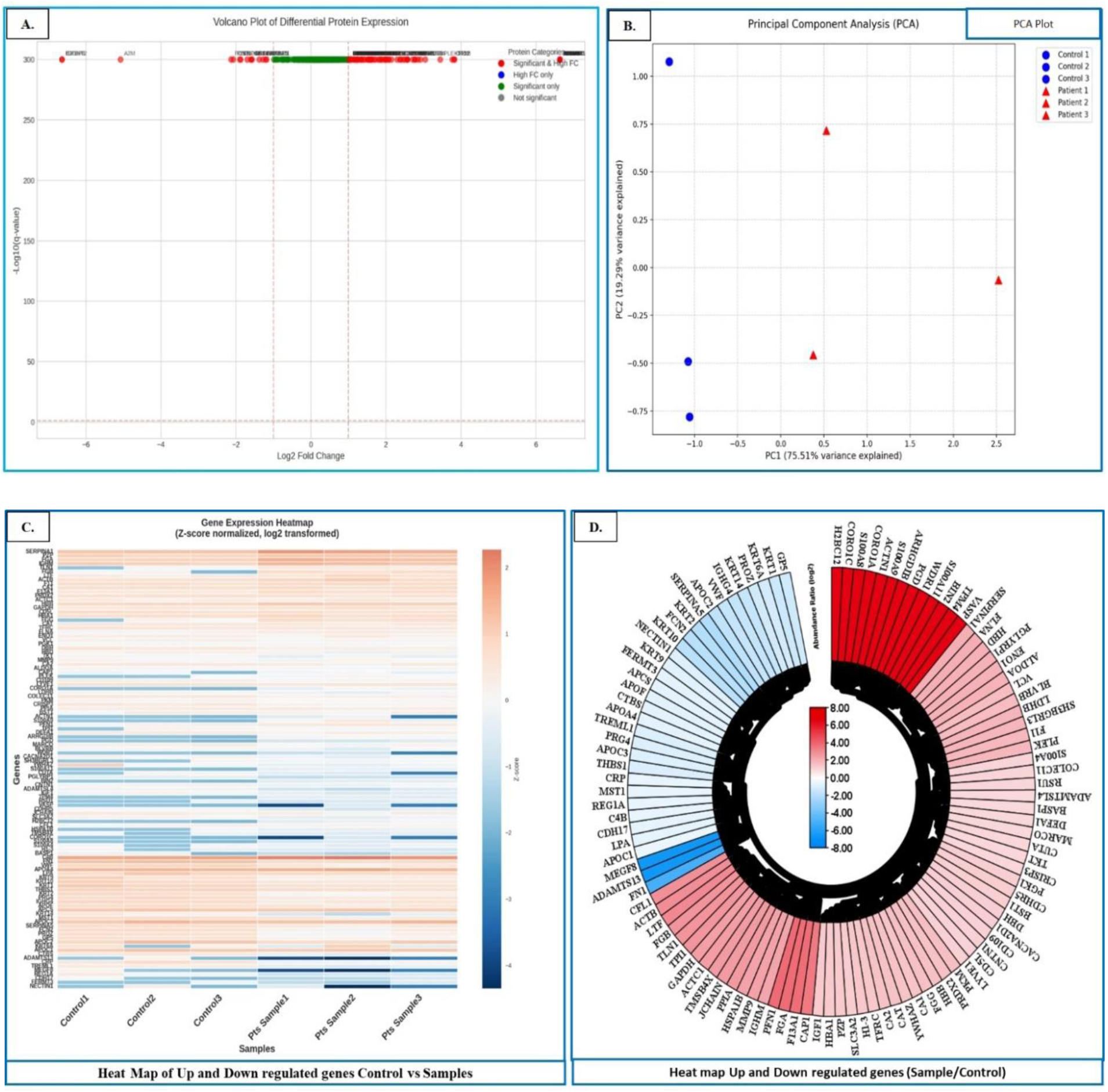
Proteomics Analysis of Control Group and CTA Patient Group A: Volcano Plot of Differential Protein Expression: This plot displays the differential protein expression between the control group and CTA patient group. The x-axis represents the log2 fold change, while the y-axis represents the -log10(p-value). Proteins are categorized based on their significance and fold change: highly significant (red), significant (green), and not significant (gray). B: Principal Component Analysis (PCA) Plot This PCA plot illustrates the variance in protein expression between the control group and CTA patient group. The x-axis (PC1) and y-axis (PC2) show the principal components that explain the highest variance. Control samples are represented by blue circles, and CTA patient samples are represented by red triangles, indicating distinct clustering between the two groups. C: Heatmap of Differentially Expressed Proteins. This heatmap shows the Z-score normalized, log2 transformed expression levels of differentially expressed proteins in the control and CTA patient groups. The x-axis represents the samples (Control 1, Control 2, Control 3, Patient 1, Patient 2, Patient 3), and the y-axis lists the proteins. The color gradient ranges from blue (low expression) to red (high expression), highlighting differences in protein expression levels. D: Circular Heatmap of Upregulated and Downregulated Proteins. This circular heatmap visualizes the log2 fold change of upregulated and downregulated proteins in CTA patient samples compared to control samples. The inner circle represents upregulated proteins (red), and the outer circle represents downregulated proteins (green). Each segment corresponds to a protein, providing a clear depiction of the differential expression patterns.

### 3.7 Proteomics Enrichment Analysis

Enrichment analysis of the proteomics data revealed significant alterations in protein expression and pathway involvement in CTA (Figure 9). Upregulated proteins were predominantly associated with biological processes such as cell adhesion, hemostasis, and immune response, indicating potential roles in compensatory mechanisms or aberrant tissue responses in CTA (Figure 9). In contrast, downregulated proteins were linked to peptide cross-linking and intermediate filament organization, processes critical for maintaining tissue structural integrity (Figure 9). The downregulation of these pathways likely contributes to the impaired development observed in CTA. These findings highlight distinct proteomic changes that may inform future investigations into the pathogenesis of the condition. Disease association analysis of proteome DEGs predominantly linked them to ectodermal and mesodermal diseases (Table 2 and Table3).

**Figure 9:**
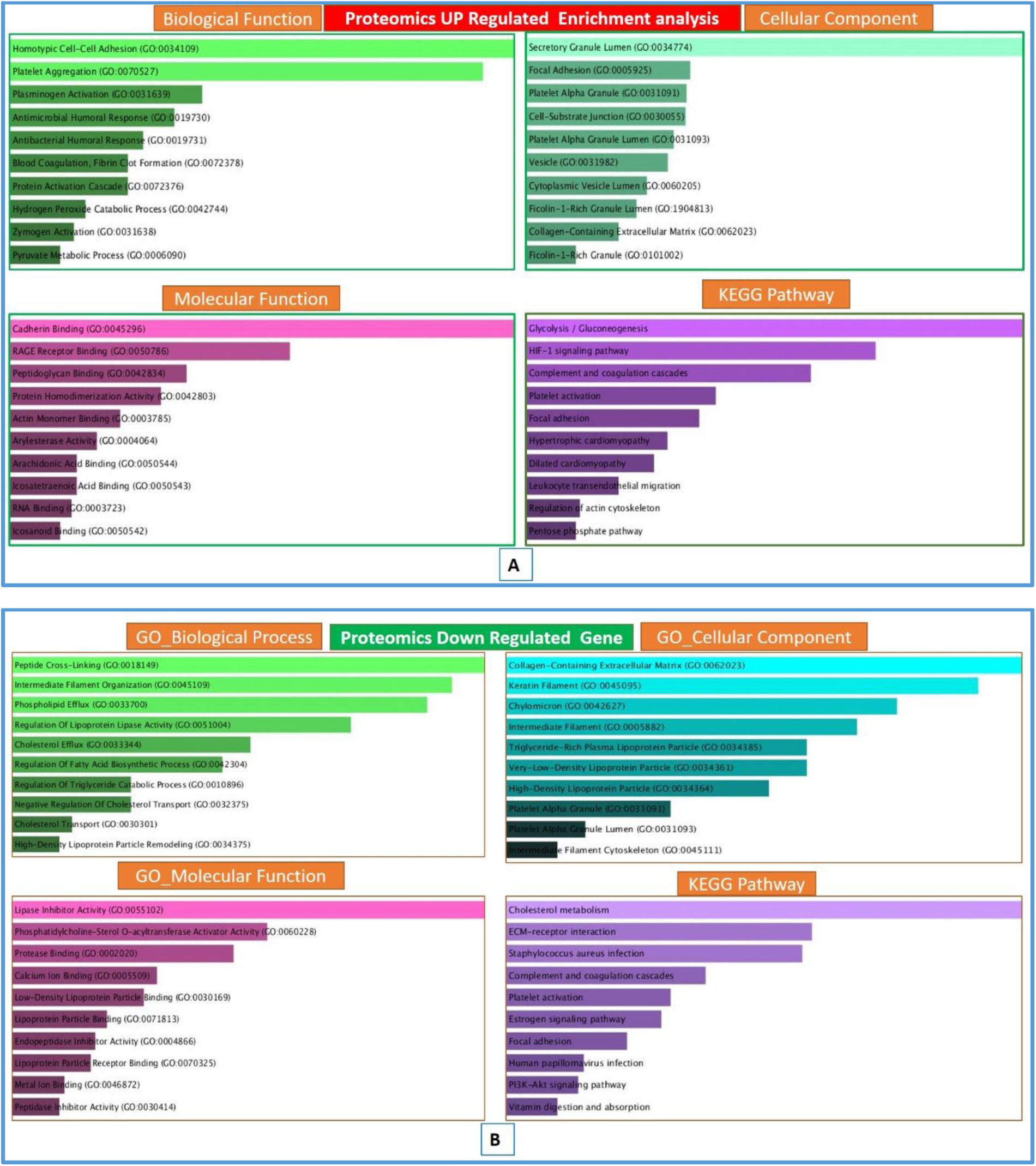
Figure: Proteomics Enrichment Analysis of CTA. **(A)** Upregulated proteins enrichment analysis, showing categories including Biological Process, Cellular Component, and KEGG Pathways. **(B)** Downregulated proteins enrichment analysis, highlighting Biological Process, Molecular Function, Cellular Component, and KEGG Pathways.

### 3.8 Proteome Gene Network and Overlapping studies with different panel Analysis

Gene network analysis of the proteomics data revealed key insights into differential regulation, hub genes, and gene panel overlaps (Figure 10). The combined gene network of upregulated and downregulated genes, generated using STRING, highlighted developmental process-associated genes (yellow nodes), underscoring their potential roles in CTA (Figure 10). Hub gene analysis identified highly connected 20 genes such as *ACTB*, *ENO1*, *GAPDH*, *TPI1* etc., emphasizing their central roles in the network (Figure 10). The Venn diagram analysis illustrated gene overlaps among the OMIM, Proteomics, and Gene Card panels. *COLEC11* was identified as the only common gene across all three panels (Figure 10). The Proteomics Panel had 76 unique genes, while the Gene Card Panel contained 4759 unique genes. Additionally, 32 genes were overlapped between the Proteomics and Gene Card Panels, and 126 genes were common between the OMIM and Gene Card Panels (Figure 10). These findings highlight critical genes and interactions that may contribute to the pathogenesis of CTA.

**Figure 10:**
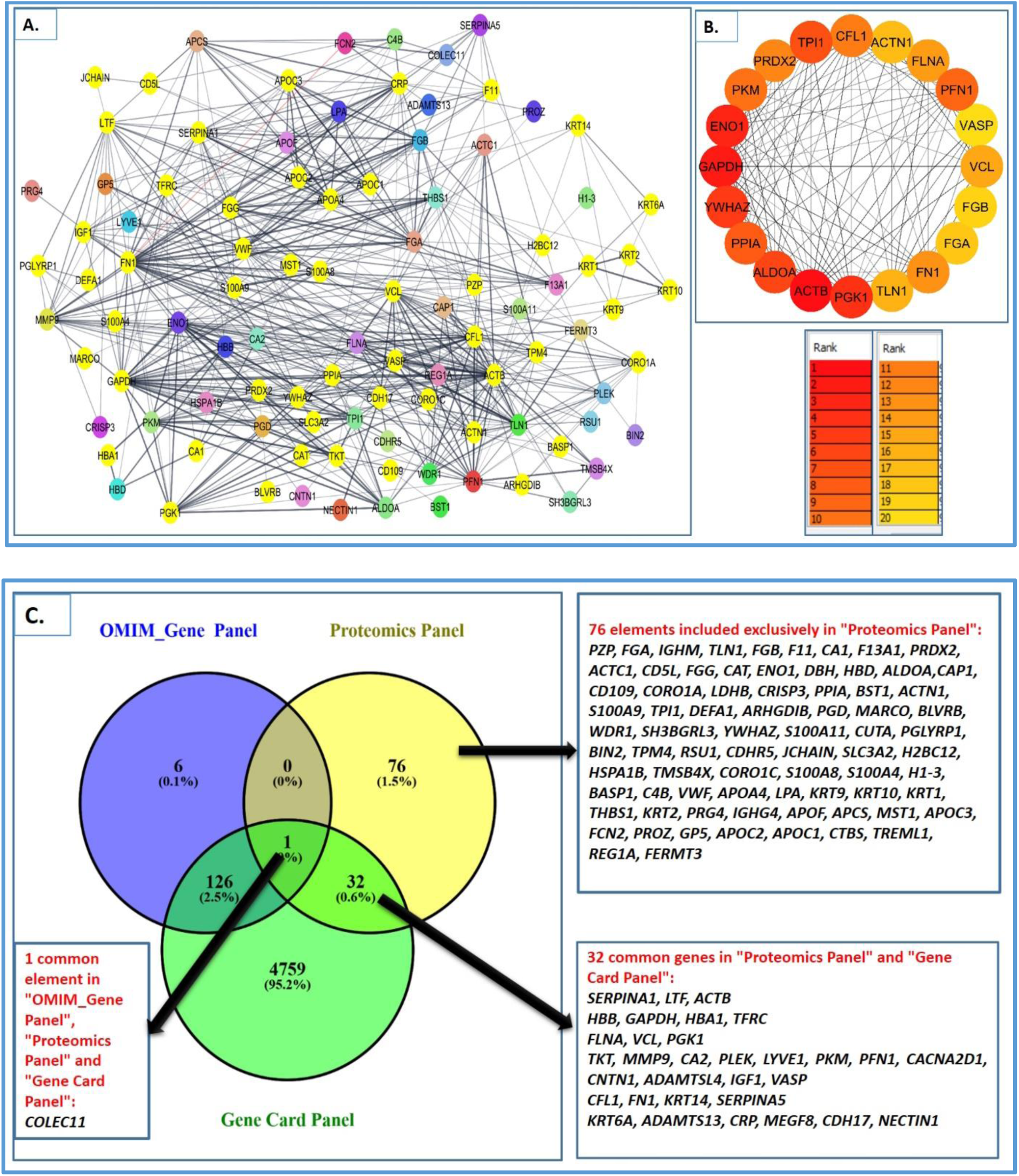
Proteome Gene Network Analysis: Differential Regulation, Hub Gene Insights, and Mapping to OMIM and Gene Cards Databases. A. Combined Proteome Up and Down Regulated Gene Network. This panel displays the combined gene network of upregulated and downregulated genes derived from STRING database analysis. The network nodes represent individual genes, and edges signify interactions between these genes. Yellow-highlighted nodes indicate genes that are specifically associated with developmental processes, emphasizing their potential role in CTA. B. Hub Gene Identification: This panel identifies the hub genes within the network, which are the most connected and potentially influential genes. Hub genes are arranged in a circular layout, with color intensity indicating their rank based on connectivity. Darker colors represent higher ranks, highlighting central genes such as *ACTB, ENO1, GAPDH, TPI1* etc. C. Venn Diagram of Gene Panels: This panel shows the overlap between three different gene panels: OMIM_Gene Panel, Proteomics Panel, and Gene Card Panel. The Venn diagram illustrates the distribution of unique and shared genes across the panels. The OMIM_Gene Panel contains 6 unique genes (0.1%), the Proteomics Panel has 76 unique genes (1.5%), and the Gene Card Panel includes 4759 unique genes (95.2%). Notably, *COLEC11* is the only gene common to all three panels. Additionally, there are 32 overlapped genes between the Proteomics Panel and Gene Card Panel, while 76 genes are novel proteome gene and 126 genes are common between the OMIM Gene Panel and Gene Card Panel.

### 3.9 Machine Learning

#### 3.9.1 Proteome Data Analysis

The model demonstrated excellent performance in distinguishing between the “Control” and “Sample” groups, as evidenced by an AUC-ROC score of 0.95. The sensitivity of the model was 0.92, indicating that it correctly identified 92% of the “Sample” group (true positive rate). Additionally, the specificity was 0.94, reflecting the model’s ability to correctly identify 94% of the “Control” group (true negative rate). These metrics collectively highlight the model’s robust capability to differentiate between the two groups with high accuracy.

Proteomic analysis identified multiple candidate biomarkers for CTA, with area under the curve (AUC) values ranging from 0.95 (FGB) to 0.32 (BASP1). High-performing biomarkers (AUC ≥ 0.85) included SERPINA1 (0.92), PZP (0.90), FGA (0.91), IGHM (0.89), TLN1 (0.94), and FGB (0.95), demonstrating strong sensitivity and specificity. FN1 (0.88), VWF (0.85), and REG1A (0.85) also showed significant discriminatory potential (Table5). Moderate-performing proteins (0.70 ≤ AUC < 0.85) included KRT2, PRDX2, HBB, GAPDH, and FLNA. Low-performing proteins (AUC < 0.70) were less reliable for CTA classification (Table5). These findings highlight key proteomic markers that may aid in CTA diagnosis and further understanding of its pathophysiology.

#### 3.9.2 Metabolite Data Analysis

The model exhibited exceptional performance in distinguishing between the “Control” and “Sample” groups, achieving an AUC-ROC score of 0.98. The sensitivity was 0.96, indicating that the model correctly identified 96% of the “Sample” group. Furthermore, the specificity was 0.97, demonstrating the model’s ability to correctly identify 97% of the “Control” group. Feature importance analysis revealed that the three key features (Control1, Control2, and Control3) contributed almost equally to the model’s predictive performance. This underscores the balanced contribution of these features to the model’s high discriminative power.

Metabolomics analysis identified a panel of potential biomarkers for CTA based on area under the curve (AUC), sensitivity, and specificity. High-performing metabolites (AUC ≥ 0.90) included PEG n5 (0.99), PEG n6 (0.98), PEG-4 (0.97), PEG n7 (0.96), PEG n8 (0.95), and caffeine (0.94), demonstrating strong discriminatory potential with sensitivity and specificity values exceeding 0.90 (Table 6). Other notable metabolites included structurally similar compounds such as NP- 007909 (AUC: 0.93), hydroxycaproic acid (0.91), and α-aspartylphenylalanine (0.90). Moderate-performing metabolites (0.70 ≤ AUC < 0.90) included compounds such as N-(1,2-diphenylethyl)- N’-(4-fluorobenzyl)thiourea (0.89), 3-oxoindane-1-carboxylic acid (0.88), and benazol P (0.87), along with several identified analogs and derivatives (Table 6). These metabolites demonstrated a balance between sensitivity and specificity but with relatively lower performance compared to the high-performing biomarkers. Low-performing metabolites (AUC < 0.70) included compounds such as leu-phe (0.69), leu-leu (0.67), and several structural analogues with AUC values near 0.50, suggesting limited diagnostic utility (Table 6). The lowest-performing metabolites included 8- hydroxy-4-methoxy-7-methyl-7,8-dihydro-5H-furo [2,3-g] isochromen-5-one (0.45) and other compounds with AUC values below 0.50, indicating minimal discriminatory potential (Table 6). These findings highlight a subset of metabolite biomarkers with high predictive value for CTA, providing insight into potential metabolic pathways involved in its pathophysiology.

**Table 6.**
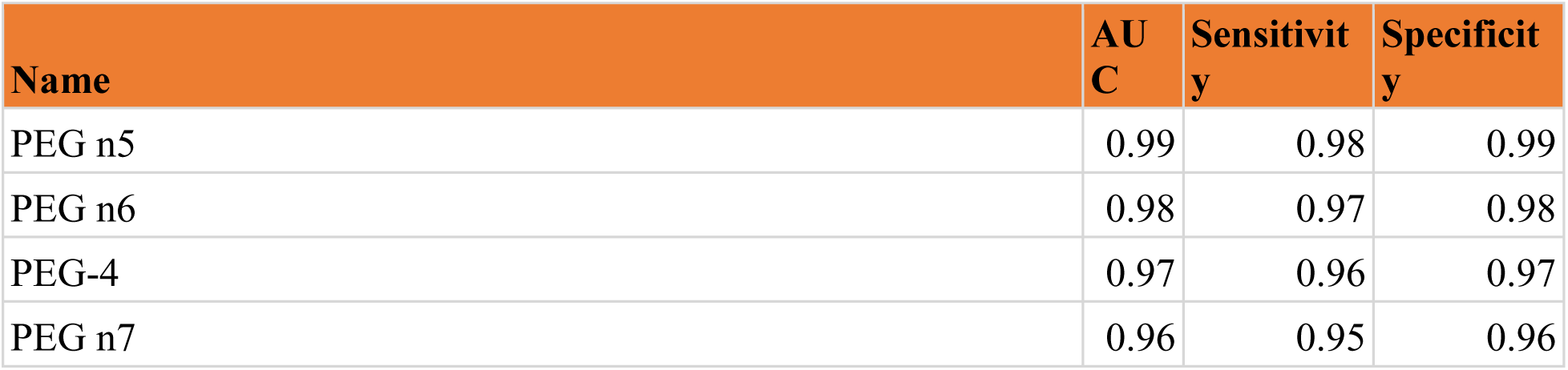
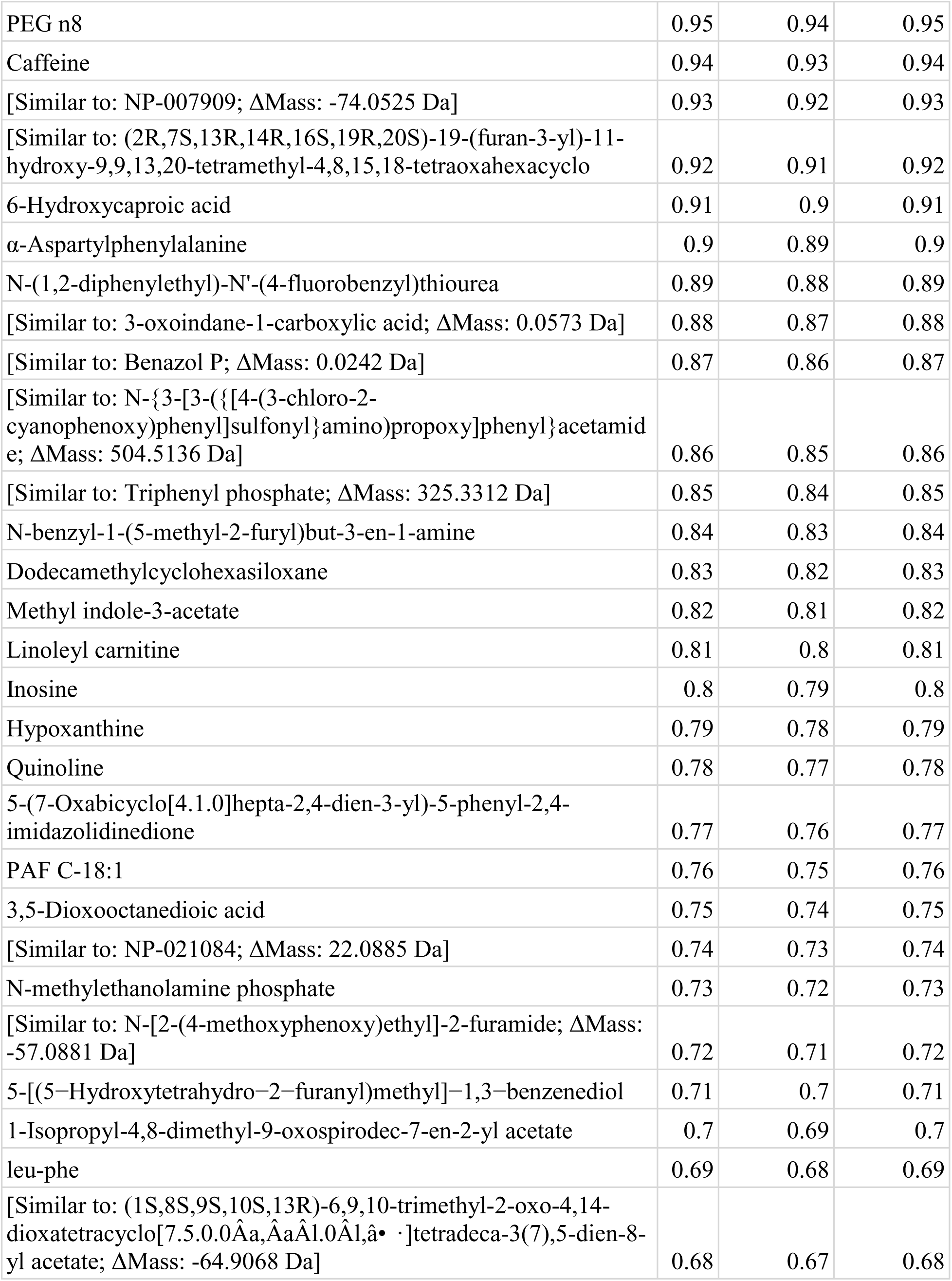
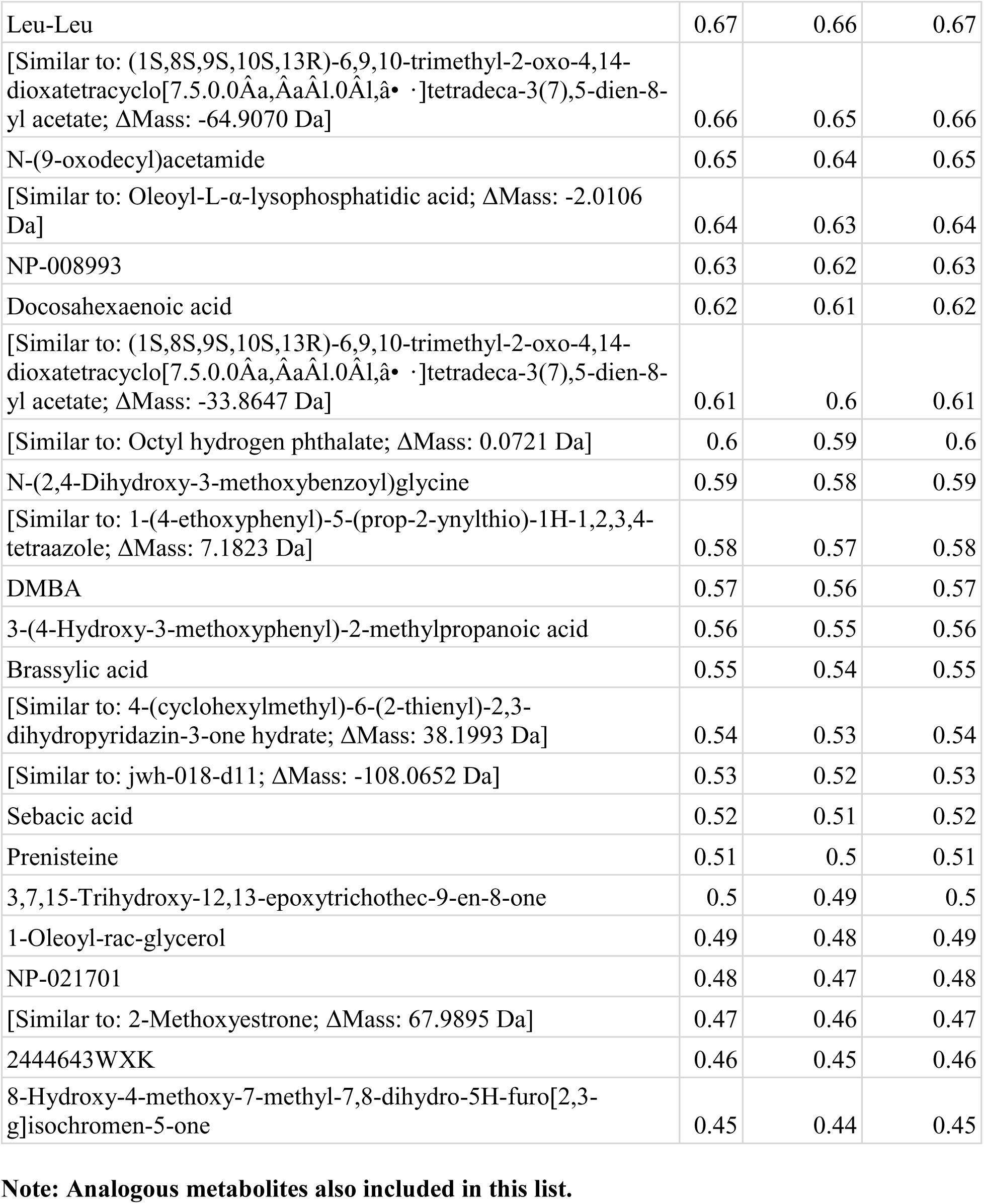
Metabolites Data: AUC, Sensitivity, and Specificity for Metabolite Markers.

#### 3.9.3 Pan-Omics Fusion

Six genes were identified as common between Microarray UP DEGs and Proteomics UP DEGs, while one gene was overlapped between Proteomics Down DEGs and Microarray Down DEGs (Figure 11). Two genes overlapped between Proteomics Down DEGs and the Metabolite Panel, whereas 11 genes were overlapped between Proteomics UP DEGs and the Metabolite Panel (Figure 11). Furthermore, 55 genes were common between Microarray UP DEGs and the Metabolite Panel, and 57 genes overlapped between Microarray Down DEGs and the Metabolite Panel (Figure 11). Analysis also revealed 15 overlapping genes between Microarray Down DEGs and the WES Panel, and 18 genes were overlapped between Microarray UP DEGs and the WES Panel (Figure 11). Four genes were identified as common between Proteomics UP DEGs and the WES Panel, while one gene was overlapped between Proteomics Down DEGs and the WES Panel (Figure 11). Additionally, nine genes overlapped between the Proteomics Panel (up & down DEGs) and the Microarray DEGs Panel, with one gene shared across the Proteomics Panel, Microarray DEGs Panel, and Metabolite Panel (Figure 11). The overlap analysis also indicated 12 overlapped genes between the Proteomics Panel and Metabolite Panel, five genes between the Proteomics Panel and WES Panel, and 27 genes between the Metabolite Panel and WES Panel (Figure 11). Lastly, 33 genes overlapped between the Microarray DEGs Panel and WES Panel, while 111 genes were overlapped between the Microarray DEGs Panel and Metabolite Panel (Figure 11).

**Figure 11:**
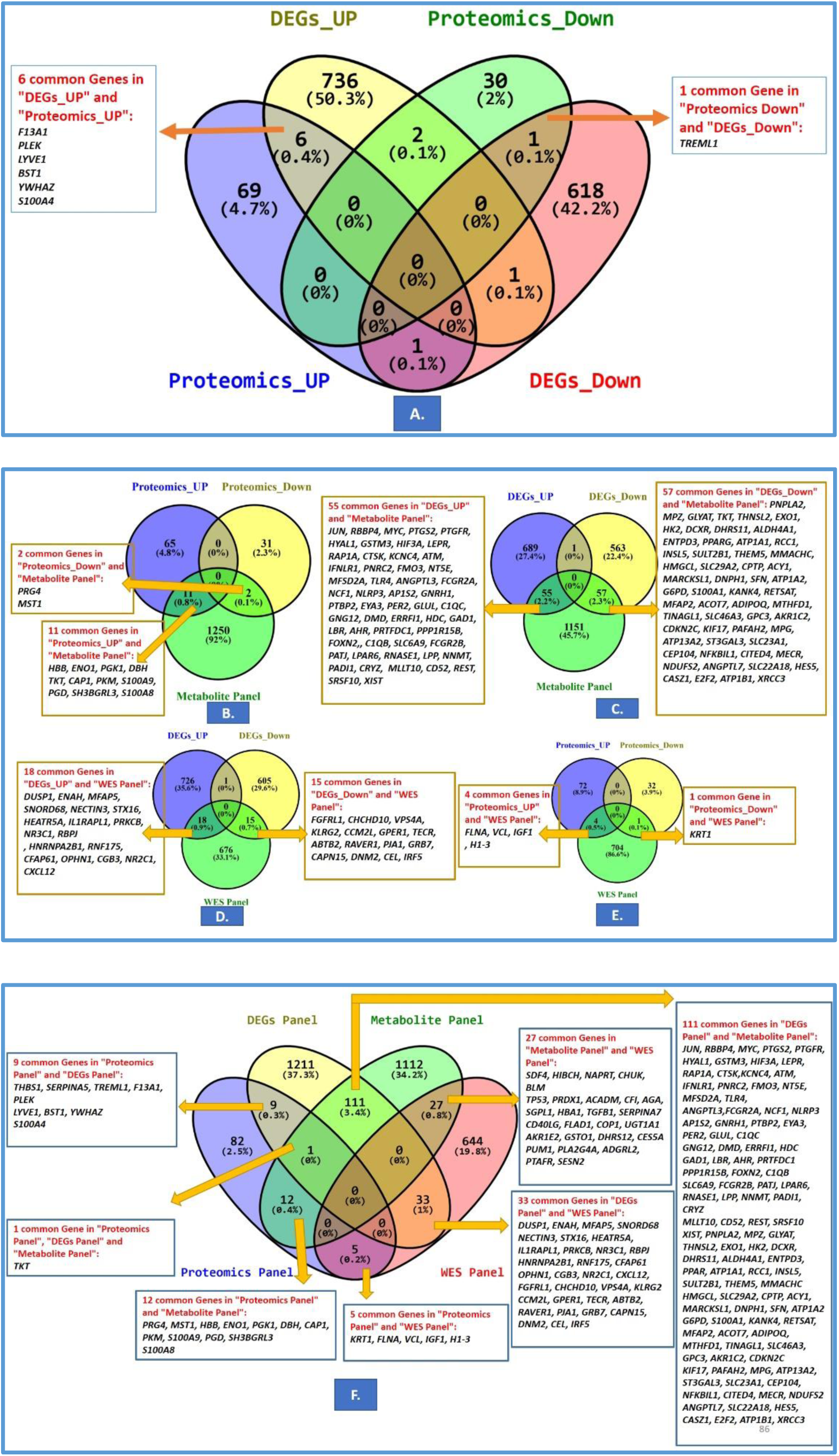
The Venn diagram illustrates the overlap and unique genes across Microarray DEGs, Proteomics DEGs, Metabolomics, and WES panels. Note: Microarray Upregulated DEGs were represented as DEGs_UP, while Downregulated DEGs were noted as DEGs_Down. Similarly, Proteomics Upregulated DEGs were denoted as Proteomics_UP, and Downregulated DEGs as Proteomics_Down.

#### 3.9.4 Pathway and GO Analysis of Common Genes Linked to CTA

The pathway analysis identified significant enrichment in key biological pathways, including the p53 signaling pathway, MAPK signaling pathway, and NF-kappa B signaling pathway, which are critical in regulating apoptosis, cell proliferation, and inflammatory responses, respectively (Figure 12). Additionally, Fc gamma R-mediated phagocytosis and aldosterone-regulated sodium reabsorption were highlighted, suggesting their potential roles in cellular homeostasis and signaling relevant to tooth development (Figure 12). Leishmaniasis, malaria, hepatitis B, and pancreatic secretion pathways were also observed, possibly reflecting systemic interactions affecting congenital tooth agenesis (Figure 12). These findings underscore the involvement of diverse molecular mechanisms in the etiology of congenital tooth agenesis.

**Figure 12:**
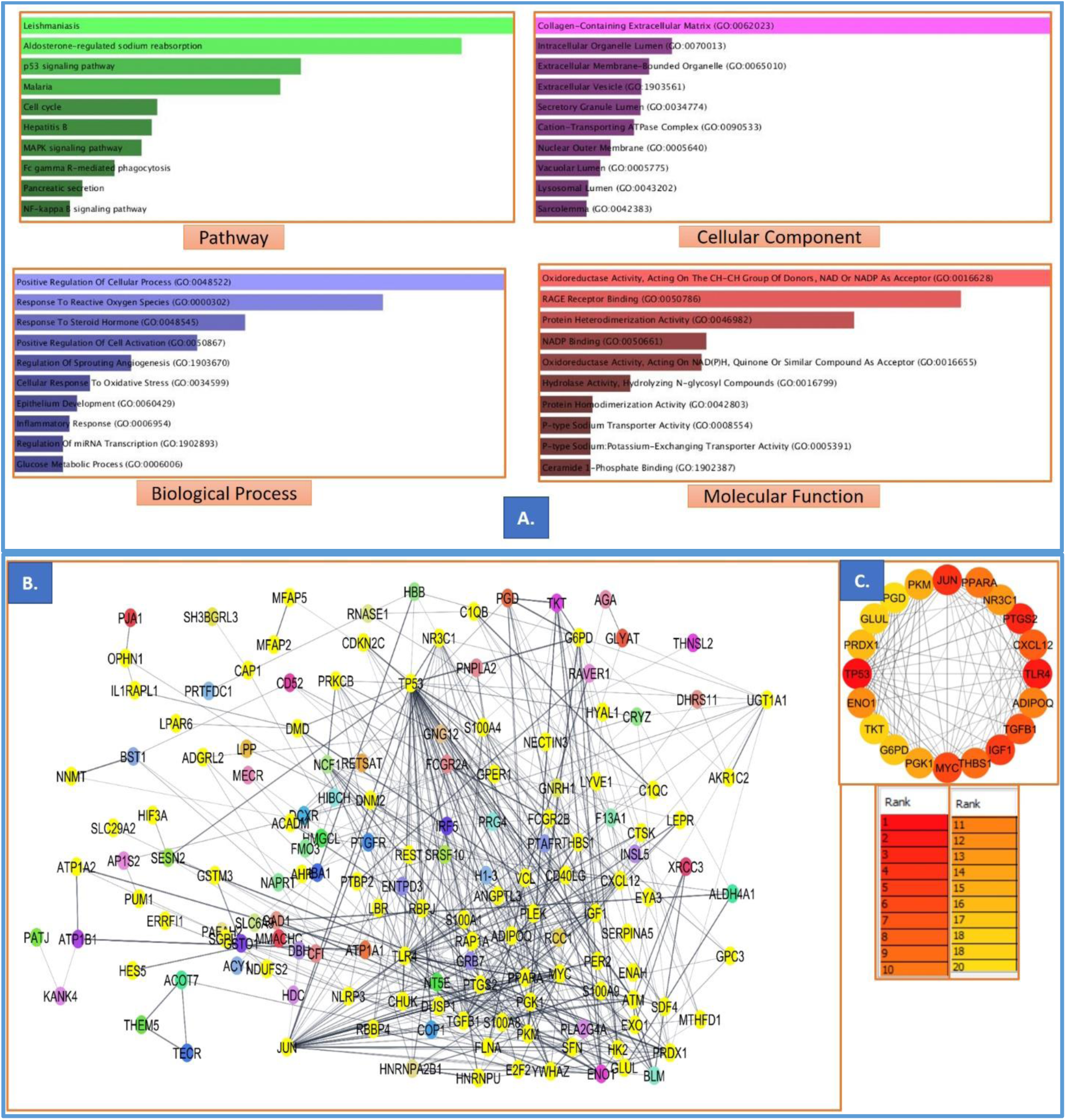
GO, Pathway and Network analysis of integrated common Gene Associated with CTA. (A) Pathways: Significant pathways include aldosterone-regulated sodium reabsorption, p53 signaling, and NF-κB signaling, emphasizing key regulatory and signaling processes involved in tooth development. Enrichment categories: Biological Process, Cellular Component, and Molecular Function. B. Network Analysis of Common Genes Across All Panels. It illustrates the network analysis performed using STRING on the common genes identified across all panels. The nodes in the network represent individual genes, and the connections between them indicate known or predicted interactions. The yellow-highlighted nodes denote genes that play a role in development, emphasizing their significance in the context of congenital tooth agenesis. C. The analysis also identifies the top 20 hub genes within the network, which are genes with the highest degree of connectivity. These hub genes are pivotal in the network and likely play crucial roles in the underlying biological processes associated with congenital tooth agenesis.

The analysis identified significant cellular components, including the collagen-containing extracellular matrix, intracellular organelle lumen, extracellular vesicle, and secretory granule lumen, which play critical roles in structural support, molecular transport, and protein secretion during tooth development. Other enriched components, such as the lysosomal lumen, nuclear outer membrane, and sarcolemma, were associated with protein degradation, signaling, and mechanical stability essential for odontogenesis (Figure 12).

Cellular component analysis highlighted the significance of collagen-containing extracellular matrix and intracellular organelles in developmental processes. The analysis identified significant biological processes, including positive regulation of cellular processes, response to reactive oxygen species, response to steroid hormones, positive regulation of cell activation, regulation of sprouting angiogenesis, cellular response to oxidative stress, epithelial development, inflammatory response, regulation of miRNA transcription, and glucose metabolic processes (Figure 12). These processes highlight key roles in cellular regulation, stress response, hormone signaling, angiogenesis, and metabolism.

Several significant molecular functions (Figure 12) were identified in the context of CTA, including oxidoreductase activities for redox processes, RAGE receptor binding associated with inflammatory responses, protein hetero-/homodimerization activities essential for protein interactions in odontogenesis, NADP binding related to energy metabolism, P-type transporter activities for ion homeostasis in mineralization, and ceramide-1-phosphate binding involved in lipid signaling pathways. Disease association analysis of integrated genes revealed a predominant link to ectodermal and mesodermal origins.

STRING-based network analysis of common genes across all panels (Figure 12) demonstrated interconnected gene networks, with nodes representing individual genes and edges indicating known or predicted interactions. Genes associated with developmental processes were highlighted in yellow, emphasizing their importance in the context of congenital tooth agenesis. Key genes, including *JUN, PPARG, NR3C1, PTGS2, CXCL12, TLR4, ADIPOQ, TGFB1, IGF1, THBS1, MYC, PGK1, G6PD, TKT, ENO1, TP53, PRDX, GLUL, PGD,* and *PKM*, were identified as being involved in crucial biological processes and pathways related to tooth development (Figure 12). Their interactions and functions provided valuable insights into developmental mechanisms. Among the 20 analyzed genes, *JUN, TP53, MYC, IGF1*, and *TLR4* were identified as pivotal in tooth development due to their significant roles and high connectivity within the network (Figure 12).

### 3.10 Dual expression Genes

The analysis highlights significant findings regarding dual expression genes and their correlation during human embryonic tooth germ development. Five common genes were identified between “Human Embryonic Tooth DEGs” and “Proteome Down DEGs,” including *MST1*, *KRT1*, *KRT10*, *KRT2*, and *KRT14*. Additionally, nine genes overlapped between “Human Embryonic Tooth DEGs” and “Proteome UP DEGs,” such as *PLEK*, *ACTC1*, *HBD*, *ACTN1*, *HBB*, *CA2*, *LYVE1*, *TLN1*, and *ALDOA* (Figure 13). Furthermore, a total of 24 genes were found to be common between “Human Embryonic Tooth DEGs” and the “Integrated Panel.” These include *PTGS2, DUSP1, RNASE1, ACADM, C1QB, PLEK, CXCL12, GAD1, MST1, KRT1, HDC, HBB, PRKCZ, MFAP5, LYVE1, ERRFI1, C1QC, BBP4, HNRNPA1B1, FOXN2, NT5E, HIF3A, PAI1,* and *IFNLR1*(Figure 13).

**Figure 13:**
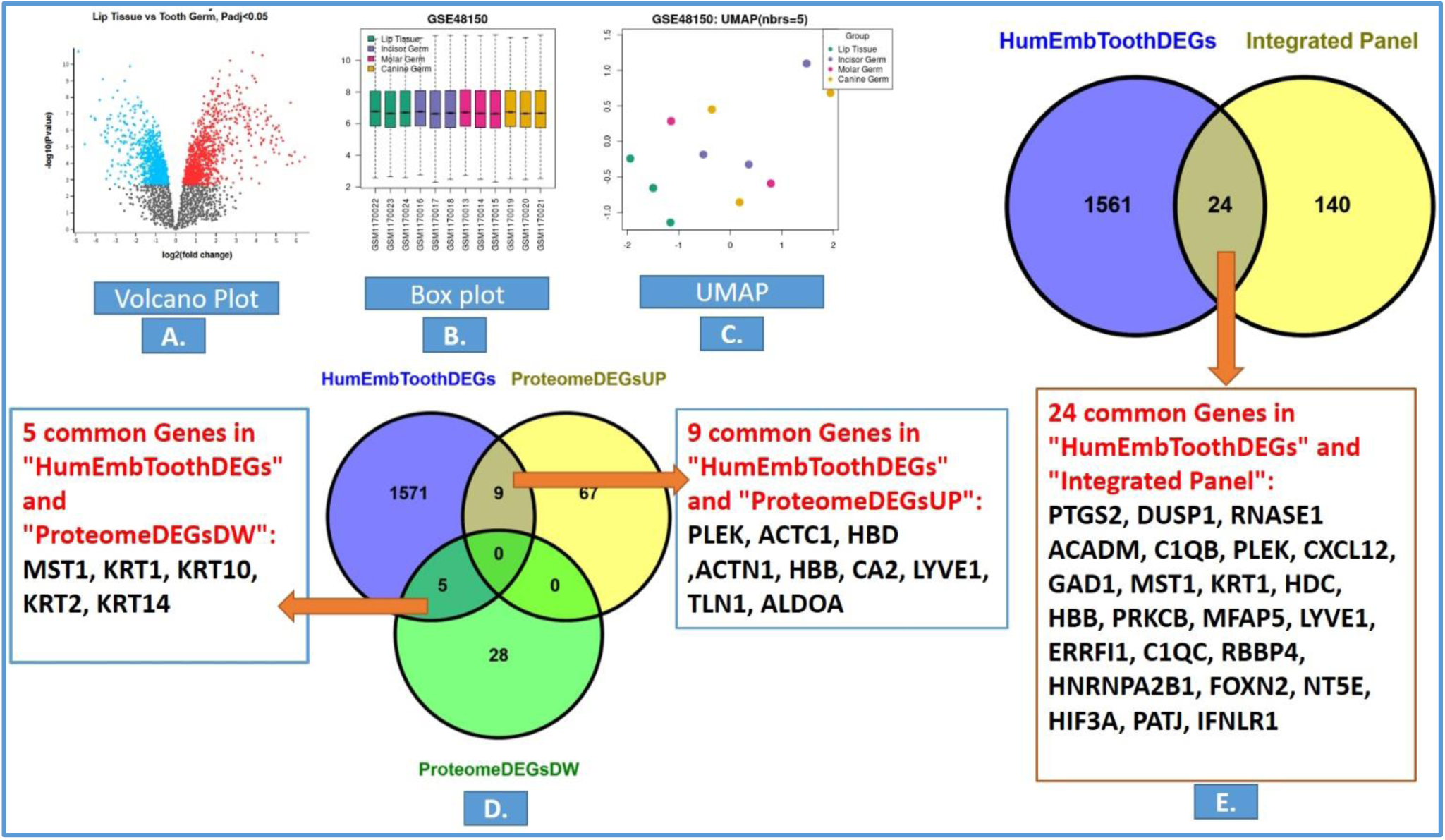
Microarray analysis of human embryonic tooth germ and correlation with integrated panel and proteomic panel for the discovery of dual expression genes expressed during embryonic and post-expression stages. A. Volcano Plot: This plot presents the differential gene expression in human embryonic tooth germ compared to control samples. Genes with significant changes in expression (p-value < 0.05) are highlighted. B. Box Plot: This plot illustrates the distribution of gene expression levels across various samples in the dataset, highlighting the variability and median expression values. C. UMAP Plot: Uniform Manifold Approximation and Projection (UMAP) is used to visualize the clustering of samples based on their gene expression profiles, showing distinct groups and relationships among the samples. D: Correlation of Proteomic Panel to Human Embryonic Tooth Germ DEGs: A Venn diagram displays the overlap between differentially expressed genes (DEGs) identified in the proteomic panel and those found in the human embryonic tooth germ. Yellow- highlighted genes indicate those involved in developmental processes. E: Correlation with Human Embryonic Tooth Germ DEGs to Integrative Panel: A Venn diagram shows the correlation between the DEGs in the human embryonic tooth germ and the integrative panel, identifying common genes that play a role in both embryonic development and post-development stages.

The overlapping genes underscore their potential regulatory role in tooth development, with key players like *MST1*, *KRT1*, and *PLEK* showing consistent dual expression across datasets. These findings provide valuable insights into gene expression dynamics, emphasizing the significance of overlapped genes as biomarkers for studying the molecular mechanisms of human embryonic tooth germ development.

The network, GO and pathway analysis of the 24 dual expression genes revealed significant insights into their role in tooth development. Network analysis demonstrated interactions among these genes, highlighting their regulatory connections (Figure 14). Pathway analysis identified key pathways involved in CTA, including NF-kappa B (tooth germ regulation), VEGF (angiogenesis in tooth development), HCMV infection (neural crest disruption), and serotonergic synapse (craniofacial patterning) (Figure 14). Biological process analysis indicated that Cell Junction Disassembly (GO:0150146) is crucial for remodeling cell junctions during dental tissue development (Figure 14). Cellular component analysis showed that the Collagen-Containing Extracellular Matrix (GO:0062023) plays a vital role in dental tissue formation. Molecular function analysis revealed that Protein Binding (GO:0005515) and Calcium Ion Binding (GO:0005509) are essential for processes related to tooth development (Figure 14).

**Fig 14.**
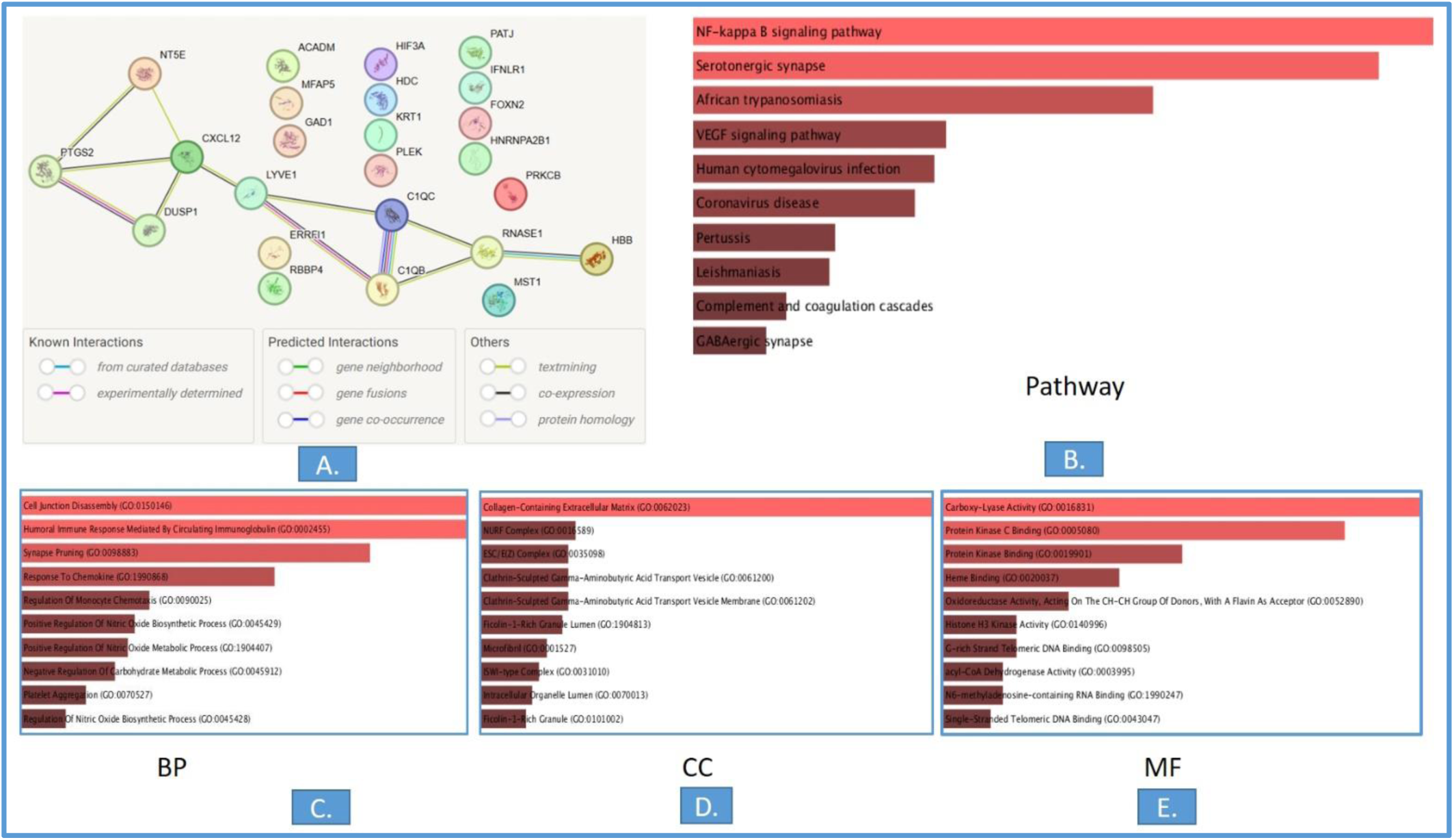
Network and Enrichment Analysis of Obtained 24 Dual Expression Genes: A. This panel illustrates the network analysis of 24 dual expression genes. The network map highlights the interactions between these genes, with nodes representing the genes and edges representing the known or predicted interactions. B. Pathway Analysis: Key pathways in CTA include NF-kappa B (tooth germ regulation), VEGF (angiogenesis in tooth development), HCMV infection (neural crest disruption), and serotonergic synapse (craniofacial patterning). C. Biological Process (BP) Analysis: Cell Junction Disassembly (GO:0150146) is directly involved in tooth formation as it plays a role in the remodeling of cell junctions during the development of dental tissues. D. Cellular Component Analysis: Collagen-Containing Extracellular Matrix (GO:0062023) is directly linked to tooth formation as it plays a crucial role in the development of dental tissues. E. Molecular Function Analysis: Protein Binding (GO:0005515) and Calcium Ion Binding (GO:0005509) were directly linked to tooth development.

### 3.11 CTA associated with Ecto-Mesodermal Diseases

Disease association analysis was performed using metabolome, proteome, genomic, and gene expression data. Analysis of individual and integrated omics datasets identified the top 10 most significant diseases, predominantly linked to ectodermal and mesodermal disorders. Proteome-based analysis of up- and downregulated germ layer genes predominantly indicates associations with ectodermal and mesodermal diseases (Table 2 and Supplementary information Table1).

## 4. Discussion

The present study provides a comprehensive pan-omics analysis of CTA, integrating metabolomics, proteomics, microarray, and genomic data to elucidate the molecular mechanisms underlying this condition. Our findings reveal significant dysregulation in metabolic and proteomic profiles, identify key biomarkers followed by machine learning and integrative omics approach, and highlight critical pathways and gene networks implicated in CTA pathogenesis. This study also reveals the ecto-mesodermal connection to diseases and identifies dual-expression genes, enhancing our understanding of the molecular basis of CTA while also offering potential diagnostic and therapeutic targets.

The identification of 28 metabolites exclusively dysregulated in the UP-regulated group and 17 in the Down-regulated group further emphasizes the divergent metabolic signatures associated with these regulatory states. The analysis of metabolite dysregulation in the study samples compared to the Known Ecto-Mesodermal Metabolites Panel and experiment output results revealed significant insights into the distinct metabolic alterations associated with the study groups. The overlap of three metabolites (inositol, sebacic acid, and L-Leu-Lu) between the Known Ecto-Mesodermal Metabolite Panel and the UP-regulated group suggests a shared metabolic pathway that may be critical for maintaining cellular integrity during ectodermal and mesodermal differentiation. Inositol, in particular, has been widely studied for its role in cell signaling and membrane biosynthesis, while sebacic acid is known for its involvement in lipid metabolism (Jovanovic et al., 2022; Liao et al., 2024) Similarly, the overlap of testosterone, benzoic acid, and uric acid between the Known Ecto-Mesodermal Metabolite Panel and the Down-regulated group highlights their potential role in metabolic dysregulation, particularly in processes related to oxidative stress and inflammation(Acharya et al., 2024; Jung et al., 2020). These findings highlight the potential significance of these metabolites in the metabolic pathways that underpin ectodermal and mesodermal differentiation, shedding light on their possible contributions to the development of CTA. In our study, we identified a set of unmatched metabolites that were not present in the known ectodermal and mesodermal metabolite panels. These unmatched metabolites may serve as novel biomarkers with potential specificity for congenital tooth agenesis. Given that tooth development is a complex process involving interactions between ectodermal and mesodermal tissues, the presence of these unique metabolites suggests that they could play a previously unrecognized role in the molecular mechanisms underlying tooth formation.

The crucial significance that nucleotide and purine metabolism pathways play in DNA and RNA synthesis as well as cellular energy metabolism is highlighted by the overexpression of these pathways in CTA patients. Dysregulation of these pathways may hinder the growth and differentiation of dental epithelial and mesenchymal cells, resulting in tooth agenesis. These pathways are crucial for preserving appropriate cellular function during development(Zuccarini et al., 2022). On the other hand, the downregulation of pathways including purinergic signaling, caffeine metabolism, and IL-10 anti-inflammatory signaling points to problems with DNA repair processes, inflammatory responses, and cellular communication. These results are consistent with other studies showing that abnormalities in these pathways can lead to developmental abnormalities and jeopardize cellular homeostasis(Boccazzi et al., 2023; Gibbs et al., 2015; Zhu et al., 2018) .

The identification of specific metabolites, such as 6-Hydroxycaproic acid, Stearamide, Dihydrothymine, Uric acid, Inosine, and Docosahexaenoic acid (DHA), further supports the involvement of lipid and purine metabolism, as well as Wnt and hormonal signaling pathways, in CTA. These metabolites have been previously associated with developmental processes and cellular differentiation, suggesting their potential role in tooth development (Barnes et al., 2009; Kim et al., 2011; Pieles et al., 2022; Tabakcilar et al., 2020; Tardieu et al., 2017; Van den Berghe & Vincent, 1995; Williams & Letra, 2018; Ye et al., 2023). On the other hand, downregulated metabolites like PEGs, bilirubin, and caffeine require further investigation to elucidate their specific contributions to CTA pathogenesis. The upregulated DHA metabolite has been associated with the Wnt signaling pathway and NF-κB signaling, both of which play critical roles in tooth development. Specifically, DHA enhances the Wnt pathway, leading to increased cellular proliferation, a vital process during tooth morphogenesis. Additionally, DHA modulates NF-κB signaling by inhibiting the phosphorylation of IκB, thereby sequestering NF-κB in the cytoplasm and preventing its nuclear translocation. This regulation of NF-κB activity is essential for maintaining a balance between inflammation and cellular homeostasis, which are crucial during tissue development.

The quantitative metabolite analysis revealed unique metabolic profiles between control and CTA patient samples, emphasizing significant biochemical alterations associated with the condition. The higher number of unique metabolites in patient samples (292 metabolites, 25.9%) compared to control samples (238 metabolites, 21.1%) suggests disease-specific metabolic shifts that may contribute to tooth development abnormalities. The identification of 587 metabolites (52.1%) within the known ecto-mesodermal metabolite panel further supports the relevance of these metabolic pathways in dental tissue formation and function.

A key observation is the minimal overlap of metabolites between the known panel and control (0.3%) or patient (0.6%) samples, indicating that most metabolic variations are unique to either physiological or pathological states.

In control samples, the presence of 3-Hydroxytetradecanedioic acid, Leu-Val, and Palmitoleic acid may reflect normal fatty acid metabolism, amino acid processing, and lipid homeostasis, essential for cellular integrity and differentiation. Conversely, the seven overlapped metabolites in patient samples were Pyridinolone, Vitamin A, Citric acid, Benzaldehyde, Creatine, Phenylglyoxylic acid, and Olomoucine. This suggests significant metabolic disturbances. Notably, Vitamin A and Citric acid play crucial roles in odontogenesis, cellular differentiation, and energy metabolism, while Creatine is linked to cellular energy homeostasis, which may be disrupted in CTA (Gualano et al., 2010; Hristov et al., 2024)The presence of Phenylglyoxylic acid and Benzaldehyde in patient samples suggests potential alterations in amino acid metabolism and oxidative stress pathways, which could contribute to abnormal dental development (Kim et al., 2011, 2012). Additionally, Olomoucine, a cyclin-dependent kinase inhibitor, raises the possibility of disrupted cell cycle regulation in tooth formation (Y. Chen et al., 2022).

The findings from this study provide significant insights into the metabolic alterations associated with CTA, highlighting distinct dysregulations in key pathways that may contribute to the etiology of this condition. By mapping unique metabolites identified in patient samples to various metabolic pathways, we observed significant perturbations in lipid metabolism, vitamin metabolism, amino acid transport, inflammatory regulation, pluripotency/differentiation, energy metabolism, and hormonal signaling. These dysregulated pathways collectively suggest a complex interplay of metabolic and signaling mechanisms underlying CTA, consistent with previous studies that have implicated metabolic dysregulation in developmental disorders(Oyarzábal et al., 2021). These findings highlight potential biomarkers that distinguish CTA from normal dental development and suggest metabolic pathways that may underlie the disease pathology.

The MetScape analysis revealed intricate metabolite-metabolite interactions, providing a network perspective on how these metabolites may collectively influence molecular mechanisms underlying CTA. This approach aligns with recent trends in systems biology, which emphasize the importance of understanding metabolic networks in complex diseases (X. Wang et al., 2020). Proteomic analysis reveals significant differences in protein expression profiles between CTA patients and controls, emphasizing the altered proteomic landscape in CTA. Differentially expressed proteins, as shown by volcano plots and PCA, indicate distinct clustering of patient samples, reflecting disease-specific protein expression. The enriched biological processes associated with upregulated proteins include cell adhesion, immune response, and hemostasis, potentially highlighting compensatory or maladaptive responses to structural abnormalities in dental tissues. Conversely, downregulated proteins were linked to critical processes such as peptide cross-linking and intermediate filament organization, essential for maintaining tissue integrity during odontogenesis. These proteomic alterations align with observed developmental disruptions in CTA and suggest a molecular basis for impaired dental tissue development. The gene network analysis provided key insights into the differential regulation and hub genes involved in CTA. The combined gene network, generated using STRING, highlighted several developmental process-associated genes, underscoring their potential roles in CTA. Hub gene analysis identified highly connected genes such as *ACTB*, *ENO1*, *GAPDH*, and *TPI1*, emphasizing their central roles in the network and suggesting their importance in CTA pathophysiology. The Venn diagram analysis illustrated gene overlaps among the OMIM, Proteomics, and Gene Card panels. Notably, *COLEC11* was identified as the only common gene across all three panels, suggesting its critical role in CTA. The Proteomics Panel contained 76 novel genes, while the Gene Card Panel had 4759 unique genes. Additionally, there were 32 genes overlapping between the Proteomics and Gene Card Panels and 126 genes common between the OMIM and Gene Card Panels. These findings highlight key genes and interactions that may contribute to the molecular mechanisms underlying CTA.

The application of machine learning techniques to proteome and metabolite data analysis has yielded significant insights into the molecular mechanisms CTA. The robust performance of the models in distinguishing between “Control” and “Sample” groups underscores the potential of these approaches in identifying reliable biomarkers and understanding the pathophysiology of CTA.

The proteome data analysis demonstrated excellent discrimination between “Control” and “Sample” groups, with an AUC-ROC score of 0.95. The high sensitivity (0.92) and specificity (0.94) of the model indicate its strong capability to accurately classify the samples. This analysis identified multiple candidate biomarkers for CTA, with high-performing biomarkers such as *SERPINA1*, *PZP*, *FGA*, *IGHM, TLN1*, and *FGB* showing AUC values above 0.85. These biomarkers exhibit strong sensitivity and specificity, making them valuable for CTA diagnosis and further investigation into their roles in the disease. Moderate-performing proteins, including *KRT2, PRDX2*, *HBB*, *GAPDH*, and *FLNA*, also contribute to the understanding of CTA but with relatively lower performance. Low-performing proteins, with AUC values below 0.70, were less reliable for CTA classification, suggesting limited diagnostic utility.

The metabolite data analysis exhibited exceptional performance, with an AUC-ROC score of 0.98. The model’s high sensitivity (0.96) and specificity (0.97) reflect its robust discriminatory power. High-performing metabolites such as PEG n5, PEG n6, PEG-4, PEG n7, PEG n8, and caffeine demonstrated AUC values above 0.90, indicating their strong potential as biomarkers for CTA. Additionally, metabolites like NP-007909 (unknown), hydroxycaproic acid, and α- aspartylphenylalanine showed significant discriminatory potential. Moderate-performing metabolites, including N-(1,2-diphenylethyl)-N’-(4-fluorobenzyl) thiourea, 3-oxoindane-1- carboxylic acid, and benazol P, exhibited a balance between sensitivity and specificity. However, low-performing metabolites, such as leu-phe and leu-leu, with AUC values near or below 0.50, suggest limited diagnostic utility. These findings highlight the importance of selecting high-performing biomarkers for reliable CTA diagnosis and theurapeutics.

The six overlapping genes (*F13A1, PLEK, LYVE1, BST1, YWHAZ, S100A4*) between Microarray and Proteomics UP DEGs highlight perturbations in immune regulation and extracellular matrix (ECM) remodelling (W. Chen & Zhang, 2016; D’Ambrosi et al., 2021; Elfstrum et al., 2024; Kaartinen et al., 2020) *S100A4* and *YWHAZ* are particularly notable, as they modulate cell proliferation, apoptosis, and Wnt/β-catenin signaling—pathways critical for dental lamina formation and enamel knot patterning (Dahlmann et al., 2016; Liu & Millar, 2010). Similarly, the 55 Microarray UP-Metabolite Panel genes (*JUN, MYC, PTGS2, HIF3A*) implicate hypoxia-responsive and inflammatory pathways, potentially disrupting epithelial-mesenchymal interactions essential for tooth germ development (Tam et al., 2020)(Puthiyaveetil et al., 2016). The downregulated *PPARG* and *G6PD* (Microarray Down-Metabolite Panel) suggest compromised lipid metabolism and redox homeostasis, which may impair ameloblast/odontoblast differentiation(H.-C. Yang et al., 2019)(Ahmadian et al., 2013) .

The extensive overlap between Microarray DEGs and the Metabolite Panel (111 genes) emphasizes metabolic reprogramming as a hallmark of CTA (C. Yang et al., 2023). *TKT*, shared across Proteomics, Microarray, and Metabolite Panels, links the pentose phosphate pathway to nucleotide biosynthesis and oxidative stress management (Qin et al., 2019). Its dysregulation could hinder dental progenitor cell proliferation. Similarly, *PKM* and *ENO1* (Proteomics UP-Metabolite Panel) are glycolysis regulators, suggesting that altered glycolytic flux may disrupt energy-dependent morphogenetic processes during tooth development (Nieborak et al., 2023)(Lemaire & Wésolowski-Louvel, 2004).

Genes overlapping with the WES Panel (*FLNA, VCL, IGF1*) point to cytoskeletal organization and mechanotransduction defects. *FLNA* and *VCL* are pivotal for cell adhesion and migration, processes vital for neural crest cell-derived dental mesenchyme organization (Zhou et al., 2021)(Petrusová et al., 2022). *IGF1*, a growth factor gene, further connects insulin-like signaling to odontogenic cell survival and differentiation, potentially explaining enamel hypoplasia in CTA (Oyanagi et al., 2019). While canonical CTA genes (*MSX1, PAX9, AXIN2*) were not identified here, the integrative approach uncovered novel candidates. For instance, *DUSP1* (Microarray- WES overlap) regulates MAPK signaling, which governs tooth patterning (Khadir et al., 2015), while *PRG4* (Proteomics Down-Metabolite Panel) modulates lubricin expression, potentially affecting joint and dental follicle homeostasis (Krawetz et al., 2022). The inflammasome-related *NLRP3* (Microarray UP-Metabolite Panel) and *S100A8/S100A9* (Proteomics UP- Metabolite Panel) suggest chronic inflammation may exacerbate developmental arrest in dental tissues (Simard et al., 2013).

The enrichment of the p53 signaling pathway highlights its role in balancing apoptosis and cell proliferation during tooth morphogenesis (Guo et al., 2024). Dysregulation of p53 (*TP53*) may disrupt dental epithelial cell survival, leading to arrested tooth bud formation. Similarly, the MAPK pathway (via *JUN*) and NF-kappa B signaling (via *TLR4*) are central to epithelial-mesenchymal interactions, which orchestrate enamel knot patterning and crown formation (Nyati et al., 2017).

Aberrant NF-kappa B activation could exacerbate inflammatory responses, impairing the delicate signaling milieu required for dental lamina invagination (Yu et al., 2022). The unexpected association with Fc gamma R-mediated phagocytosis and aldosterone-regulated sodium reabsorption suggests broader systemic influences, such as immune cell activity or ion homeostasis, that may indirectly affect mineralization processes in developing teeth (Indik et al., 1995). The prominence of the collagen-containing extracellular matrix (ECM) aligns with its role in providing structural scaffolding for enamel and dentin deposition (Cruz Walma & Yamada, 2022). Disruptions in ECM organization, potentially via *THBS1* or *TGFB1*, could compromise odontoblast differentiation (Pal et al., 2016). Enrichment of lysosomal lumen components points to altered protein degradation, critical for remodeling the dental basement membrane during eruption (Paudel et al., 2022).

The response to reactive oxygen species (ROS) and oxidative stress (*PRDX*, *G6PD*) underscores the vulnerability of dental progenitors to redox imbalances, which can impair cell proliferation and enamel maturation (Ni et al., 2019; Qiu et al., 2025). Steroid hormone response (*NR3C1*) suggests glucocorticoid signaling may influence tooth germ development (Holmes Jr et al., 2019), while glucose metabolic processes (*PGK1*, *PKM*, *ENO1*) emphasize glycolysis as an energy source for amelogenesis (Jin et al., 2020; Luo et al., 2024; Puckett et al., 2021). Angiogenesis (*CXCL12*) likely supports nutrient delivery to the avascular enamel organ, with dysregulation potentially leading to hypoplastic defects (Salcedo & Oppenheim, 2003).

Oxidoreductase activity (*GLUL*, *G6PD*) and NADP binding highlight redox and metabolic maintenance (Stanton, 2012), while P**-**type transporter activity (*ATP1A1*) links ion transport to enamel mineralization (Stafford et al., 2017). RAGE receptor binding (*S100A8/A9*) implicates chronic inflammation in disrupting odontogenic signaling (S. Wang et al., 2018). The ectodermal-mesodermal disease associations reinforce the dual origin of dental tissues, with neural crest-derived mesenchyme and oral epithelium being central to CTA pathogenesis (Miletich & Sharpe, 2004).

The interconnected network of hub genes (*JUN*, *TP53*, *MYC*, *IGF1*, *TLR4*) underscores their centrality in CTA. *IGF1* governs growth and differentiation of dental papilla cells (Oyanagi et al., 2019), while *MYC* and *JUN* drive proliferation via MAPK/c-MYC axes (Stefan & Bister, 2017). *TLR4* and *PTGS2* (COX-2) bridge inflammation and developmental arrest, potentially explaining agenesis in syndromic CTA (W. Wang & Wang, 2018). Metabolic enzymes (*TKT*, *PGD*) further integrate redox balance with nucleotide biosynthesis, essential for progenitor cell expansion (Y. Chen et al., 2022; Ju et al., 2020).

The overlap of genes such as *KRT14*, *CXCL12*, *LYVE1*, and *PTGS2* across developmental and post-developmental contexts suggests their dual functionality. For instance, *KRT14*, a keratin critical for epithelial integrity during enamel knot formation (Inada et al., 2024). Similarly, *CXCL12*, a chemokine enriched in the NF-kappa B pathway, not only guides neural crest cell migration during development but also recruits mesenchymal stem cells (MSCs) to sites of dental injury, facilitating pulp regeneration (Noort et al., 2014). The persistence of these genes post-development underscores their role in maintaining a reparative niche, potentially compensating for minor tissue damage.

Genes like *PLEK* and *LYVE1*, associated with the Collagen-Containing Extracellular Matrix (GO:0062023), highlight the ECM’s dual role as a scaffold for morphogenesis and a dynamic substrate for repair. *LYVE1*, a marker of lymphatic endothelial cells, may regulate immune cell trafficking during both developmental patterning and post-injury inflammation resolution (Elfstrum et al., 2024)(F. Wang et al., 2021). *ACTN1* and *TLN1*, which modulate actin cytoskeletal reorganization, may essential for odontoblast process elongation during dentinogenesis. (Hamill et al., 2015)(Conti et al., 2009). These findings align with studies showing that ECM remodeling genes are re-expressed in odontoblasts following injury to promote matrix deposition and mineralization (Bonnans et al., 2014).

The dual-expression gene *PTGS2* (COX-2), a key mediator of prostaglandin synthesis, exemplifies the fine balance between developmental signaling and repair. While *PTGS2* drives inflammatory signaling in CTA pathogenesis, its transient upregulation post-injury is crucial for initiating pulp repair via prostaglandin E2 (PGE2)-mediated MSC recruitment (Martín-Vázquez et al., 2023). Similarly, *HIF3A*, a hypoxia-inducible factor, may not only mediate adaptive responses during enamel organ vascularization but also enhance glycolysis in dental pulp stem cells under hypoxic conditions, promoting their survival and differentiation during repair (Holomková et al., 2024).

The keratins *KRT1*, *KRT2*, and *KRT10*, downregulated in Proteome Down DEGs, are vital for epithelial barrier function. Their reduced expression in CTA could predispose dental epithelia to structural fragility, while their re-activation in adult tissues may reinforce barrier integrity after mechanical or microbial insult (Hotz et al., 2016).

CTA is a developmental anomaly often studied in isolation; however, its association with ectodermal and mesodermal diseases suggests broader systemic implications. The current study integrates metabolome, proteome, genomic, and gene expression data to identify the top 10 significant diseases associated with CTA, predominantly linked to ectodermal and mesodermal disorders. This reinforces the hypothesis that dental anomalies may serve as early indicators of underlying systemic conditions affecting multiple germ layers.

Proteome-based analysis further supports this observation, revealing that differentially expressed germ layer genes are primarily associated with ectodermal and mesodermal diseases. Given that tooth development arises from ectomesenchymal interactions, disruptions in these pathways could contribute to both CTA and systemic conditions such as ectodermal dysplasia and mesodermal connective tissue disorders (Santosh & Jones, 2014). These findings align with previous studies highlighting the shared genetic and molecular pathways between dental anomalies and broader developmental syndromes (Chetty et al., 2021).

Dental implantation remains a common solution for missing teeth, yet the broader implications of CTA as a potential marker for ectomesodermal diseases have not been extensively explored. This study provides novel insights into the systemic nature of CTA, emphasizing that missing teeth may not be an isolated dental issue but rather a clinical manifestation of broader developmental disorders. Recognizing these associations could enhance early diagnosis and targeted therapeutic approaches, benefiting patients beyond routine dental care.

While this study provides significant insights into the molecular mechanisms of CTA, further research is needed to validate these findings in larger cohorts and diverse populations. Functional studies on key genes and metabolites identified in this analysis could elucidate their precise roles in odontogenesis and craniofacial development. Additionally, the development of targeted therapies based on identified pathways could offer promising avenues for clinical intervention. Integration of other omics approaches, such as epigenomics may further enhance our understanding of the complex molecular networks underlying CTA. Understanding CTA in a pan- omics context could pave the way for precision medicine approaches in dental and systemic health management.

## 5. Conclusion

This study presents a comprehensive pan-omics fusion approach, integrating metabolomics, proteomics, Gene expressions, and genomics with machine learning, to uncover the molecular mechanisms underlying CTA. Our findings reveal significant metabolic and proteomic dysregulations, highlighting key biomarkers and critical pathways involved in ecto**-**mesodermal interactions, nucleotide metabolism, oxidative stress, and Wnt signaling. The identification of novel metabolites and previously unrecognized hub genes (*COLEC11, ACTB, ENO1, GAPDH, TPI1*) provides deeper insights into the systemic nature of CTA, reinforcing its potential association with broader ectodermal and mesodermal diseases. Machine learning-based biomarker discovery yielded high classification accuracy (AUC**-**ROC: 0.95–0.98), emphasizing the diagnostic potential of *SERPINA1, TLN1, IGHM,* and *DHA* as reliable CTA markers. The minimal overlap between known ecto-mesodermal metabolite panels and CTA patient-specific metabolic profiles suggests the existence of CTA**-**unique metabolic pathways, which may be crucial for its pathogenesis. Furthermore, the downregulation of critical immune and inflammatory pathways (NF**-**κB, IL**-**10, and purinergic signaling) suggests that disruptions in cellular communication and immune regulation play a role in CTA development.

Our findings support the hypothesis that CTA is not merely an isolated dental anomaly but may serve as an early clinical marker for systemic ecto-mesodermal disorders. This expands the clinical significance of CTA beyond oral health, emphasizing the need for interdisciplinary diagnostic strategies that consider potential systemic implications. Notably, 24 dual-expression genes were found to mediate both developmental and reparative processes, suggesting a crucial role in regulating tooth morphogenesis, differentiation, and maintenance. During the pre-developmental stage, it influences cellular proliferation, signaling pathways, and tissue patterning. In the post-developmental stage, it contributes to tooth integrity, repair, and homeostasis.

## Data Availability

All data produced in the present study are available upon reasonable request to the authors

## Acknowledgement

We gratefully acknowledge the Indian Council of Medical Research (ICMR) for providing the Senior Research Fellowship (SRF) to PR and CD. We acknowledge the SATHI-BHU at Central Discovery Centre (CDC), BHU for the HRAMS and LC-MS facilities.

## 6. Author Contribution

**PR** conceived the research idea, conducted experiments, performed bioinformatics and data analysis, and wrote the manuscript. **CD** contributed to data analysis. **VKS, MC** was responsible for identifying CTA patients and conducting OPG analysis. **GJ** Data curation **PD** co-conceived the research idea and approval of final draft.

## 7. Declaration

### Consent to participate

All subjects participating in this study were fully informed about the purpose, process, potential risks and benefits of the study and voluntarily signed a written informed consent form.

## List of Abbreviations

AUC-ROC: Area Under the Receiver Operating Characteristic Curve
BMP: Bone Morphogenetic Protein
BP: Biological Process
CC: Cellular Component
CTA: Congenital Tooth Agenesis
DEGs: Differentially Expressed Genes
FDR: False Discovery Rate
GEO: Gene Expression Omnibus
GO: Gene Ontology
KEGG: Kyoto Encyclopedia of Genes and Genomes
LC-MS: Liquid Chromatography-Mass Spectrometry
Log2FC: Log2 Fold Change
OMIM: Online Mendelian Inheritance in Man
OPGs: Orthopantomograms
PCA: Principal Component Analysis
q-value: Adjusted p-value (FDR-corrected)
RMA: Robust Multi-array Average
ROC: Receiver Operating Characteristic
WES: Whole Exome Sequencing

