## Supplementary material for "Pan-Omics Fusion and Machine Learning Unveil Congenital Tooth Agenesis-Ecto-mesodermal Diseases Link and Biomarker Discovery": Supplimentary_table1

**Table1:** **Pathway analysis of unique metabolites present in control and patient samples and their comparative analysis:** illustrates the comparative pathway analysis of unique metabolites present in control and patient samples, supporting the characterization and understanding of CTA. The table categorizes pathways based on their specific involvement in control and patient samples, highlighting the metabolic alterations and their implications.

| **Pathway Category** | **Control-Specific Pathways** | **Patient-Specific Pathways** | **Description** |
| --- | --- | --- | --- |
| Lipid Metabolism | Omega-9 fatty acid synthesis | Fatty acid biosynthesis and metabolism | - Control: Supports cell membrane integrity and energy storage. - Patient: Defective lipid signaling and energy production. |
| Vitamin Metabolism | Vitamin A and carotenoid metabolism | Vitamins A and D - action mechanisms | - Control: Critical for epithelial cell differentiation and enamel formation. - Patient: Altered dynamics disrupt epithelial differentiation and mineralization. |
| Amino Acid Transport | - | Amino acid transport defects (IEMs) | - Control: Not specified. - Patient: Essential for protein synthesis in odontogenesis. |
| Inflammatory Regulation | - | Arachidonic acid metabolism | - Control: Not specified. - Patient: Regulates inflammatory processes and tissue repair during tooth development. |
| Pluripotency/Differentiation | Embryonic stem cell pluripotency pathways | RA (Retinoic Acid) biosynthesis | - Control: Crucial for early cellular differentiation in dental tissue formation. - Patient: Altered RA levels affect odontoblast differentiation and dentinogenesis. |
| Energy Metabolism | Cholesterol metabolism | Citric acid cycle and respiratory transport | - Control: Supports membrane structure and signaling critical for tooth morphogenesis. - Patient: Increased cellular stress and compromised energy production. |
| Hormonal Pathways | - | Peptide hormone metabolism | - Control: Not specified. - Patient: Impaired signaling pathways affect tooth development and homeostasis. |
